# Plasma Proteomic Analysis of *APOE* ε4 Homozygotes Identifies Preclinical Alzheimer’s Disease Alterations Potentially Modulated by Semaglutide

**DOI:** 10.64898/2026.02.14.26346321

**Authors:** Eric B. Dammer, Shiva Afshar, Shijia Bian, Nicholas T. Seyfried, The Global Neurodegeneration Proteomics Consortium (GNPC), Allan I. Levey, Juan Fortea, Erik C.B. Johnson

## Abstract

Individuals who carry two copies of the apolipoprotein E ε4 (*APOE*ε4) allele are at high risk of developing Alzheimer’s disease (AD), yet the effects of *APOE* ε4 homozygosity on biological pathways related to AD over the lifespan are unknown. Here we analyzed the plasma proteomes of *APOE*ε4/ε4 individuals with and without AD-related cognitive impairment (*n*=413) and compared them to the proteomes of cognitively unimpaired individuals with *APOE* ε3/ε3 genotype (*n*=2764) from ages 20 to 90. Multiple biological pathways were altered in young adulthood in ε4 homozygotes including metabolism and glucagon-like peptide 1/insulin growth factor (GLP-1/IGF), mitochondrial, microtubule, proteostasis, and synaptic pathways. Semaglutide—a GLP-1 receptor agonist—demonstrated reversal effects on metabolic and synaptic pathway alterations in ε4 homozygotes at preclinical and clinical AD stages. Targeting metabolic and other pathways for therapeutic intervention in ε4/ε4 individuals by at least age 50 will likely be the most effective approach to decrease risk for AD in this special population.

**One Sentence Summary:** Dammer *et al*. characterize proteomic changes in *APO*E ε4 homozygotes from ages 20 to 90 and identify disease pathways potentially treatable with GLP-1 receptor agonists.

## INTRODUCTION

The apolipoprotein E ε4 allele (*APOE* ε4) is the strongest genetic risk factor for Alzheimer’s disease (AD) at the population level, with ε3/ε4 heterozygotes having an approximately 3-fold risk and ε4/ε4 homozygotes a 12-fold risk of developing AD, with precise risk levels depending on genetic background(*1–3*). Consequently, about 50% of patients with AD carry at least one ε4 allele(*4*). *APOE* ε4 was identified as an AD risk factor 30 years ago(*5–7*), and subsequent studies have illustrated its effect on multiple biological pathways that may influence risk for AD, including a recent large-scale plasma proteomics study that identified an inflammatory signature in ε4 carriers that may impart risk for AD(*8*). *APOE*ε4 homozygotes represent a special population of people at risk for AD given their very high lifetime risk for developing disease symptoms and the relatively low population frequency (∼2-3%) of the genotype. In the National Alzheimer’s Coordinating Center (NACC) autopsy dataset and in other cohorts with biomarker data, ε4 homozygotes exhibit near complete penetrance for AD neuropathologic change(*9*). Furthermore, ε4 homozygotes develop cognitive symptoms at approximately age 65, with variance in the age of symptom onset similar to individuals who carry autosomal dominant mutations in the amyloid precursor protein (*APP*), presenilin 1 (*PSEN1*), and presenilin 2 (*PSEN2*) genes, as well as individuals who develop AD due to Down syndrome (DSAD)(*9–11*). Because of the high lifetime risk and relative predictability of symptom onset, ε4 homozygotes have been suggested to represent a distinct genetic form of AD(*9*).

Although the effects of *APOE* ε4 on the plasma proteome have been explored in recent studies(*8, 12–14*), the temporal evolution of these changes remains unclear, particularly in ε4 homozygotes. Knowledge on the temporal progression of disease-related changes in ε4 homozygotes is important when considering the potential timing of pharmacologic or non-pharmacologic interventions to reduce risk for AD in this high-risk population. In prior studies we have used proteomic analysis of cerebrospinal fluid (CSF) to identify the biological pathways affected over the course of AD in individuals with autosomal dominantly inherited AD (ADAD) as well as in individuals with DSAD, and the temporal progression of pathological changes in these genetic forms of AD(*15, 16*). In these studies, we used cross-sectional data to estimate longitudinal changes based on the relative predictability of symptom onset in genetic AD, using the estimated year of onset (EYO) construct to place cross-sectional measurements in a longitudinal framework. In the current study, we apply the same conceptual framework to ε4 homozygotes to characterize the temporal progression of biological pathway changes starting in early adulthood in ε4/ε4 individuals compared to ε3/ε3 individuals with neutral risk for AD. We also investigate the potential for semaglutide—a glucagon-like peptide 1 receptor agonist (GLP-1RA)—to modulate these biological pathway differences at early-life, preclinical AD, and clinical AD time points.

## RESULTS

### The GNPC dataset

We used plasma proteomic data from the Global Neurodegeneration Proteomics Consortium (GNPC) harmonized dataset v1.3 for our analysis(*17*). The dataset included 19 cohorts with plasma proteomic data generated on the SomaLogic SomaScan platform, providing a total of 7,289 protein measurements encompassing 6,391 unique protein gene symbols across 17,484 individuals and 22,392 samples. We applied a quality control pipeline previously used to analyze multiple SomaScan plasma proteomic datasets from different centers to the GNPC dataset(*18*). Because of the large number of independent subjects in the dataset, we were able to develop a machine learning model to impute *APOE* genotypes with a high degree of accuracy (94% after 100-fold cross validation on hold-out test data) for those individuals who did not have an *APOE* genotype listed in the dataset (**Supplementary Tables 1 and 2**). The features driving *APOE*ε4 prediction had strong overlap with proteins strongly associated with *APOE*ε4 identified in prior studies(*8, 12, 18*). After data harmonization and quality control **(Supplementary Figure 1A-C)**, analysis of proteins altered by *APOE* ε4 genotype, age, sex, and AD were highly consistent with expected results and previous work (**Supplementary Figure 1C, D** and **Supplementary Figure 2**)(*18*), indicating successful data harmonization and quality control procedures.

From the cleaned harmonized GNPC dataset we selected *APOE* ε4 homozygotes who were either cognitively unimpaired or had mild cognitive impairment or AD, excluding non-AD neurodegenerative diseases. For the comparison group, we selected *APOE* ε3 homozygotes who were cognitively unimpaired and did not carry a neurodegenerative disease diagnosis given our goal to understand early changes in ε4 homozygotes. The selection procedure yielded *n*=413 *APOE* ε4/ε4 and *n*=2764 *APOE* ε3/ε3 cases spanning ages 20 to 90. Individuals with imputed genotypes represented a minority of each group (ε4/ε4 *n*=50, 12.1%; ε3/ε3 *n*=658, 23.8%) (**Supplementary Table 3**). We used age 65.6 as EYO 0 based on the average age of symptom onset in ε4 homozygotes as reported in the National Alzheimer’s Coordinating Center (NACC) database(*9*), and therefore the data included measurements from EYO –46 to +25 (**Supplementary Figure 3).**

*Temporal Progression of Protein Alterations in* ε4 *Homozygotes*

We applied a modeling approach previously used in ADAD and DSAD to estimate protein levels in ε4/ε4 and ε3/ε3 groups and differences in their levels across the EYO continuum(*15, 16*). For individuals who had more than one sample collection, we selected the earliest collected sample for analysis given our goal to understand early protein changes in ε4 homozygotes. An example of the model output for two proteins—tubulin specific chaperone A (TBCA) and synaptosomal-associated protein 23 (SNAP23)—is shown in **Figure 1**. TBCA was identified as an ε4-associated protein in our previous study(*18*). TBCA was decreased throughout life in ε4 homozygotes compared to ε3 homozygotes, whereas SNAP23 was increased in ε4 homozygotes beginning at EYO –22 (or age 43.6). We identified a total of *n*=878 proteins that were different in ε4 homozygotes compared to ε3 homozygotes at any timepoint (**Figure 2**, **Supplementary Figure 4**, **Supplementary Table 4**). A separate analysis using an ε3/ε3 control group that also contained MCI and AD diagnoses yielded similar overall results (**Supplementary Figure 5**), supporting the unimpaired ε3/ε3 comparator group as a valid approach to identify early ε4/ε4- and disease-related changes. There were more proteins increased (*n*=489) *versus* decreased (*n*=369) in ε4 homozygotes (with *n*=20 displaying mixed direction of change), but proteins decreased in ε4/ε4 tended to be decreased earlier in life than those that were increased, and more proteins were constitutively decreased from age 20 than increased. We performed a sensitivity analysis to determine the effect of including the minority of individuals with imputed genotypes on our results (**Supplementary Figure 6**). Inclusion of imputed samples increased power to detect protein abundance differences in ε4 homozygotes at early EYO time points and reduced effect size estimates at later EYO time points, leading to an overall fewer number of proteins considered significantly different at any EYO in ε4 homozygotes. However, the overall pattern of temporal changes was similar with or without imputation.

**Figure 1.**
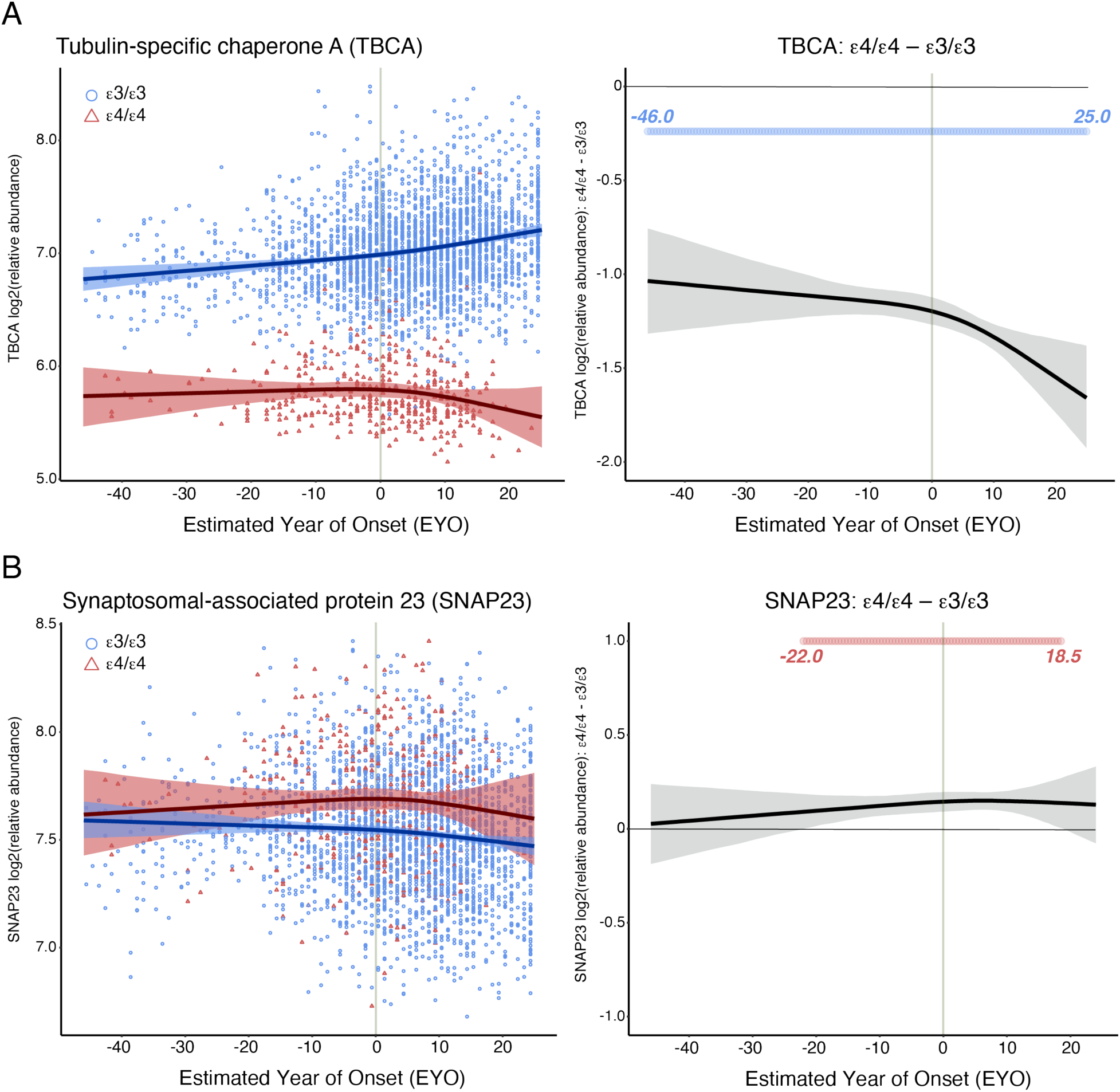
Modeling Individual Protein Levels across EYO in *APOE* ε4 Homozygotes. A total of *n*=413 *APOE* ε4/ε4 and *n*=2764 *APOE* ε3/ε3 cases were used to model protein levels across estimated year of onset (EYO). (A) Protein levels for tubulin-specific chaperone A (TBCA) in ε3/ε3 (blue) versus ε4/ε4 (red) (left). (Right) Difference between ε3/ε3 and ε4/ε4. The blue circles indicate periods of significantly lower levels of TBCA in ε4/ε4 individuals. (B) Protein levels for synaptosomal-associated protein 23 (SNAP23) (left). (Right) Difference between ε3/ε3 and ε4/ε4. The red circles indicate periods of significantly higher levels of SNAP23 in ε4/ε4 individuals. Shaded areas indicate the 99 percent credible interval.

**Figure 2.**
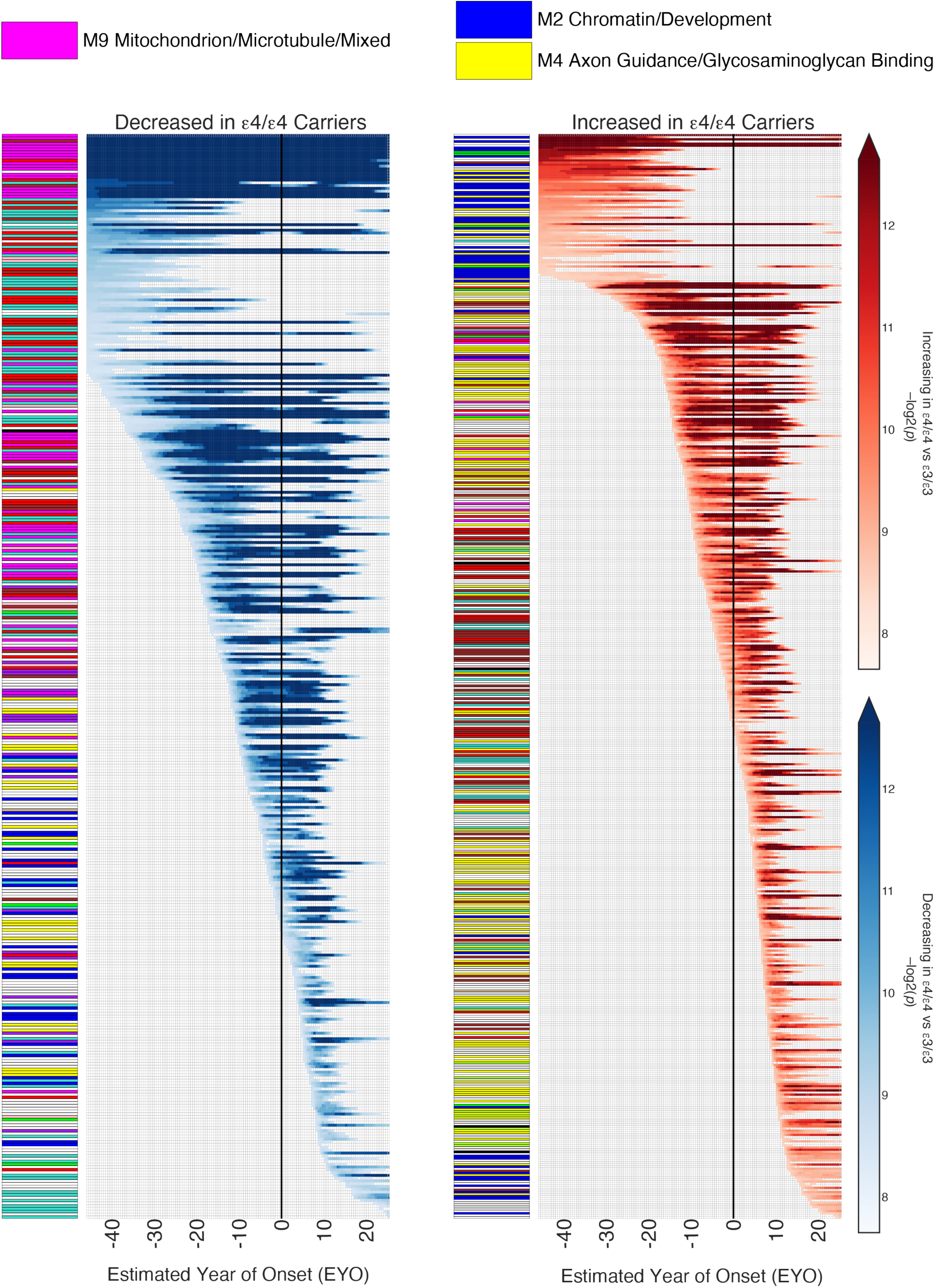
Proteins Altered in *APOE* ε4 Homozygotes by EYO. A total of *n*=878 proteins were different in ε4 homozygotes at any EYO as assessed in 0.5 EYO increments. (Left) Proteins decreased in ε4 homozygotes. (Right) Proteins increased in ε4 homozygotes. Heat is proportional to degree of statistical significance. The vertical black line indicates EYO 0, or age 65.6. Each protein was annotated with the plasma protein co-expression network module in which it resides. Network module colors and annotations for all modules are provided in **Supplementary** Figure 7 and **Supplementary Tables 5 and 6**.

To gain insight into the biological pathways associated with the altered proteins in ε4 homozygotes, we generated a protein co-expression network on the entire GNPC dataset (*n*=22,392 samples) which yielded 12 protein co-expression modules associated with different biological pathways (**Supplementary Figure 7A, Supplementary Tables 5 and 6**, **Supplementary Information)**, and mapped each protein to its cognate module (**Figure 2**). Many proteins that were either constitutively decreased or decreased early in ε4 homozygotes mapped to the M9 Mitochondrion/Microtubule/Mixed module. The M9 module contained many ε4-related proteins (**Supplementary Figure 7B, Supplementary Table 5**), and its eigenprotein was decreased in ε4 homozygotes from age 20 (**Supplementary Figure 7C, D**) consistent with the individual protein mapping results. Many proteins that were increased in ε4 homozygotes mapped to the M2 Chromatin/Development and M4 Axon guidance/Glycosaminoglycan binding modules, with an apparent biphasic response in M4 represented by an early and late period of increase. Some of the proteins that mapped to these modules were sex-dependent, with a higher level of M4 proteins observed early in females and higher levels in males closer to symptom onset. This pattern appeared to be inverted for some M9 proteins (**Supplementary Figure 8**, **Supplementary Table 7**). A model to test *APOE* ε4/ε4 interaction with sex by EYO identified 61 proteins with a significant interaction at some EYO (**Supplementary Table 7**). Overall, these results suggest that sex may influence ε4/ε4-related proteins even early in life.

### Overlap with AD Endophenotypes

We leveraged our prior analysis on plasma proteins associated with different AD endophenotypes to investigate the relationship between ε4/ε4-associated proteins and these endophenotypes(*18*). In an analysis of the Religious Orders Study and Rush Memory and Aging Project (ROSMAP) endophenotypes, we found ε4/ε4-associated proteins primarily enriched at a nominal level in plasma proteins associated with neurofibrillary tangles and cerebral atherosclerosis prior to symptom onset (**Supplementary Figure 9, Supplementary Table 8**). ε4/ε4-associated proteins were also enriched in proteins associated with cerebral β-amyloidosis at all time points in a meta-analysis across four cohorts(*18*), consistent with the known association between early AD neuropathologic change in ε4 homozygotes and the influence of the ε4 genotype on amyloid-β (Aβ) deposition(*19*).

### Temporal Biological Pathway Changes in ε4 Homozygotes

To obtain more granular resolution of the pathways associated with proteins altered in ε4 homozygotes than what could be provided by protein co-expression network analysis, we performed a sliding window enrichment analysis across the EYO continuum of ε4/ε4-associated proteins for individual ontology categories from the Gene Ontology (GO) and Reactome ontology sets. We clustered enriched ontologies from similar pathways that were identified across these two ontology sets (**Figure 3, Supplementary Table 9**). To better understand directionality of ε4/ε4-related effects, we then performed the same analysis separately for increased versus decreased proteins in ε4 homozygotes, with the understanding that not all proteins within a pathway will have the same direction of change if a pathway is influenced by the ε4 allele (**Figure 4**, **Supplementary Figure 10, Supplementary Tables 10 and 11**). We identified 18 general categories of pathway changes related to the ε4/ε4 genotype. Overall metabolism, including small molecule and NAD^+^ metabolism, was strongly decreased throughout life in ε4 homozygotes. GLP-1 pathways were altered both early and late, whereas the insulin growth factor binding proteins (IGFBPs) were predominantly altered around the time of symptom onset. As expected, lipoprotein pathways were altered throughout life. RNA processing was altered early, but the effect diminished with age. Eye and retinol metabolism pathways were decreased throughout life. Heparan sulfate metabolism pathways were altered prior to symptom onset, but other glycosaminoglycan and extracellular matrix pathways were strongly elevated after symptom onset. Proteostasis pathways, including ubiquitin processing and neddylation, were decreased early, with notable elevations in lysosomal and vacuolar lumen pathways also at early timepoints. Synaptic and neuronal pathways showed a biphasic response, with pre- and post-synaptic pathways elevated early and neuronal developmental pathways altered late. Immune pathways altered early included negative regulation of interferon-alpha production and complement. Overall protein translation was also decreased prior to symptom onset in ε4 homozygotes. In summary, multiple pathways were affected early in ε4 homozygotes, with prominent alterations in metabolism, proteostasis, synaptic/neuronal, retinol, and protein translation pathways, illustrating the pleotropic effects of *APOE*ε4 homozygosity.

**Figure 3.**
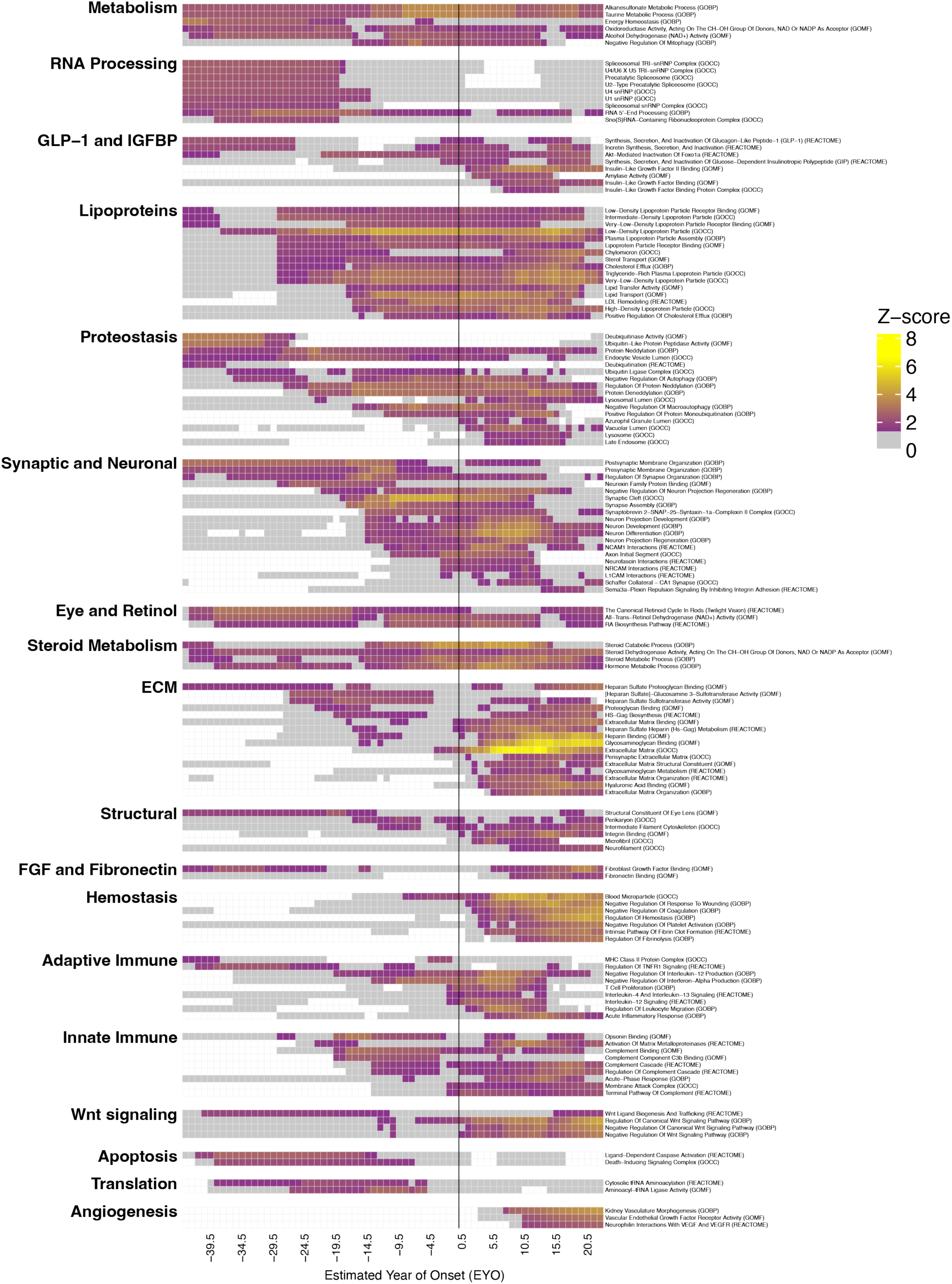
Biological Pathways Enriched in ε4/ε4-associated Proteins. The top pathways enriched in ε4/ε4-associated proteins from the GO biological process (GOBP), molecular function (GOMF), cellular component (GOCC), Reactome ontology sets were grouped by category, with 0.5 EYO periods of significant enrichment indicated by heat color and near-significant enrichment indicated by gray. The vertical black line indicates EYO 0, or age 65.6. Information on pathways and periods of enrichment is also provided in **Supplementary Table 9**.

**Figure 4.**
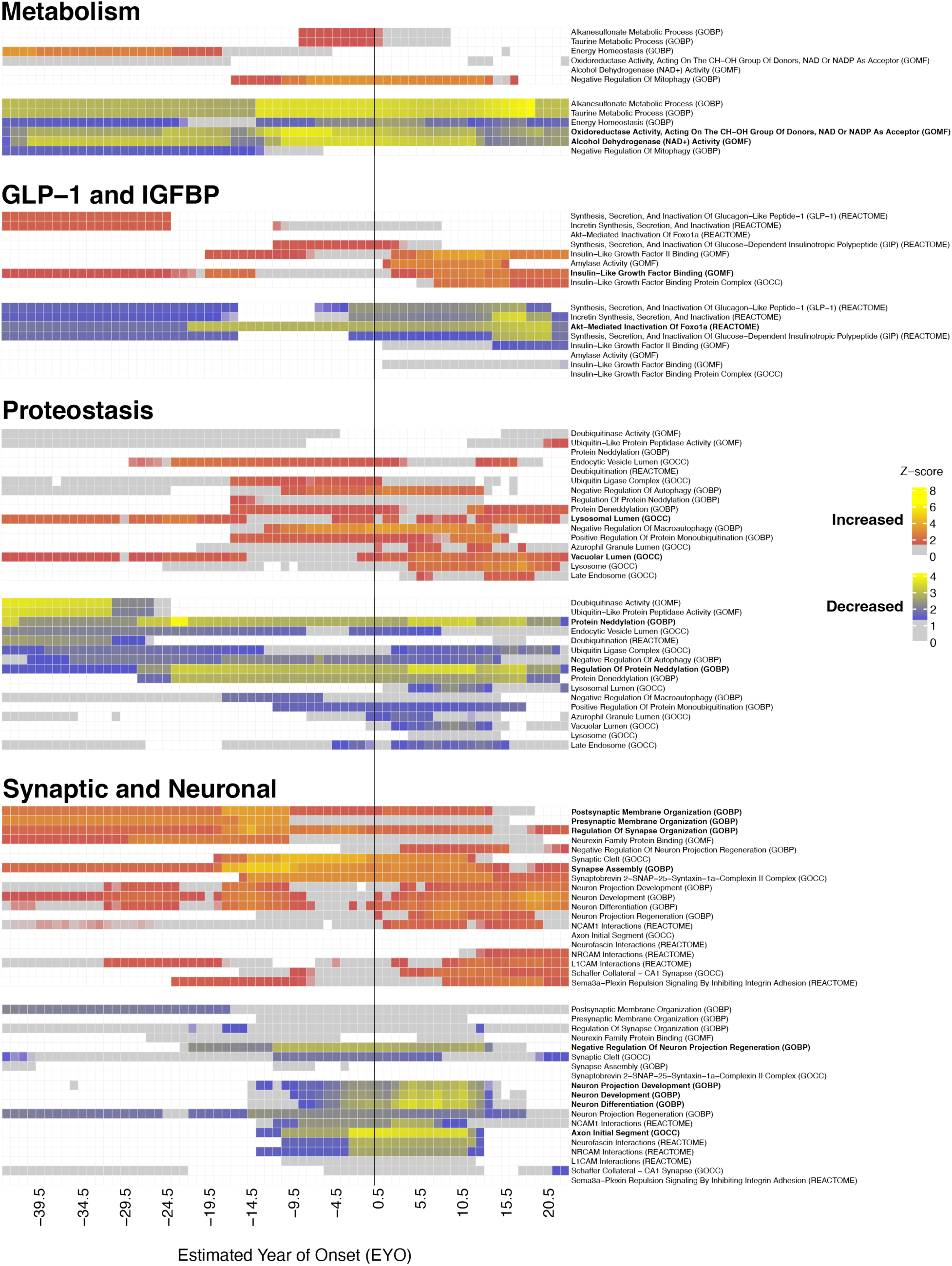
Selected Biological Pathways Enriched in Proteins Increased or Decreased in ε4 Homozygotes. Pathways enriched in proteins increased (red) or decreased (blue) in ε4 homozygotes from the GO biological process (GOBP), molecular function (GOMF), cellular component (GOCC), and Reactome ontology sets were grouped by category, with 0.5 EYO periods of significant enrichment using a 5-year sliding window indicated by heat color and near-significant enrichment indicated by gray. A selection of four AD-related pathways is shown here, with the full set of 18 ontology groups provided in **Supplementary** Figure 10 and **Supplementary Tables 10 and 11**. The vertical black line indicates EYO 0, or age 65.6.

### Overlap with AD Risk Proteins

Plasma proteomics performed in large longitudinal cohorts has identified proteins associated with risk of incident cognitive impairment, which can occur decades later. We examined overlap of proteins affected by *APOE* ε4 homozygosity prior to age 50 (when Aβ begins to deposit in ε4 homozygotes(*9*)) with proteins associated with risk of incident cognitive impairment identified in the Atherosclerosis Risk in Communities (ARIC) study(*20–22*), the Age, Gene/Environment Susceptibility (AGES)-Reykjavik study(*12, 23*), the Whitehall II study(*24*), the UK Biobank(*25*), and the Religious Orders Study and Rush Memory and Aging Project (ROSMAP)(*18, 26*) (**Figure 5**), with the goal of identifying proteins altered early in ε4 homozygotes that may be related to dementia risk. Proteins that were altered in ε4 homozygotes prior to age 50 and were identified as risk factors for dementia in at least two other cohorts included IGFBP2, growth/differentiation factor 15 (GDF15), metalloproteinase inhibitor 4 (TIMP4), and the extracellular matrix SPARC-related modular calcium-binding protein 1 (SMOC1). Most of the associations were in the same direction between ε4 homozygotes and the risk identified in these studies. These findings were consistent with proteins altered in AD in the GNPC (**Supplementary Figure 2A**). They were also consistent across multiple SomaScan assays of the same protein where available—in the case of IGFBP2, across four separate assays (**Supplementary Figure 11A**)—and across proteomic platform in the case of the UK Biobank, which used the Olink platform. We also tested for overlap with proteins that comprise the dementia SomaSignal® test (dSST) panel that can be used to predict risk of all-cause dementia (**Supplementary Figure 11B**)(*27*). Five out of 25 proteins in the dSST overlapped with proteins altered in ε4 homozygotes, four of which—TBCA, ankyrin repeat and SOCS box protein 9 (ASB9), neurofilament light polypeptide (NEFL), and protein S100-A13 (S100A13)—were altered prior to age 50, and one of which, neuronal pentraxin receptor (NPTXR), was altered after age 50 but prior to age 65. TBCA, NEFL, and S100A13 have previously been associated with *APOE* ε4(*18*), and NPTXR has been associated with cognitive decline(*15, 28*). In summary, we identified protein alterations in ε4 homozygotes prior to age 50 that have been associated with risk for incident cognitive impairment, with elevation in IGFBP2, GDF15, TIMP4, and SMOC1 identified as highly robust findings, highlighting the importance of biological pathways associated with these proteins—energy metabolism (IGFBP2/GDF15), stress response/immune (GDF15/TIMP4), vascular (TIMP4/SMOC1), and extracellular matrix/heparin binding (SMOC1)—for imparting risk of cognitive decline in *APOE* ε4 homozygotes.

**Figure 5.**
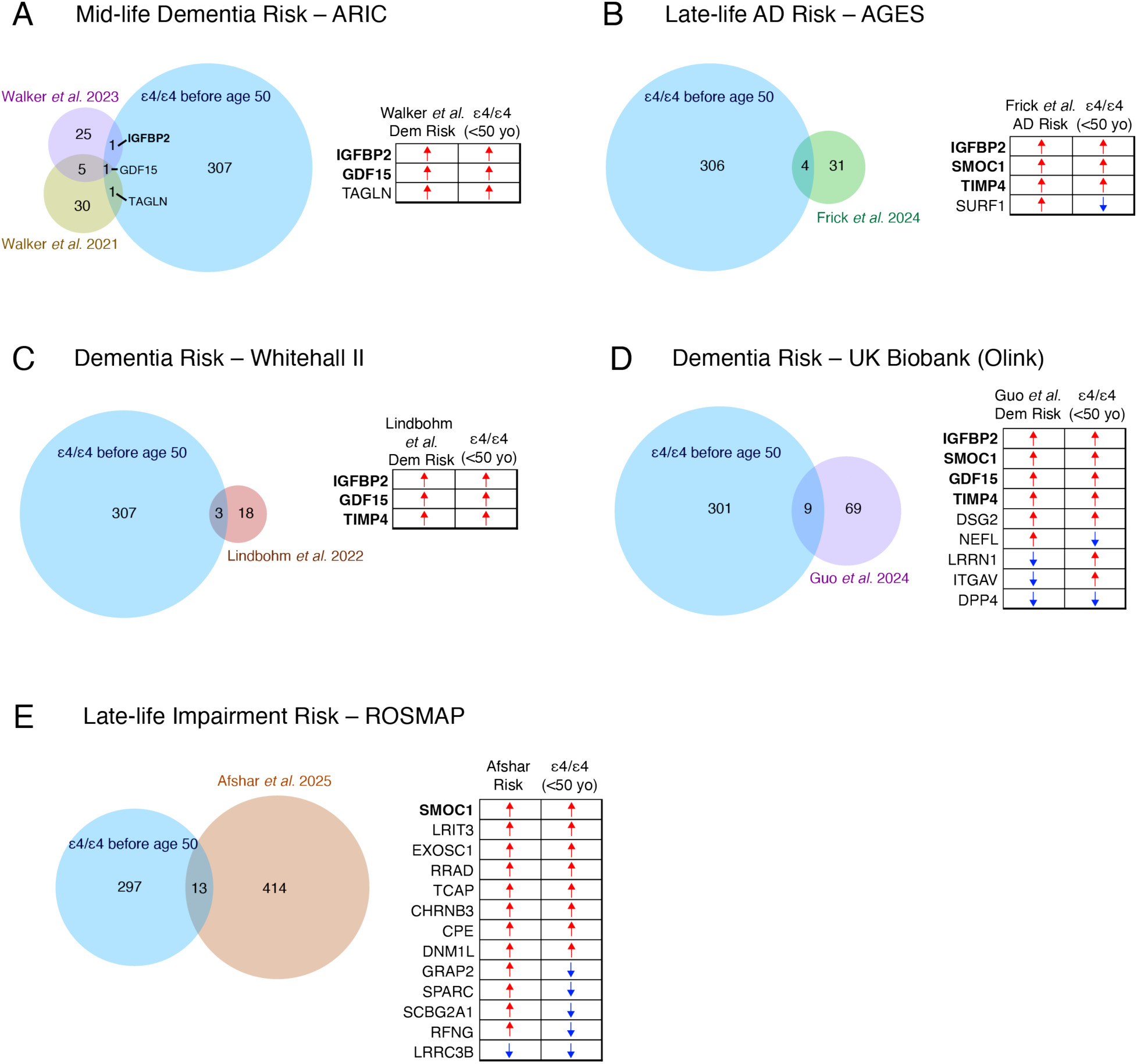
Proteins Altered Early in ε4 Homozygotes Associated with Risk of Incident Cognitive Impairment. Proteins altered in ε4 homozygotes prior to age 50 were assessed for overlap with proteins associated with mid-life dementia risk in the ARIC cohort (Walker *et al*.(*20, 21*)) (A), late-life AD risk in the AGES-Reykjavik cohort (Frick *et a*l.(*12*)) (B), dementia risk in the Whitehall II cohort (Lindbohm *et al*.(*24*)) (C), dementia risk in the UK Biobank (Guo *et al*.(*25*)) (D), and risk of late-life cognitive impairment in the ROSMAP cohort (Afshar *et al*.(*18*)) (E). Proteins in the UK Biobank were measured using the Olink proteomic platform. Risk proteins in ROSMAP were at nominal statistical significance, without correction for multiple comparisons. Protein overlaps in common in at least two other cohorts are highlighted in bold. Direction of risk is provided by colored arrows (red, higher protein levels lead to increased risk; blue, lower protein levels lead to increased risk). No overlaps between the referenced protein sets and this study were significant.

### Effects of Semaglutide on ε4/ε4 Risk Proteins and Altered Biological Pathways

Semaglutide is a GLP-1RA currently used for the treatment of type II diabetes and obesity. However, the effects of semaglutide extend beyond blood glucose control and appetite suppression and include beneficial effects on systemic inflammatory and metabolic pathways, among others, leading to lower risk of adverse cardiovascular events, renal impairment, and progression of metabolic dysfunction-associated steatohepatitis (MASH)(*29*). We investigated the potential for semaglutide to modify biological pathway alterations in ε4 homozygotes by leveraging plasma proteomic data from the STEP1 clinical trial of semaglutide treatment in overweight or obese individuals without diabetes (*n*=1,311 participants with serum SomaScan proteomic data before and after 68 weeks of subcutaneous semaglutide, average age 47.5), using proteins for our analysis that were significantly altered by semaglutide independent of changes in body weight or blood glucose(*30*). We found a significant overlap between proteins affected by ε4/ε4 and semaglutide (44 out of 153 proteins affected by semaglutide, *p* = 1.6e^−8^), which was primarily driven by proteins affected by ε4/ε4 after age 50 (overlap before age 50, 5 out of 153, *p* = 0.81; overlap after age 50, 44 out of 153, *p* = 8.7e^−10^). Proteins influenced by semaglutide independent of changes in body weight and blood glucose were used to calculate the potential effect of semaglutide treatment on biological pathways altered in ε4 homozygotes at three epochs—early-life (EYO –46 to –15.5), preclinical AD (EYO –15 to –0.5), and AD stages (EYO 0 to 25)—through a semaglutide connectivity score whereby a positive score indicates a semaglutide effect in the same direction as the ε4/ε4 effect, and a negative score indicates a semaglutide effect in the opposite direction of the ε4/ε4 effect (**Figure 6A**, **Supplementary Figures 12 and 13**, **Supplementary Table 12**). We observed overall consistent reversal effects in metabolism, GLP-1/IGFBP, synaptic/neuronal, and eye/retinol pathways in both preclinical and clinical AD stages, although no effects survived FDR correction applied across all ontologies and EYO epochs. Reversal effects in adaptive/innate immune pathways and protein translation were most consistently observed at the clinical AD stage. RNA processing showed concordant direction of change for semaglutide and ε4/ε4 at all three epochs. Hemostasis and both adaptive and innate immune pathways showed concordant effects in the preclinical AD stage. A sensitivity analysis conducted on the ε4/ε4 effect size threshold to include for connectivity analysis showed that reversal effects in synaptic/neuronal, metabolism, and GLP-1/IGFBP pathways were generally stronger at lower effect size thresholds, whereas reversal effects in adaptive and innate immune pathways were generally stronger at higher effect size thresholds, particularly in the clinical AD stage (**Supplementary Figures 12 and 13**). These results suggest that many proteins in synaptic and metabolic pathways affected in a reversal direction by semaglutide have relatively small fold-changes in ε4/ε4 individuals, whereas the fold-changes in immune pathways affected by semaglutide are relatively larger in ε4/ε4 individuals, especially in the clinical AD stage, potentially reflecting immune system changes related to ongoing neurodegeneration. Overall, these analyses highlight the time-dependent effects of semaglutide on certain pathways altered in ε4 homozygotes.

**Figure 6.**
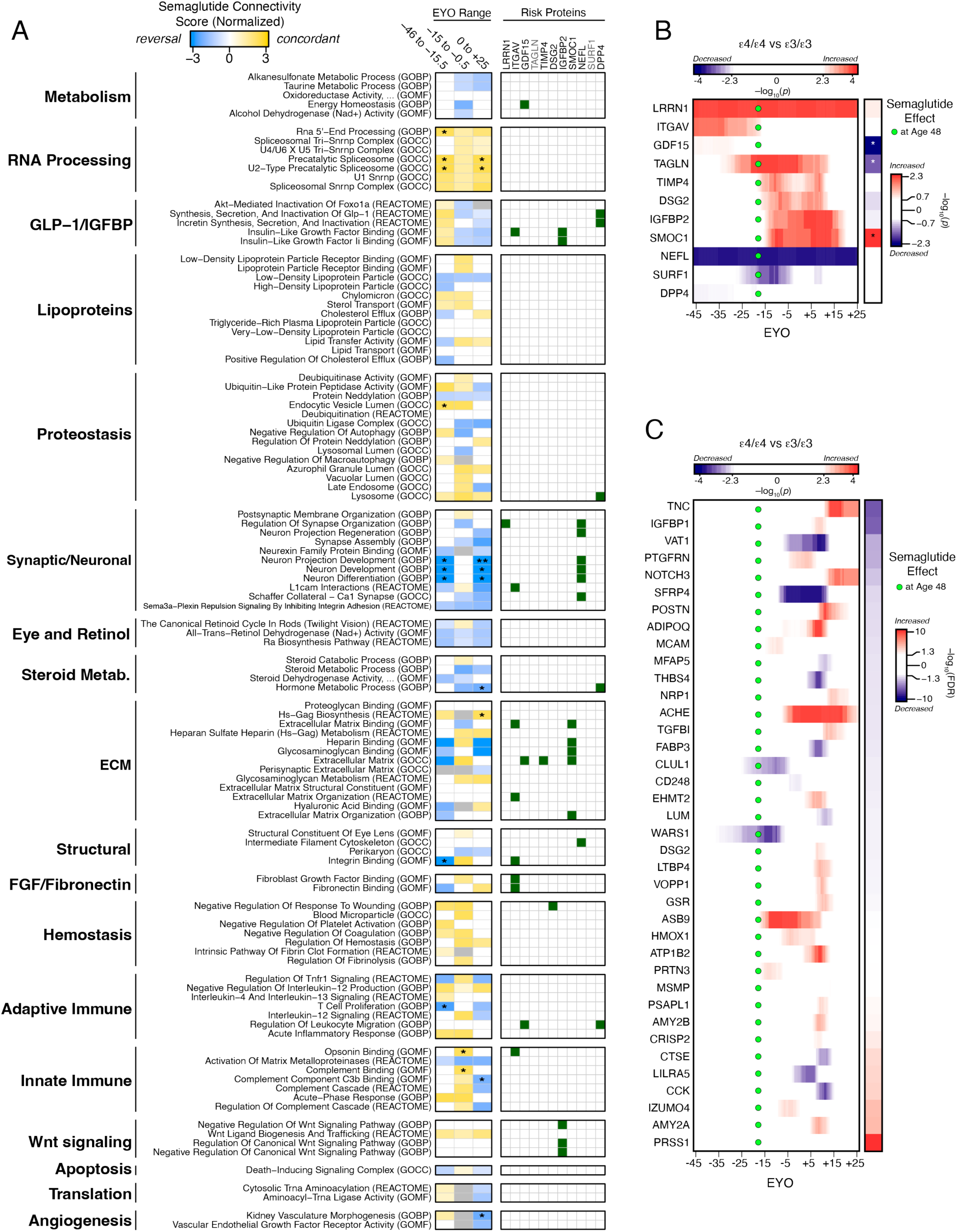
Predicted Effect of Semaglutide on Pathways and Proteins Altered in ε4 Homozygotes. Proteins influenced by semaglutide in the STEP1 trial as described by Maretty *et al*.(*30*) after adjustment for weight and blood glucose changes were used to assess the potential effect of semaglutide treatment on biological pathways altered in ε4 homozygotes from the GO molecular function (GOMF), biological process (GOBP), cellular component (GOCC), and Reactome ontology sets (A). Treatment effects were assessed in early-life (EYO –46 to –15.5), preclinical (EYO –15 to –0.5), and clinical (EYO 0 to 25) AD stages. Semaglutide connectivity score indicates the direction of predicted effect (gold, same direction as ε4/ε4; blue, opposite direction as ε4/ε4). Dementia risk proteins in the ARIC, AGES, Whitehall II, and UK Biobank studies that overlapped with proteins altered in ε4 homozygotes prior to age 50 as described in Figure 5 were mapped to the corresponding biological pathways affected by semaglutide (green squares). (B) Difference in protein levels between ε4 and ε3 homozygotes across EYO for the eleven dementia risk proteins in (A) (red, increased levels in ε4/ε4; blue, decreased levels in ε4/ε4), and the effect of semaglutide on these proteins in the STEP1 trial (red, increased with semaglutide treatment; blue, decreased with semaglutide treatment). Green circles indicate average age of participants (age 48) in the STEP1 trial. (C) Other proteins different in ε4 homozygotes and also affected by semaglutide in the STEP1 trial (*n*=38). Connectivity score in (A) is reported after permutation. *\*p* < 0.05; \*\**p* < 0.01; \*\*\**p* < 0.001 after 100,000 permutations. No score was significant after FDR applied across all ontologies and EYO epochs. Results of an ε4/ε4 effect size threshold sensitivity analysis are provided in **Supplementary Figures 12 and 13**. Threshold for ε4/ε4 differences is at the 99% credible interval (*p* < 0.005) in (B) and (C). Threshold for semaglutide effect color is at nominal *p* < 0.2 in (B) (\**p* < 0.05), and *p* < 0.05 after FDR in (C). All proteins shown in (C) were significant at *p* < 0.05 after FDR.

We also analyzed the effect of semaglutide on the eleven ε4/ε4-altered proteins that overlapped with dementia risk proteins identified in the ARIC, AGES, Whitehall II, and UK Biobank studies, excluding ROSMAP due to the nominal effects observed in this study (**Figure 6B**). Some of these proteins such as IGFBP2, integrin alpha-V (ITGAV), and dipeptidyl peptidase 4 (DPP4) mapped to pathways affected by semaglutide (**Figure 6A**). Among these proteins, the strongest effect of semaglutide treatment was observed with GDF15, transgelin (TAGLN), and SMOC1, where a reversal effect was observed for GDF15 and TAGLN and a concordant effect was observed for SMOC1. A non-significant but consistent reversal effect was observed for all four assays of IGFBP2 at the time this protein becomes altered in ε4 homozygotes, at approximately age 50 (**Supplementary Figure 11C**). There were 38 other proteins significantly altered by both ε4/ε4 and semaglutide that were not identified as dementia risk proteins in the previous studies (**Figure 6C**). Most of these proteins were altered in the symptomatic stage of disease including tenascin (TNC), acetylcholinesterase (ACHE), and IGFBP1, which showed reversal effects with semaglutide. Exceptions included clusterin-like protein 1 (CLUL1) expressed in the retina(*31*), tryptophan-tRNA ligase (WARS1) involved in protein translation and vascular biology(*32*), and ASB9 involved in ubiquitination(*33*) that were altered closer to the time of the semaglutide effect observed in the clinical trial. In summary, we identified potentially beneficial effects of semaglutide on metabolic, GLP-1/IGFBP, and synaptic/neuronal proteins and pathways altered in ε4 homozygotes prior to and after symptom onset, suggesting GLP-1RAs such as semaglutide may have therapeutic benefit in ε4 homozygotes in preclinical and clinical AD stages.

## DISCUSSION

In this study we analyzed the plasma proteome signature of *APOE* ε4 homozygotes from ages 20 to 90 and characterized the evolution of biological pathway changes from early adulthood to later in life when most ε4 homozygotes develop AD. Our results illustrate that many AD-related pathways altered near the time of symptom onset in ε4 homozygotes, including glucose and oxidative metabolism, proteostasis, synapse/neuronal, and immune pathways, are also altered prior to age 50, and that some of these pathways could be targeted for therapeutic modulation with GLP-1RAs to prevent, delay, or treat AD in ε4/ε4 individuals. These findings suggest that ε4 homozygosity imparts multiple detrimental biological effects in early life that influence risk for AD, and highlight the likely importance of early multi-modal therapeutic intervention with pharmacologic and non-pharmacologic strategies to prevent AD in this special population. We discuss some of the key pathways influenced by ε4 homozygosity below.

### Metabolism

*APOE* ε4 has been shown to affect metabolism and mitochondrial function in multiple brain cell types, including neurons, astrocytes, and microglia(*34–36*). The mechanism by which ε4 affects metabolism in brain cell types may be through lipid transport effects, mitophagy(*37, 38*), changes in respiratory complex subunits leading to decreased oxidized nicotinamide adenine dinucleotide (NAD^+^) and increased reactive oxygen species (ROS)(*39*), and direct mitochondrial toxicity from proteolytic fragments of ε4(*34, 40*). Although glucose is the brain’s primary energy source under normal dietary conditions, the role of systems such as the astrocyte-neuron lactate shuttle in which astrocytes provide metabolic substrates to neurons for mitochondrial metabolism are known to be critical for proper neuronal function(*41*). A signature endophenotype of AD is reduced glucose metabolism in AD-susceptible brain regions, and the same endophenotype with less severity has been observed in ε4 homozygotes in young adulthood(*42, 43*). Here we have observed a metabolic signature in ε4 homozygotes in plasma in young adulthood which is likely related to systemic differences in bioenergetics in homozygotes, with the organ that has the most bioenergetic demand—the brain—perhaps most susceptible over time to such baseline bioenergetic differences. We have previously observed proteomic changes in mitochondrial and oxidative stress pathways in cerebrospinal fluid related to *APOE*ε4(*44*). Here we also observe changes in GLP-1 and IGFBP pathways in ε4 homozygotes, including in forkhead box protein O1 (FOXO1), which is a master regulator of insulin signaling that regulates metabolic homeostasis in response to oxidative stress(*45*). The IGFBPs important for insulin growth factor signaling were increased around symptom onset except for IGFBP2, which was increased at least 15 years prior to symptom onset and has been identified as a marker of risk for incident cognitive decline in multiple studies. Another risk marker altered in ε4 homozygotes, SURF1, is a component of the protein complex that regulates mitochondrial cytochrome c oxidase assembly, mutations in which cause the mitochondrial disease Leigh syndrome which is best treated with a ketogenic diet(*46*). Related to this point, recent studies have also suggested that the Mediterranean diet—which reduces risk for AD(*47, 48*)—has the largest effect for AD risk reduction in ε4 homozygotes(*49*), potentially through lipid, mitochondrial metabolism and oxidative stress pathways. Semaglutide has been shown to improve mitochondrial fitness and reduce reactive oxygen species(*50*), and demonstrated a reversal effect on the metabolic phenotype in ε4 homozygotes starting at least 15 years prior to symptom onset and extending into the clinical phase of AD. Given the metabolic differences in ε4 homozygotes and the potential beneficial effects of GLP-1RAs in reducing risk for dementia observed in prior studies(*51*), and the potential beneficial effect specifically for ε4 homozygotes observed in this study, a treatment and/or prevention trial of GLP-1RAs in ε4 homozygotes to treat or reduce risk for AD should be given serious consideration. Overall, our findings suggest early dietary and pharmaceutical intervention to support healthy brain metabolic function may be particularly effective in ε4 homozygotes to reduce risk for AD.

### Proteostasis

Protein quality control is particularly important in the central nervous system where most cells are post-mitotic and highly metabolically active. In plasma, we observed alterations in proteostasis pathways in ε4 homozygotes from early adulthood, including ubiquitination, neddylation, autophagy, lysosomes and endosomes, suggesting proteostasis is systemically altered in ε4 homozygotes throughout life. In general, proteins in these pathways were lower in ε4 homozygotes with the exception of lysosomal and vacuolar lumen proteins, which were elevated in plasma, perhaps reflecting dysfunction in these structures leading to extracellular protein secretion. It is also possible that proteostasis in ε4 homozygotes is shifted across the three primary cellular systems involved in proteostasis: ATP-dependent chaperone-assisted refolding, proteasome-mediated degradation, and lysosome-mediated degradation, as has been observed in AD brain tissue(*52*). We have previously observed differences in neddylation—a process similar to ubiquitination in which the protein NEDD8 is conjugated to protein substrates and plays a key role in cell cycle, immune function, and protein degradation(*53, 54*)—in CSF in *APOE* ε4 carriers(*44*), likely reflecting the effects of ε4 on this pathway in the brain. In ROSMAP we observed an association between ε4/ε4-related proteins and proteins previously identified to be associated with tau NFTs(*18*), including TBCA and kinetochore protein SPC25 (SPC25) involved in microtubule function, potentially linking NFT formation to ε4 through its effect on microtubules. In addition to earlier accumulation of Aβ plaques and tau neurofibrillary tangles in ε4 carriers, ε4 is also associated with the development of other protein aggregates in brain such as Lewy bodies and TAR DNA-binding protein 43 (TDP-43) inclusions(*55, 56*), suggesting that decreased overall protein quality control as measured in the periphery is manifested in the brain by formation of various proteinopathies. We also observed differences in RNA-binding proteins required for the spliceosome complex, such as the small nuclear ribonucleoproteins (snRNPs), in ε4/ε4 plasma at early time points. The functional consequences of this difference could be reflected by altered protein splicing, but could also predispose to snRNP aggregation in the brain, which is an early neuropathological feature in AD(*57–59*). Semaglutide may augment the observed changes in RNA processing, and to the extent the changes in RNA-binding proteins observed in ε4 homozygotes are involved in AD pathophysiology, clinical trials with semaglutide should ideally monitor changes in this pathway. Nevertheless, augmenting autophagy and endosome/lysosome pathways to support proteostasis by administration of small molecule therapeutics such as rapamycin or metformin, perhaps in combination with a GLP-1RA such as semaglutide to target metabolic dysfunction, may be particularly effective in reducing AD risk in ε4 homozygotes(*60*).

### Synapses and Neurons

Proteins associated with both pre-synaptic and post-synaptic terminals were altered in plasma in ε4 homozygotes in early adulthood, well before the development of classic AD neuropathology. In neuronal iPSC culture, ε4 leads to an increased number of synapses(*61*). However, ε4 carriers have been found to have smaller medial temporal lobe structures in young adulthood by MRI(*62, 63*), and hippocampal synaptic loss prior to plaque deposition has been observed by PET imaging(*64*) that may be due in part to loss of inhibitory interneuron synaptic function(*65*). How ε4 leads to increased synaptic proteins in plasma is not clear. It is important to note that levels of some synaptic and neuronal proteins may be anticorrelated between plasma and CSF compartments(*18, 66*), and therefore inferences on brain health based on the directionality of change in plasma should be made with caution until the relationship between brain, CSF, and plasma compartments in the levels of these proteins is better understood. In general, more synaptic proteins were increased rather than decreased early, perhaps reflecting a difference in central nervous system (CNS) development in ε4 homozygotes, but near symptom onset there was a shift to decreased levels of neuronal proteins associated with developmental processes. This shift began around age 50, or about 15 years prior to symptom onset, and is likely associated with Aβ deposition and neurodegeneration(*67*). Semaglutide demonstrated an overall reversal effect in this pathway which was particularly evident in semaphorin-plexin signaling. Our findings suggest that ε4 homozygotes have altered synaptic and neuronal physiology in early adulthood that is likely attributable to the CNS and may predispose them to synaptic and neuronal dysfunction associated with AD later in life. They also suggest that GLP-1RAs may be effective in reversing, at least in part, this synaptic and neuronal vulnerability.

### Adaptive and Innate Immune

*APOE* ε4 is associated with a general immune system phenotype that is considered pro-inflammatory and is observed across multiple neurodegenerative diseases(*8*). We grouped inflammatory changes in ε4 homozygotes into adaptive and innate categories to distinguish complement from other immune changes, although it is possible that the changes observed in the adaptive category mediate their effects through the innate immune cells of the CNS—microglia. ε4 causes a pro-inflammatory microglial phenotype in dose-dependent fashion(*68*), with higher levels of tumor necrosis factor alpha (TNF-alpha), interleukin 6 (IL-6), and IL-12 that worsen neurodegeneration in AD models(*69, 70*). We observed an immune profile in ε4 homozygotes consistent with a pro-inflammatory state, with alterations in TNF, interferon alpha (IFN-alpha), and IL-12. T cell signaling was elevated around the time of symptom onset, which has been proposed to mediate cognitive decline in ε4 carriers(*14*). Similar to DSAD(*16*), we also observed changes in the complement pathway, with decreases in proteins involved in complement binding observed about 15 years prior to symptom onset perhaps leading to dysregulated complement activity(*71*). This change occurred slightly before the decrease in neuronal proteins, potentially linking altered complement activity with neuronal loss(*72*). ε4 homozygotes also have the highest risk for hemorrhagic complications from anti-amyloid immunotherapy(*73*), which is hypothesized to be caused by higher levels of complement deposition in the cerebral vasculature leading to aberrant immune activation in the vessel wall and loss of vascular integrity(*74*). Although early and more intense activation of adaptive and innate immune pathways may confer a survival benefit in early adulthood for ε4 homozygotes in high pathogen environments(*75*), repeated activation through various environmental exposures throughout life and subsequent potential low-grade chronic activation may serve as a risk factor for development of AD in homozygotes later in life. Although the effect of semaglutide on innate and adaptive immune pathways was variable and dependent on disease stage, GLP-1RAs are known to decrease levels of inflammatory mediators such as TNF-alpha, IL-6, and IL-12(*50, 76*), and may therefore have therapeutic benefit in ε4 homozygotes if administered at the appropriate time.

### Other Key Protein Markers

GDF15 was elevated prior to age 50 in ε4 homozygotes and has been associated with incident dementia risk in multiple cohorts. GDF15 is a central senescence factor(*77*) that operates at the nexus between the immune system and metabolism(*78*) and has been linked to mitochondrial dysfunction(*79*). Elevation of GDF15 suggests ε4 homozygosity may accelerate cellular senescence. TAGLN, which was identified as a risk factor in the ARIC cohort and is strongly associated with AD in GNPC, is an actin-binding protein potentially involved in senescence of vascular smooth muscle cells(*80*). Both GDF15 and TAGLN may be reduced in ε4 homozygotes by treatment with semaglutide. TIMP4 is elevated in cerebral vasculature with cerebral amyloid angiopathy (CAA) pathology and correlates with CAA severity and hemorrhage(*81*), linking the increased presence of CAA in ε4 homozygotes to this dementia risk protein years before symptom onset. DPP4, which was decreased in both the UK Biobank and in ε4 homozygotes, is a regulator of T cell activation which is a process that has been linked to cognitive impairment(*14, 18*). Finally, SMOC1 has been identified as a risk factor in multiple cohorts and was increased in ε4 homozygotes prior to age 50. SMOC1 is associated with both CAA(*82*) and cerebral amyloid deposition(*18, 83*), and is altered in CSF in ADAD and DSAD at the earliest stages of disease prior to elevations in pTau217(*15*), highlighting the importance of this extracellular matrix protein in plasma as a biomarker for AD risk. SMOC1 may be increased by semaglutide treatment in ε4 homozygotes, although the potential therapeutic consequence of SMOC1 alteration in plasma remains unknown.

### Eye and Retinol

We identified biological pathway changes in ε4 homozygotes that, while not directly associated with AD, may be important to consider for ε4-related therapeutic development. In addition to a potential survival benefit in the young conferred by ε4 homozygosity, ε4 homozygotes display approximately half the risk for developing age-related macular degeneration compared to ε3 homozygotes(*84, 85*). The mechanism driving this protective effect is not known. Here we observe that one of the strongest affected pathways in ε4 homozygotes is retinol metabolism and the retinoid cycle in rods, with decreased levels of these pathway proteins potentially providing a mechanistic link between retina health and the ε4 allele. ε4 also reduces risk of retinal damage from glaucoma(*86*), which has been proposed to be mediated by poor microglial activation in the retina in the setting of increased intraocular pressure(*87*). Semaglutide counters the ε4/ε4 effect on retinol metabolism. The beneficial effects of ε4 homozygosity on retinal health should be considered when developing therapeutics that directly target *APOE*(*88, 89*), and retinal heath would be an important outcome to monitor in a clinical trial of semaglutide in ε4 homozygotes.

### Implications for Clinical Translation

Recent clinical trials of GLP-1RAs in AD have shown negative results. The evoke and evoke+ phase 3 clinical trials of semaglutide in early-stage symptomatic AD showed no slowing of disease progression, although some beneficial effects in an exploratory analysis of CSF AD, immune, and neuronal biomarkers was observed(*90*), consistent with the predicted reversal effects we observe in synapse/neuronal and immune pathways in ε4 homozygotes in this study. A phase 2b clinical trial of liraglutide in mild to moderate AD was negative on the primary outcome measure of change in cerebral glucose metabolic rate, but did show preservation of brain volumes compared to placebo(*91*). These results stand in contrast to the potential unique benefits of GLP-1RAs for dementia risk reduction in individuals with diabetes(*51*). The proportion of ε4 homozygotes in the evoke and evoke+ trials was approximately 12%, and results were not stratified by *APOE* genotype. Similarly, the liraglutide AD trial results were not stratified by *APOE* genotype nor was *APOE* genotype distribution in the trial population reported. Post-hoc analyses of current GLP-1RA trials in AD stratified by *APOE* genotype would be informative for the findings presented in this study, particularly for the clinical phase of AD. If ε4 homozygosity represents a unique genetic form of AD, then clinical trials that sample the general AD population may not accurately reflect the potential benefits, or lack thereof, in the ε4 homozygote population. Indeed, this may be the case for anti-amyloid immunotherapies in the symptomatic phase of the disease(*92, 93*). Regardless of *APOE* genotype, given the current data on GLP-1RAs in AD and the preclinical changes observed in GLP-1 pathways and IGFBP2 in this study and other studies, as well as the predicted reversal effects of semaglutide treatment on these pathways, it is likely that the largest benefit of GLP-1RAs for AD will be observed in a preventive treatment paradigm, and preclinical and clinical studies that focus on the asymptomatic phase of the disease are likely to be of most benefit.

Our study has some limitations. Although we analyzed over four hundred *APOE* ε4 homozygotes, there were fewer younger participants in the cohort compared to participants closer to EYO 0 and consequently less power to detect differences at the earliest time points, similar to previous studies in ADAD and DSAD. Although the Bayesian analytical approach employed in our study is well-suited to model protein levels in data sparse regions, future studies that include additional young ε4 homozygotes will be helpful to further understand differences in young adulthood. Related to this point, absolute time estimates of differences in ε4 homozygotes depend on power across the EYO range and the statistical threshold used to determine differences, and although we used a conservative 99 percent CI to estimate differences in ε4 homozygotes, these estimates may change as additional data become available for analysis. However, we do not expect the relative temporal ordering to change. Biomarkers of two important AD endophenotypes, Aβ plaques and tau NFTs, were not available for the participants to examine relationships between proteins altered in ε4 homozygotes and these endophenotypes that begin to develop in mid-life. Future versions of the GNPC dataset that include this information will be required to understand these relationships. Because we restricted our comparator group to unimpaired ε3/ε3 individuals, some protein differences at later EYO time points are likely driven by AD-related neurodegeneration rather than by ε4/ε4 *per se*, but our sensitivity analysis that included impaired ε3/ε3 individuals in the comparator group suggested overall similar results. Ancestry in the GNPC is currently nearly all Caucasian. Future studies that include individuals from diverse genetic backgrounds will be needed to understand how these findings apply to other populations. Future studies may also be able to refine EYO at an individual level using additional genetic information. Our study focused on plasma, but future studies in other compartments such as CSF when cohorts of ε3 and ε4 homozygotes with CSF sampling across the lifespan become available will be helpful to better understand the relationship between peripheral *versus* central effects of ε4 homozygosity over time. It is important to consider that some proteomic alterations in ε4 homozygotes may be compensatory in early-life, preclinical, and clinical disease stages, and if so, reversal of such compensatory changes could be detrimental to brain health. It is also possible that concordant effects of semaglutide in pathways such as RNA processing, ECM, and innate immune at certain EYO epochs could potentially counteract any beneficial reversal effects in other pathways. Our computational predictions of semaglutide effects in ε4 homozygotes require experimental validation in animal models and prospective clinical trials. Prevention trials that manipulate pathways altered in ε4 homozygotes will be required to directly test causality for development of AD.

In summary, we used plasma proteomics to identify biological pathway changes across the adult lifespan in *APOE* ε4 homozygotes, and identified many biological pathways altered in young adulthood that are also altered in AD. We identified key pathways related to metabolism and synaptic/neuronal health that are potentially treatable with GLP-1RAs such as semaglutide. A multi-faceted therapeutic approach to target key AD-related pathways by at least age 50 in ε4 homozygotes will likely have the largest potential for preventing AD in this special population.

## METHODS

### Case Selection and Quality Control of the Full GNPC Dataset

Trait, SomaScan proteomic, genetic, sample-metadata, person-mapping, and lookup data from the GNPC Harmonized Dataset(*17*) v1.3 were obtained through the GNPC ADDI Workbench PostgreSQL database on March 27, 2025. Traits were curated into a single table, removing duplicate joining keys (contributor_code, visit, sample_type, and sample_id). Cleaning of numeric value ranges was performed for each trait upon basic histogram visualization as quality control (QC), setting obvious outliers or sentinel values (for example, “999” for age) to missing. *APOE* genotype coding was harmonized across contributor sites. From these curated traits a multivariate disease group variable was defined, with conflicting diagnoses left as unknown (NA). Individuals with CDR=0 and MMSE ≥ 28 lacking a diagnosis were considered as controls. Samples were restricted to SomaScan blood plasma samples with known sex and excluded sites with contributor codes U (serum only), V (low assay count), or W (low sample and assay count). After proteomic data harmonization, quality control, and genotype imputation procedures described below on the full GNPC dataset, a subset of cases for analysis in this study were selected from the full dataset by including controls that were homozygous for *APOE* ε3 and control, mild cognitive impairment, and AD cases that were homozygous for *APOE* ε4. For cases where multiple longitudinal samples were available, we selected the earliest visit for analysis given our goal to understand early protein changes in ε4 homozygotes. Characteristics of the final selected cohort are provided in **Supplementary Table 3**. For the analysis including cognitively impaired cases in the ε3 homozygote control group, cases were selected using the same approach as the ε4 homozygote group, for a total of *n*=4199 individuals.

Sentinel values of -1 and invalid non-positive RFU values were set to missing before downstream plasma analyses.

### First-Pass Proteomic Data Harmonization and Quality Control

The resulting plasma protein abundance matrix of the full GNPC dataset (7,335 assays × 22,392 samples; log2 relative fluorescence unit (RFU) scale) was subjected to regression of two SomaScan assays we previously determined covary with distinct sources of pre-analytical variance (PAV): heterogeneous nuclear ribonucleoproteins A2/B1 (HNRNPA2B1), which is strongly related to time-to-spin, and hemoglobin subunit zeta (HBZ), which is strongly related to time-to-decant or hemolysis-associated handling effects, as previously described(*94*). Linear regression was performed for each of the 7335 assays separately within each contributor site, modeling log2 abundance as a function of the two PAV assays. Regressed values (7333) within site were then reconstructed as the intercept plus residuals for each assay. Finally, the regressed values for all 19 sites were reassembled into a single matrix.

t-SNE embeddings (*Rtsne* package, perplexity = 20) of the initial regressed matrix were inspected to evaluate contributor site effects. In addition to the 19 sites clearly separating in reduced dimensionality space, samples from site F formed three separable clusters, which were delineated manually by straight lines (segments defined in Cartesian space). Samples from this site were re-labeled into sub-batches F1, F2, and F3 based on position relative to these segmentation lines. This split was codified in the metadata, replacing original codes for site F samples. Visualization outputs documenting inter- and intra-site batch effects are provided in **Supplementary Figure 1**.

A first-pass site regression was then performed after within-site PAV regression, with site F split into F1/F2/F3. For each protein assay, a linear model of the form:

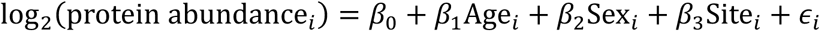

was fitted, where *Sex* was coded 0/1 (female/male). Adjusted abundances were reconstructed as the model intercept plus residuals plus fitted age and sex contributions, omitting fitted site terms, thus protecting age and sex while removing site effects. This first-pass adjusted matrix was used to train *APOE* genotype imputation models.

### APOE Genotype Imputation

Among 22,392 plasma samples, 5,715 had unknown *APOE* genotype after cohort harmonization. To impute *APOE* genotype in these samples, we applied a two-stage machine learning approach on the first-pass harmonized dataset after site regression as described above. In the first stage of computation, we defined the top 50 features for binary prediction of each of the 6 possible *APOE* genotypes, selecting unique top features for each genotype independently. Feature selection reduced the dimensionality from 7,334 assays to 300 features (the top 50 per genotype) to optimize the bias-variance tradeoff. This threshold was chosen to capture the most informative protein signatures while preventing overfitting from high-dimensional noise and maintaining computational efficiency. Feature selection was performed using an ensemble of three machine learning algorithms: elastic net regularized logistic regression (*glmnet*), gradient-boosted decision trees (*xgboost*), and probabilistic random forests (*ranger*). For each genotype, this ensemble was trained using 5-fold cross-validation repeated five times, targeting a positive predictive value (PPV) of 95%. Following cross-validation, a final ensemble model was trained on all samples with complete proteomic data and known *APOE* genotype (*n*=14,758). This function was run once for binary prediction of each genotype. Within a run, per fold, average positive call probabilities from the three learners for included samples in the fold were assigned a threshold using heuristic logic that selected the threshold to equal a probability of at least the target positive predictive value (95%) and when true positives numbered at least 30. Fallback when this criterion was not met set the threshold to the 90th percentile of positive class probabilities; finally, if no positives occurred, the threshold probability was set to 100%. The minimum threshold finally allowed was 80%. Prior to model construction, each feature’s relative log2(abundances) were scaled to Z scores across samples. Feature importance was defined separately for each learner: for *glmnet*, importance was computed as the absolute value of coefficients from cross-validated regularized logistic regression models; for *xgboost*, importance was measured using the gain metric from models trained with binary logistic loss (eta = 0.1, max_depth = 6, subsample = 0.8, colsample_bytree = 0.8, 200 boosting rounds); for *ranger*, importance scores were derived from probability forest models consisting of 500 trees. To account for class imbalance, positive samples were upweighted to 8 during the 5×5 cross-validation. In the final ensemble, positive class weights were also 8, except for the three rare genotypes (ε2/ε2, ε2/ε4, and ε4/ε4), which were set to 12, relative to a negative weight of 1. Following the final ensemble training, feature importance vectors from each learner were standardized to *Z* scores and averaged across learners to obtain a consensus importance score. This process was performed independently for each genotype. The top 50 most important features for prediction of each genotype are provided in **Supplementary Table 2**. In the 5×5 cross-validation, mean (of 25) precision values ranged from 97 to 100 percent for the 6 binary genotype predictions, and recall was consistently 100 percent for each genotype.

In the second stage, final genotype prediction was performed using 14,758 samples with complete *APOE* genotype data as ground truth. Input features consisted of the union of the genotype-specific proteins identified in stage 1, using log2-transformed protein abundances after first-pass regression protecting age and sex. Six binary classifiers (one per genotype class: ε2/ε2, ε2/ε3, ε2/ε4, ε3/ε3, ε3/ε4, ε4/ε4) were each implemented as an ensemble of *glmnet*, *xgboost*, and *ranger*, as described in the first stage learner ensemble that obtained the most important features. Out-of-fold cross-validation determined genotype-specific probability thresholds to optimize positive predictive value. For prediction, the six learners were combined into a multiclass caller that evaluated genotype-specific probabilities in a fixed order prioritizing rare or high-performing calls (ε2/ε4, ε2/ε2, ε4/ε4, ε2/ε3, ε3/ε4, and ε3/ε3); when none exceeded its threshold, the maximum-probability class was assigned. To obtain unbiased performance estimates accounting for threshold and hyperparameter optimization, model performance was evaluated using nested cross-validation. The outer loop consisted of 100 random, genotype-frequency-matched hold-out folds, each reserving 20% of samples exclusively for testing. The 100 hold-out set predictions (*n*=295,400) achieved 91.5 ±1.3% (SD) macro-F1 indicating balanced good performance across all genotype classes and 94.3% test accuracy with low variance (SD, ±0.37%) across the 100 folds, indicating stable generalization in repeated hold-out testing. The confusion matrix for the predicted sample counts underlying prediction accuracy over the 100 outer folds run on 20% hold-out samples is provided in **Supplementary Table 1.** The final model retrained on 14,758 samples achieved 99 percent accuracy when evaluated on the complete dataset, consistent with 94.3% cross-validation test accuracy, indicating stable model performance without severe overfitting. The ensemble trained on all input data was applied to the 5,715 unknown samples, and imputed genotypes were integrated into sample metadata. In the dataset used for analysis, 50 (12.1%) of *APOE*ε4/ε4 and 658 (23.8%) of *APOE* ε3/ε3 genotypes were imputed by this machine learning approach.

### Second-pass Proteomic Data Harmonization Including APOE

After genotype imputation, the final second-pass regression began from the two-PAV within-site adjusted matrix and adjusted for site effects while protecting *APOE* genotype, age, and sex as biological covariates. For each protein assay, a linear model of the form:

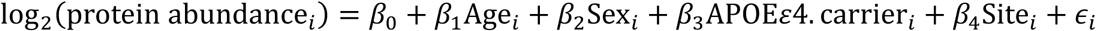

was fitted to the data after two-PAV within-site regression. *Sex* and *APOE*ε4. *carrier* were coded as binary covariates. Residualized abundances plus intercept and protected covariates (*Age*, *Sex*, *APOE*ε4. *carrier*) defined the adjusted matrix (7,333 assays × 22,392 samples, where the two assays used for PAV regression were removed). This two-pass *APOE*-aware regressed matrix was used for WGCNA, and a filtered matrix with only human protein assays (*n*=7,287) for downstream analyses.

In summary, we a) cleaned and harmonized the traits in the GNPC dataset, b) performed a first-step regression to remove as much pre-analytical variation as possible, c) performed first-pass site regression protecting age and sex, d) imputed *APO*E genotypes where needed using a machine learning model, and e) stepped back and revised the site regression approach to protect *APOE* genotype along with age and sex to arrive at the final protein abundance matrix.

### WGCNA Network Construction and Module Detection

Weighted gene co-expression network analysis (WGCNA)(*95*) was applied to the final *APOE*-aware 7,333 × 22,392 log2(abundance) matrix after removal of the two PAV proxy assay rows. Soft-threshold selection (bicor, signed network) identified power = 6 from the elbow/asymptotic region of the signed scale-free topology curve (SFT R²≈0.84). Modules were identified using blockwiseModules with parameters: deepSplit = 4, minModuleSize = 10, mergeCutHeight = 0.07, and TOMDenom = “mean”. Iterative kME reassignment was applied as previously published to refine module membership(*96*). The final network yielded 12 co-expression modules. Module sizes ranged from approximately 1,800 features (M1 turquoise) to less than 100 (M12 tan). Module ontologies were determined using the open-source R function GOparallel (https://www.github.com/edammer/GOparallel/). Human protein ontology annotations were obtained from the Bader lab database assembled in June 2025(*97*).

### Longitudinal Modeling of Plasma Protein Abundances

The primary analysis used the 3,177-sample APOE homozygote contrast, and a secondary 4,199-sample sensitivity cohort retained additional non-control *APOE* ε3/ε3 samples. All individuals had SomaScan 7k plasma proteomic measures available. Prior to constructing models for each protein, we applied pre-specified outlier removal in which values greater than 5 standard deviations (SD) from the mean within each assay were set to missing, and assays with greater than 20 percent missing values within each group were excluded from consideration. Based on these pre-specified criteria, 0.44 percent of assay values were set to missing, and no assays were removed from consideration. Bayesian Gaussian generalized linear model (GLM) regression models were fit independently for each protein assay or module eigenprotein as previously described(*15, 16*), implemented in the *rstanarm* R package which leverages the Hamiltonian Monte Carlo (HMC) algorithm, a dependable Markov Chain Monte Carlo (MCMC) method that enhanced the robustness of our analysis(*98*). To capture non-linear relationships between protein abundance and EYO, we applied a restricted cubic spline transformation to EYO, setting knots at the 10th, 50th, and 90th percentiles. This method breaks down EYO into its linear and cubic components, which guarantees a seamless and consistent model fit at each designated percentile. These components were then used to replace the original EYO for the Bayesian GLM fitting. The use of restricted cubic splines is not only substantiated by prior research validating its capability to model data non-linearities, but also through visual assessments that confirm its concordance with the actual observed data in this study(*15, 16, 99*). The spline transformation was performed using the *Hmisc* R package rcspline.eval function with the parameters nk = 3, norm = 2, pc = FALSE, inclx = TRUE, producing linear and cubic spline components (*EYO*_*L*_, *EYO*_*C*_).

For the regression coefficients, we used the default weak informative normal priors with a mean of 0 and a variance parameter set to 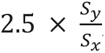, where *S* represents the standard deviation of the outcome measure and *S*_*x*_ represents the standard deviation of the independent covariate. Similarly, the intercept’s prior was also the default prior that is a weak informative normal distribution with a mean of 0 and a variance parameter of 2.5 × *S*_*y*_, promoting an approach that is more objective and data-driven(*100*). The MCMC simulations were conducted by initializing eight chains. Each chain ran for 10,000 iterations, discarding the first 5000 as a warmup. To reduce the data, every tenth iteration post-warmup was selected. We ensured the reliability of the remaining 4000 post-warmup samples by rigorously monitoring the convergence of the parameters. Bayesian two-sided credible intervals for continuous outcomes were estimated for both the ε3/ε3 and ε4/ε4 groups, as well as for the differences between these groups, at increments of 0.5 EYO units ranging from −46 to 25. EYO 0 was defined at 65.6 years of age(*9*). Additionally, we computed the empirical *p* value to evaluate the likelihood of an observed difference under the null hypothesis.

Model 1 tested *APOE* genotype differences in longitudinal EYO trajectories:

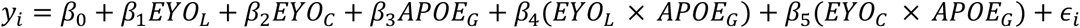

**[Model 1; Base model]**

Model 1 was fit for the 3,177-sample primary cohort, repeated in the 4,199-sample sensitivity cohort, and refit in a no-imputation sensitivity analysis restricted to directly observed *APOE* genotypes.

G is a binary indicator of *APOE* ε4/ε4 status (ε3/ε3 = 0), and in

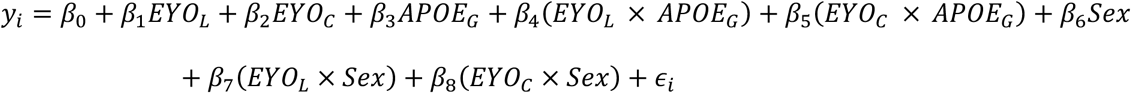

**[Model 2; Sex-stratified EYO model]**

where female = 0, male = 1 for *Sex*. Only proteins that were found to have both an interaction with *APOE* genotype and EYO in Model 1 and an interaction with sex and EYO in Model 2 were considered to be affected by sex.

### Ontology Enrichments by EYO

The 878 significant proteins from Model 1 were grouped into 5-year bins with center incremented every 1-year (67 bins, −43.5 to +22.5 EYO midpoints) and were further annotated for directionality of difference. Gene symbols from significant assays in each bin were input to the GOparallel gene ontology enrichment test framework (https://www.github.com/edammer/GOparallel). For each bin, overrepresentation of GO terms (Biological Process, Molecular Function, Cellular Component) and Reactome terms was tested using Fisher’s exact tests (signed, one-sided, with Benjamini-Hochberg correction for multiple testing). This was repeated for gene symbol lists of increased and decreased proteins separately. Ontologies were from the Bader Lab database of human ontologies(*97*) downloaded June 2025. A minimum number of three genes was required for enrichment to be considered. Positive enrichment *Z* scores from enrichment analyses were combined across the binned five-year windows into matrices of ontology × EYO bins for GO and Reactome categories. The top 100 ontologies in each category by *Z* score were selected for manual curation and ontology clustering by related pathways, resulting in 141 final ontologies clustered into 18 groups for visualization.

### Overlap with Other Studies

Overlap of unique gene symbols for proteins different in ε4 homozygotes prior to age 50 (*n*=310, **Supplementary Table 4**) was performed against proteins associated with dementia risk after multiple comparisons in the Atherosclerosis Risk in Communities (ARIC) cohort(*20, 21*) (*n*=32 and *n*=37); with AD risk in the AGES-Reykjavik cohort(*12*) (*n*=35); with dementia risk in the Whitehall II cohort(*24*) (*n*=21); and with dementia risk in the UK Biobank(*25*) (*n*=78). Overlap was also performed against proteins nominally associated with risk of cognitive impairment in the Religious Orders Study and Rush Memory and Aging Project (ROSMAP) cohort(*18*) (*n*=427).

To quantify directional concordance with external proteomic studies, summary statistics from this study and each comparator dataset were converted to signed rank scores at the gene-symbol level. For the present study, protein-level F statistics (or in the case of Age, –log10(*p*) for bicor) were multiplied by the sign of the reported directional effect, and when multiple assays mapped to the same gene symbol, the assay with the largest absolute signed statistic was retained. Signed statistics were ranked from most negative to most positive and scaled to the interval 0–1, with higher values indicating stronger positive effects. External study statistics from Walker *et al*.(*14*), Shvetcov *et al*.(*8*), Afshar *et al*.(*18*), Imam *et al*.(*17*), Frick *et al*.(*12*), and Coenen *et al*.(*101*) were processed similarly by multiplying each reported statistic by its direction, assigning ranks relative to the total number of measured symbols in that study, and scaling ranks to 0–1. External scaled ranks were then matched to the present study’s scaled ranks by gene symbol. A Spearman rho and associated Student’s *p* value for directional agreement between each external study and the corresponding scaled, ranked full data for the present study were assessed using Pearson correlation of the matched scaled ranks with WGCNA::corAndPvalue.

### Semaglutide Effects

We quantified pathway-level connectivity between plasma proteomic signatures of *APOE* ε4 homozygosity and the proteomic response to semaglutide as described in Maretty *et al*.(*30*), using summary statistics from a model adjusted for baseline and change in body mass index (BMI) and HbA1c, thereby capturing proteomic effects separate from those due to weight loss and glycemic improvement. *APOE* ε4/ε4 effect sizes (ES) were obtained from the Bayesian GLM described above across 143 EYO timepoints, represented as a matrix ES(*t, j*) of signed effect sizes for assay *j* across EYO timepoints *t* (0.5-year increments spanning EYO –46 to +25). Analyses focused on three non-overlapping EYO intervals approximating early-life (EYO –46 to –15.5), preclinical AD (EYO –15 to –0.5), and clinical AD (EYO 0 to 25) stages. For semaglutide pathway effect analyses, we used a human gene matrix transposed (GMT) collection of GO and pathway gene sets as described above and parsed it into a list of gene sets, with ontology identifiers mapped to

HUGO Gene Nomenclature Committee (HGNC) gene symbols. Ontology names and types were standardized to enable exact matching to the previously defined set of 141 ontologies (**Supplementary Tables 9-11**) used for downstream scoring.

Semaglutide effects were represented as assay-level signed T statistics derived from the model in Maretty *et al*. adjusted for BMI and HbA1c covariates. For each assay *j*, we confirmed the provided statistic was computed

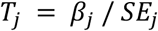

where β is the reported semaglutide effect size and SE is its standard error. SomaScan assays from Maretty *et al*. were matched on aptamer sequence ID. The resulting ranked vector contained 7,287 human assays. For each ontology-specific gene symbol list, semaglutide ranked statistics were restricted to assays whose gene symbol belonged to the ontology. When multiple assays mapped to the same gene symbol, we retained a single representative assay per gene by selecting the assay with maximal |*T*| (sign preserved), yielding an ontology-specific ranked vector *r* of length *N*.

Within a specified EYO interval, we summarized each assay’s ε4/ε4 genotype signature by its peak signed ES: for assay *j*, ES*j was defined as the value at the timepoint t* where |ESt, j| was maximal within the interval. Assays were retained if |ES*j| was greater than or equal to 0.10. The set of assays increased in ε4/ε4 were those with positive ES*j, and the set of assays decreased in ε4/ε4 were those with negative ES*j. When multiple GNPC assays represented the same gene, the assay matching the retained semaglutide representative was prioritized; otherwise, the assay with maximal |ES*j| was retained. Finally, ε4/ε4-increased and -decreased sets were intersected with the ontology-specific semaglutide assay list to ensure scoring was performed on a common feature space.

Connectivity was computed using a weighted enrichment statistic closely related to Gene Set Enrichment Analysis (GSEA) and the weighted connectivity score used in the Connectivity Map framework(*102–104*). For a given ontology, let *r* be the semaglutide ranked vector sorted in decreasing order (assays *a*1…*a*N with values *r*1…*r*N), and let *S* be a query set (ε4/ε4-increased or -decreased assays). Define indicator *I*i = 1 if *a*i ∈ *S* and 0 otherwise. Using weighting exponent *p* = 1, weights were *w*i = |*r*i|^p^. The running-sum statistic was computed as:

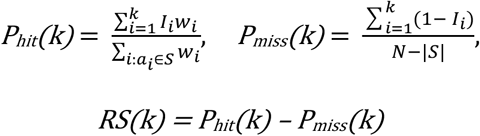

The enrichment score ES(*S*; *r*) was defined as the extreme deviation of RS(k), choosing the maximum or minimum with the largest absolute magnitude:

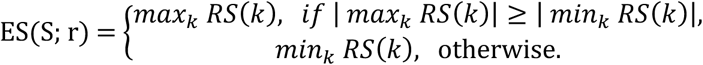

This corresponds to the weighted Kolmogorov-Smirnov-like statistic described for GSEA(*104*). We computed ESup = ES(Sup; r) and ESdown = ES(Sdown; r). The weighted connectivity score (C score) was then:

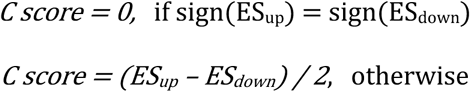

C score is bounded in [−1, 1]; positive values indicate concordant directionality between *APOE* ε4/ε4 effects and semaglutide effects within the ontology, whereas negative values indicate inverse connectivity (opposing directionality), consistent with the Connectivity Map interpretation(*102, 103*).

Connectivity was evaluated first per EYO timepoint across the full 143×141 (time×ontology) grid by defining Sup(t) and Sdown(t) from the sign and magnitude of ESt, j at each timepoint, and second within non-overlapping EYO ranges by defining Sup and Sdown from ES*j within each range. For each analysis, we recorded C score along with the individual increased- and decreased-set enrichment components, and the counts and assay identifiers contributing to each component (**Supplementary Table 12**).

Permutation-based significance testing was performed for each ontology-by-EYO-epoch cell by holding the drug ranking fixed and sampling random up and down query sets of the same sizes as the observed sets from the ontology-specific assay universe. The primary adjusted semaglutide analysis used 100,000 permutations per cell, and sensitivity sweeps repeated the analysis across *APOE* effect-size thresholds.

### Other Statistics and Visualization

Analyses were performed in R v4.5.1. Bayesian modeling was performed with R stan (rstan v2.32.7, which uses Stan v2.32.2 running under R v4.5.1). Differences between groups at the 99 percent credible interval (CI) were considered significant. Proteins with at least one EYO interval that was considered significant (*p* ≤ 0.005, 99% two-tailed CI) were used for heatmap visualizations. *P* values that were calculated as zero values were replaced with the minimum detectable value (1/8000, when none of 4000 MCMC iterations were insignificant). Heatmaps were generated in Python (v3.10) using the *seaborn* package. Ontology heatmaps were generated with the ComplexHeatmap R package; semaglutide connectivity heatmaps were exported as ComplexHeatmap figures. Significant ontology enrichment for all proteins was considered at *z* < 1.645 (or *p* < 0.05), and for separate increased or decreased proteins at *z* < 1.3 (or *p* < 0.1). Graph layouts of module member proteins organized by their intramodular kME (bicor, correlation to the first principal component of a module) were generated using the netOps buildIgraphs function (https://www.github.com/edammer/netOps/). Venn diagrams with proportionate overlaps were constructed using the R venneuler package v1.1-4. Differences between groups in the M9 eigenprotein analysis were assessed by one-way ANOVA, with differences considered significant at *p* < 0.05. Correlations were performed using biweight midcorrelation (bicor).

## Supporting information

Dammer et al. Supplementary Tables

Dammer et al. Supplementary Information

## Data Availability

The GNPC dataset is available at
https://discover.alzheimersdata.org/catalogue/datasets/e2f3536b-d97b-4303-89bd-6864200807a4

## List of Supplementary Materials

Supplementary Figures 1 to 13

Supplementary Tables 1 to 12

## Acknowledgements

We thank the research participants who made this study possible.

## Author Contributions

E.B.D, S.A., and E.C.B.J. designed the experiments; E.B.D and S.A. carried out experiments; E.B.D, S.A., and E.C.B.J. analyzed data; S.B., N.T.S., A.I.L., and J.F. provided advice on the interpretation of data and manuscript review; E.C.B.J., E.B.D, and S.A. wrote the manuscript with input from coauthors.

## Data, Code, and Materials Availability

The GNPC dataset is available at https://discover.alzheimersdata.org/catalogue/datasets/e2f3536b-d97b-4303-89bd-6864200807a4. Code is available at https://github.com/edammer/E4longitudinalPlasma.

---

**Supplementary Figure 1.**
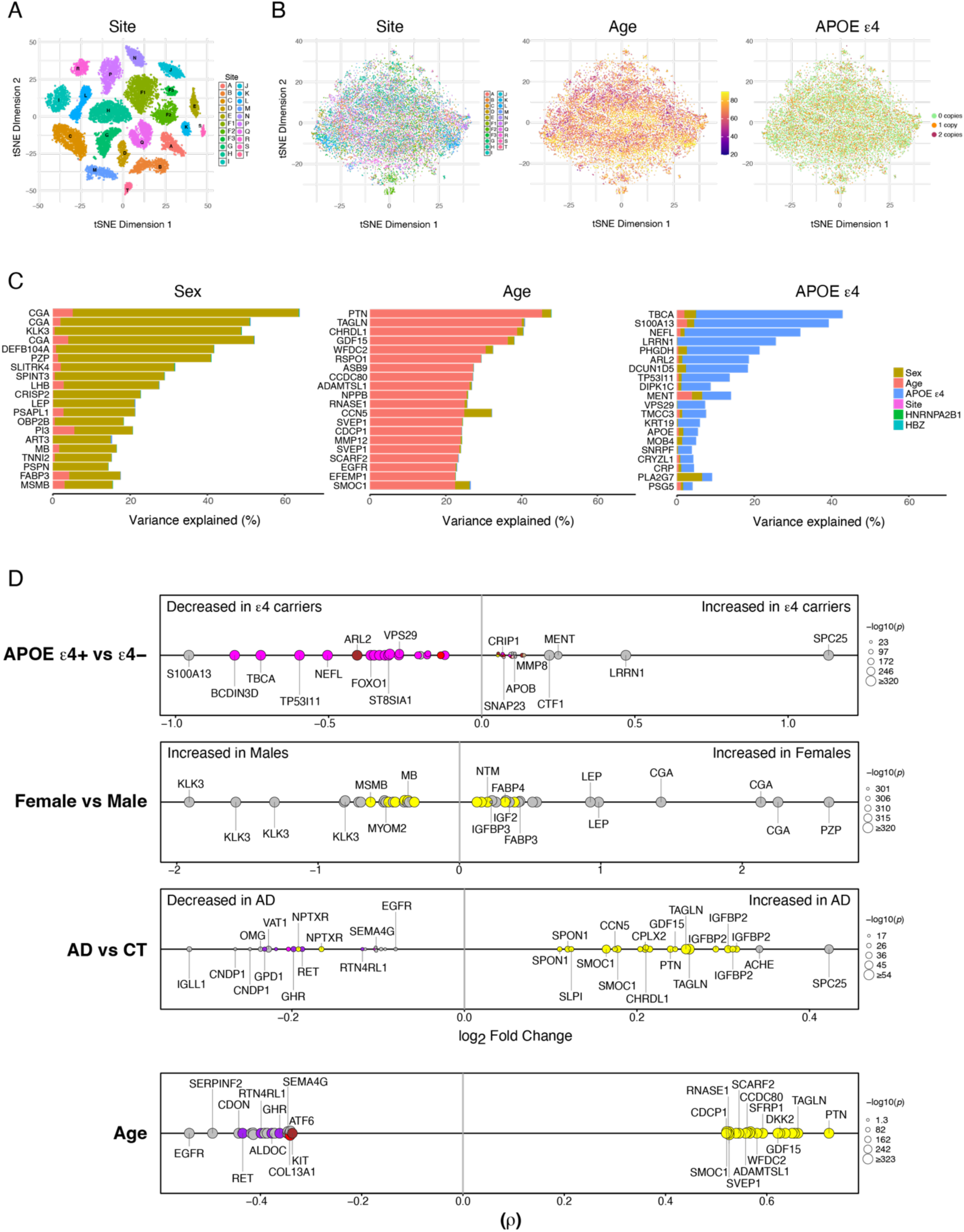
GNPC Data Harmonization and Quality Control. Harmonization was performed on *n*=22,392 plasma samples from cohorts A-T. Cohort O was excluded because it contained only CSF samples. Cohort F was split into 3 groups. (A) t-distributed stochastic neighbor embedding (tSNE) plot of GNPC cohorts after intra-site time-to-spin adjustment but prior to cohort harmonization. (B) tSNE plots for site, age, and *APOE* ε4 allele count after site regression with protection for age, sex, and *APOE* ε4 carrier status. (C) Variance partition plots for sex, age, and *APOE* ε4 showing the top 20 proteins associated with each trait. Only proteins that were present in all datasets were used for variance partition analysis, and therefore proteins present only in either the SomaScan 5k or 7k platform were excluded. Glycoprotein hormones alpha chain (CGA), prostate-specific antigen (KLK3), and pregnancy zone protein (PZP) are top proteins associated with sex; pleiotrophin (PTN) and growth/differentiation factor 15 (GDF15) with age; and tubulin-specific chaperone A (TBCA) with *APOE* ε4. (D) Differential effect size of the top 20 positively and negatively associated proteins for *APOE* ε4, sex, and AD, and the top protein correlations (biweight midcorrelation) with age.

**Supplementary Figure 2.**
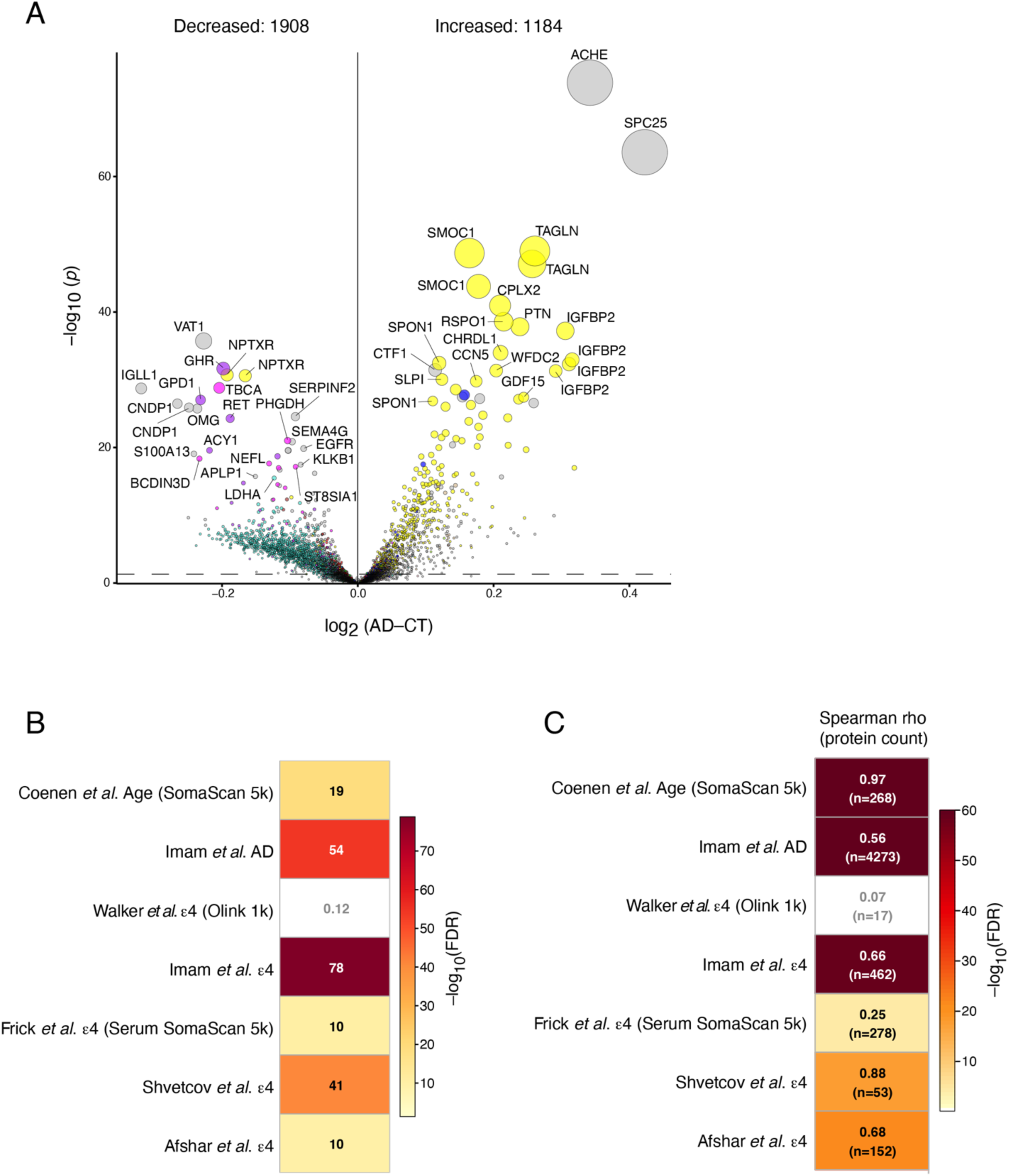
Concordance with Other Studies. (A) Differential abundance of plasma proteins in AD. Data are from *n*=5,485 unique individuals (*n*=1,030 AD and *n*=4,455 controls, last longitudinal visit) in the GNPC dataset after harmonization and quality control procedures. The horizontal line indicates threshold for statistical significance. Proteins are colored according to the protein co-expression module in which they reside; gray proteins do not map to a module. Size of circle is proportional to statistical significance. Proteins represented by more than one gene symbol indicate different Somamer assays to the same protein. (B) Overlap of plasma proteins associated with age as described by Coenen *et al*.(*101*); with AD as described by Imam *et al*.(*17*); and with *APOE* ε4 as described by Walker *et al*.(*14*), Imam *et al*.(*17*), Frick *et al*.(*12*), Shvetcov *et al*.(*8*), and Afshar *et al*.(*18*) with proteins associated with age, AD, and ε4 in this study. *P* values were adjusted for multiple testing across studies. Proteins in Coenen *et al*. and Frick *et al*. were measured on the SomaScan 5k platform. Proteins in Frick *et al*. were measured in serum rather than plasma. Proteins in Walker *et al*. (1,104) were measured on the Olink proximity extension assay platform. Numbers indicate –log10(FDR) values. (C) Rank correlation of the significant proteins reported in each study in (B) with significant proteins in this study. Significance for Imam *et al*. AD correlation was greater than –log10(300), but scale is set to max –log10(60) for visualization purposes.

**Supplementary Figure 3.**
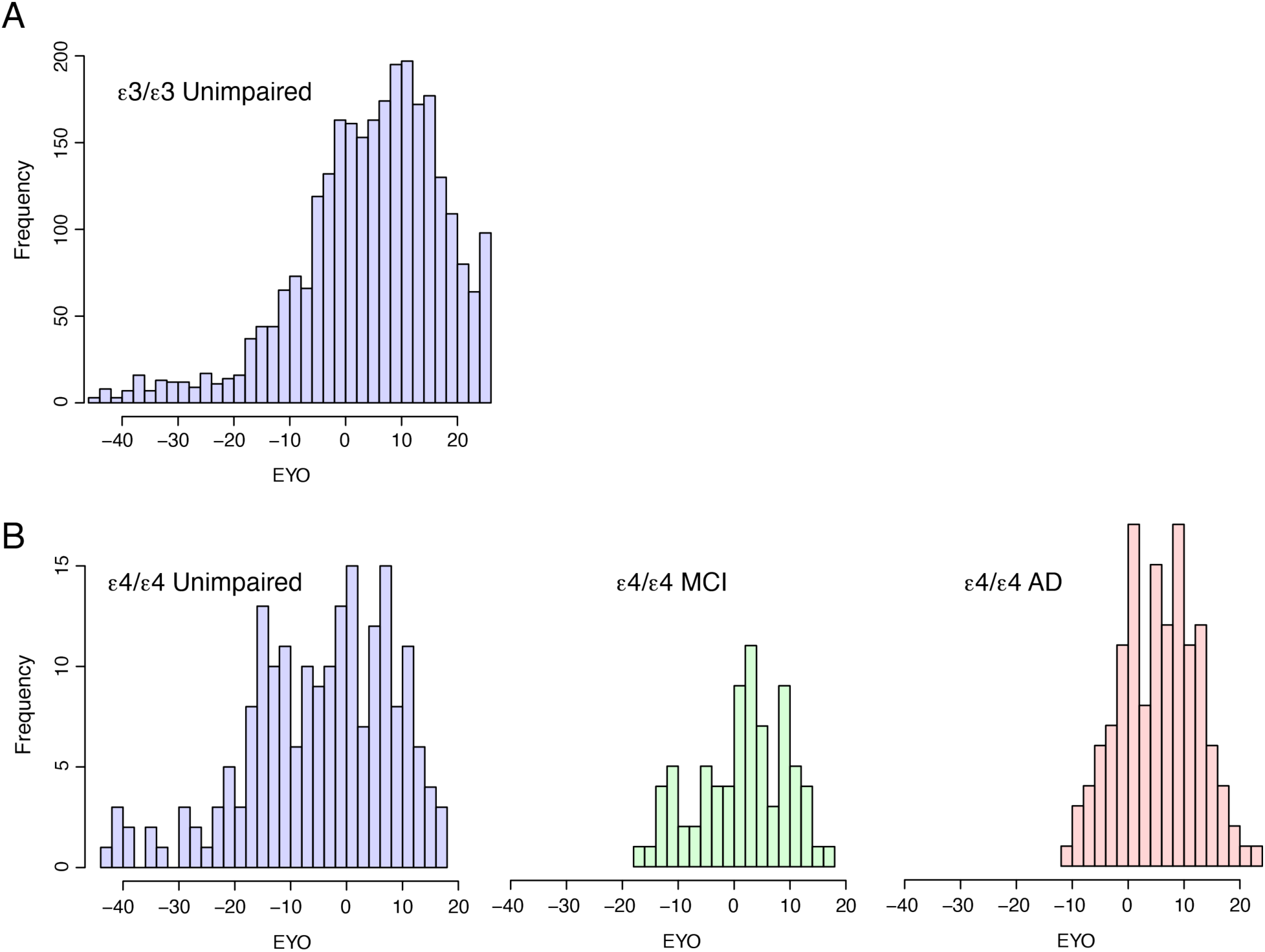
Distribution of Cases by Genotype across EYO. Data from *n*=3177 individuals used for modeling protein changes in *APOE*ε4 homozygotes across estimated year of onset (EYO). (A) Distribution of cases in *APOE* ε3/ε3 controls. (B) Distribution of cases in *APOE* ε4/ε4 unimpaired (left), mild cognitive impairment (MCI, center), and AD (right).

**Supplementary Figure 4.**
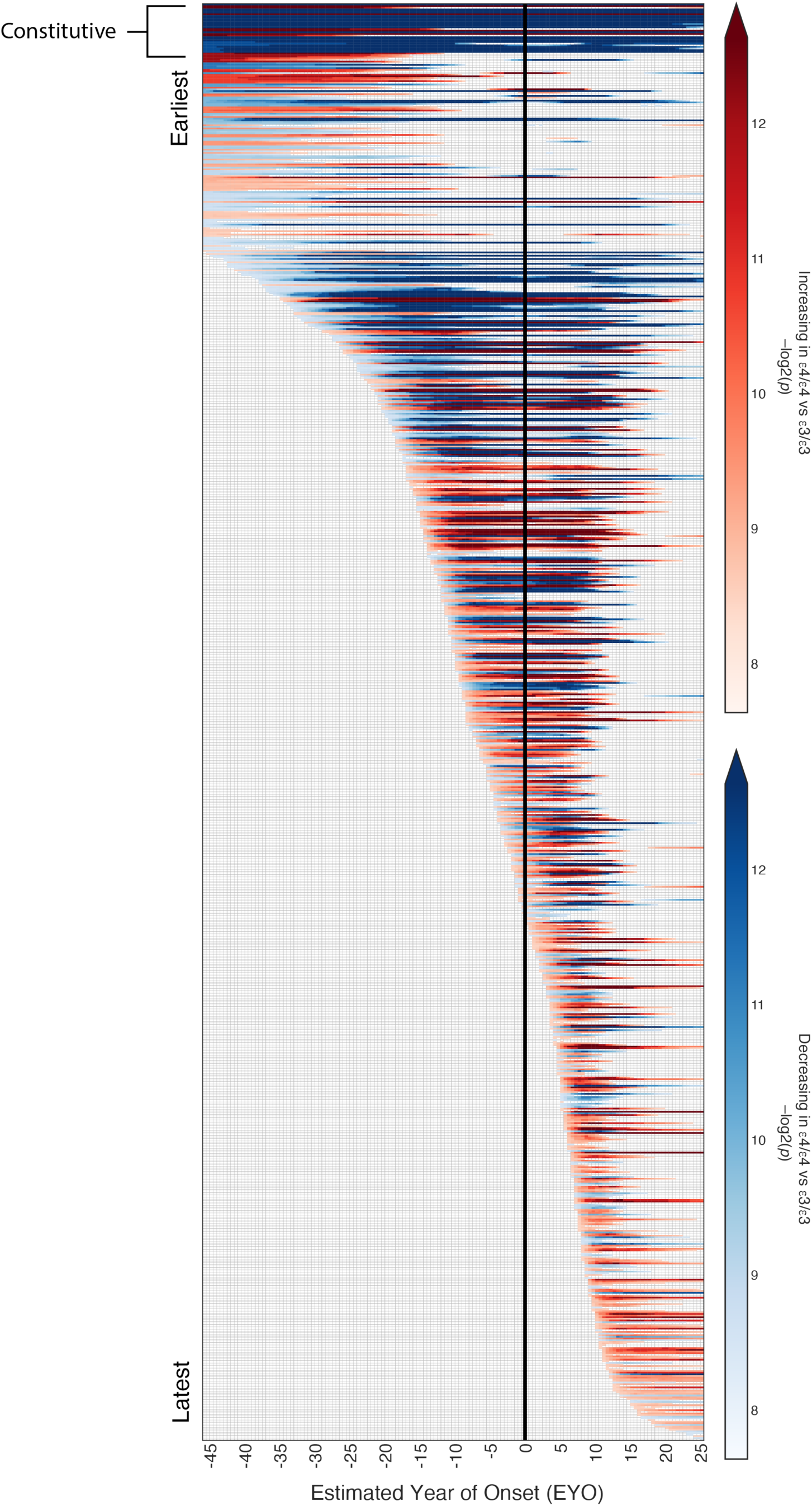
Proteins Altered in *APOE* ε4 Homozygotes by EYO. A total of *n*=878 proteins were different in ε4 homozygotes at any EYO as assessed in 0.5 EYO increments. Red indicates proteins increased in ε4 homozygotes; blue indicates proteins decreased in ε4 homozygotes. Heat is proportional to degree of statistical significance. The vertical black line indicates EYO 0, or age 65.6. Data for individual proteins are provided in **Supplementary Table 4**.

**Supplementary Figure 5.**
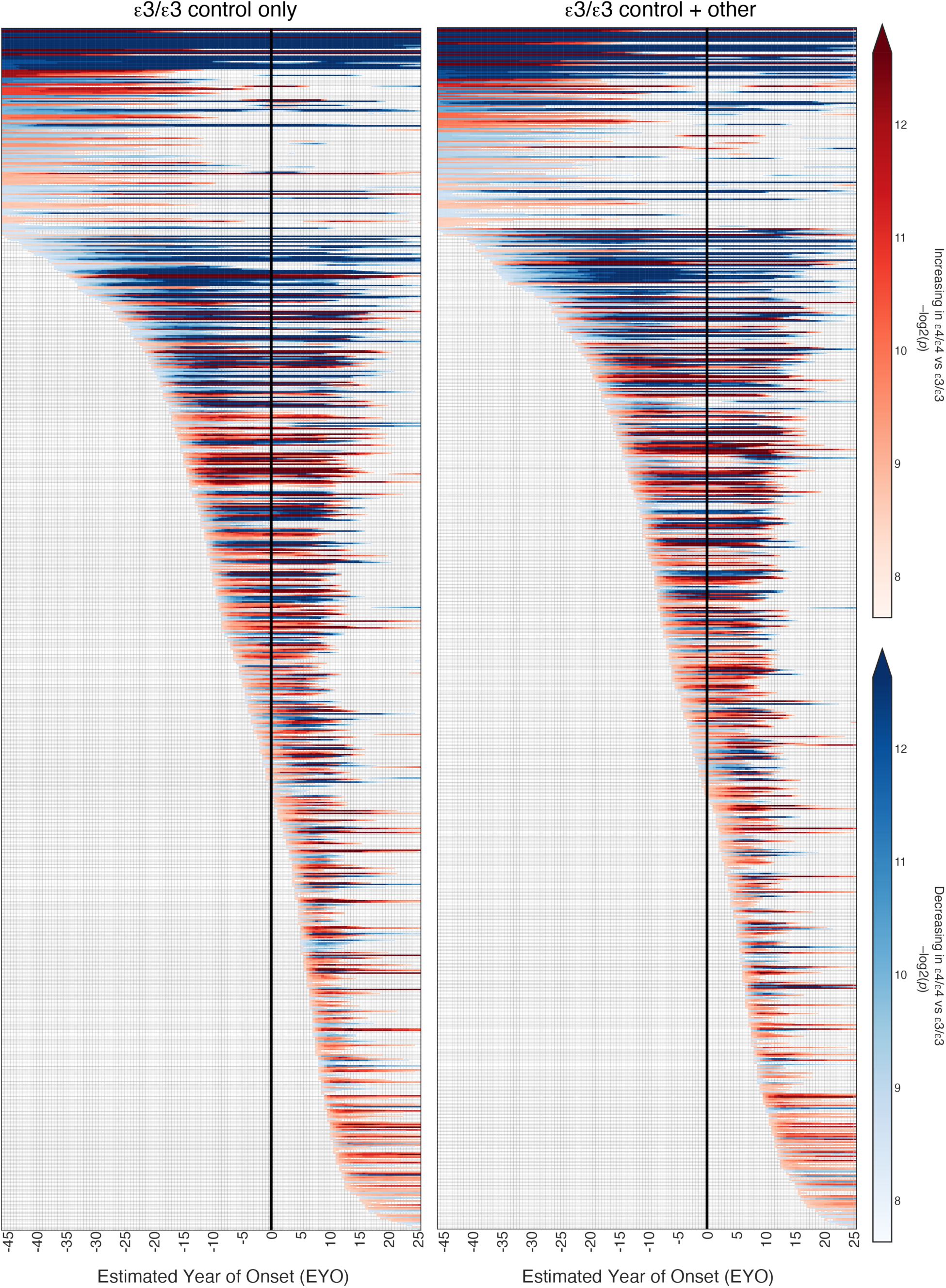
Proteins Altered in *APOE* ε4 Homozygotes by EYO Using Different ε3/ε3 Control Groups. (Left) Proteins altered in ε4 homozygotes using ε3/ε3 unimpaired controls only (*n*=3177 cases, *n*=878 significant proteins). (Right) Proteins altered in ε4 homozygotes using ε3/ε3 controls including MCI and AD (*n*=4199 cases, *n*=848 significant proteins). Red indicates proteins increased in ε4 homozygotes; blue indicates proteins decreased in ε4 homozygotes. Heat is proportional to degree of statistical significance. The vertical black line indicates EYO 0, or age 65.6.

**Supplementary Figure 6.**
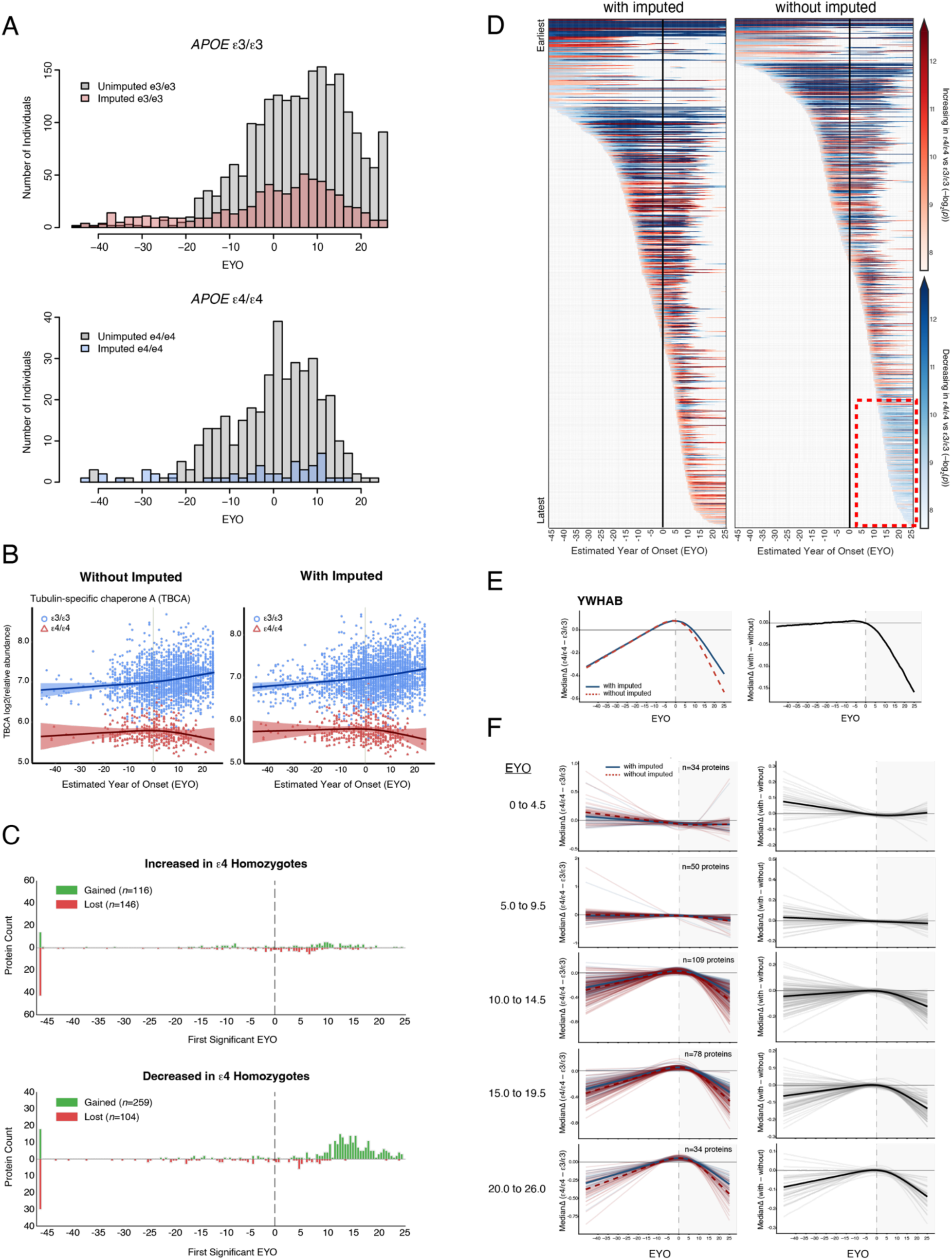
Protein Modeling without Imputed *APOE* Genotypes. (A) Number of individuals by imputation status across estimated year of onset (EYO) for *APOE* ε3/ε3 (top) and *APOE* ε4/ε4 (bottom) genotypes, grouped in two-year bins. (B) Tubulin-specific chaperone A (TBCA) levels in ε3 and ε4 homozygotes across EYO modeled without (left) or with (right) inclusion of individuals with imputed *APOE* genotypes. (C) Number of proteins increased (top) or decreased (bottom) in ε4 homozygotes gained or lost after exclusion of individuals with imputed *APOE* genotypes, grouped by earliest EYO significance. The vertical dashed line indicates EYO 0. (D) Proteins altered in ε4 homozygotes by EYO with (left) or without (right) imputed genotypes. Heat is proportional to degree of statistical significance. The vertical black line indicates EYO 0. The red dashed box highlights the group of proteins decreased in ε4 homozygotes at later EYO timepoints when imputed samples are not included in the model. (E) An example of the median difference between ε4/ε4 and ε3/ε3 individuals across EYO for 14-3-3 protein beta/alpha (YWHAB) modeled with or without imputed cases (left), and the difference between the median differences by EYO (right). (F) Proteins that gain significance after exclusion of imputed cases by five-year epoch after EYO 0. (Left) Individual protein differences between ε4/ε4 and ε3/ε3 by EYO for proteins that gain or lose significance after EYO 0, by five-year epochs. The colored dashed and solid lines indicate the median difference without and with imputed cases, respectively. (Right) Difference between the median differences by EYO.

**Supplementary Figure 7.**
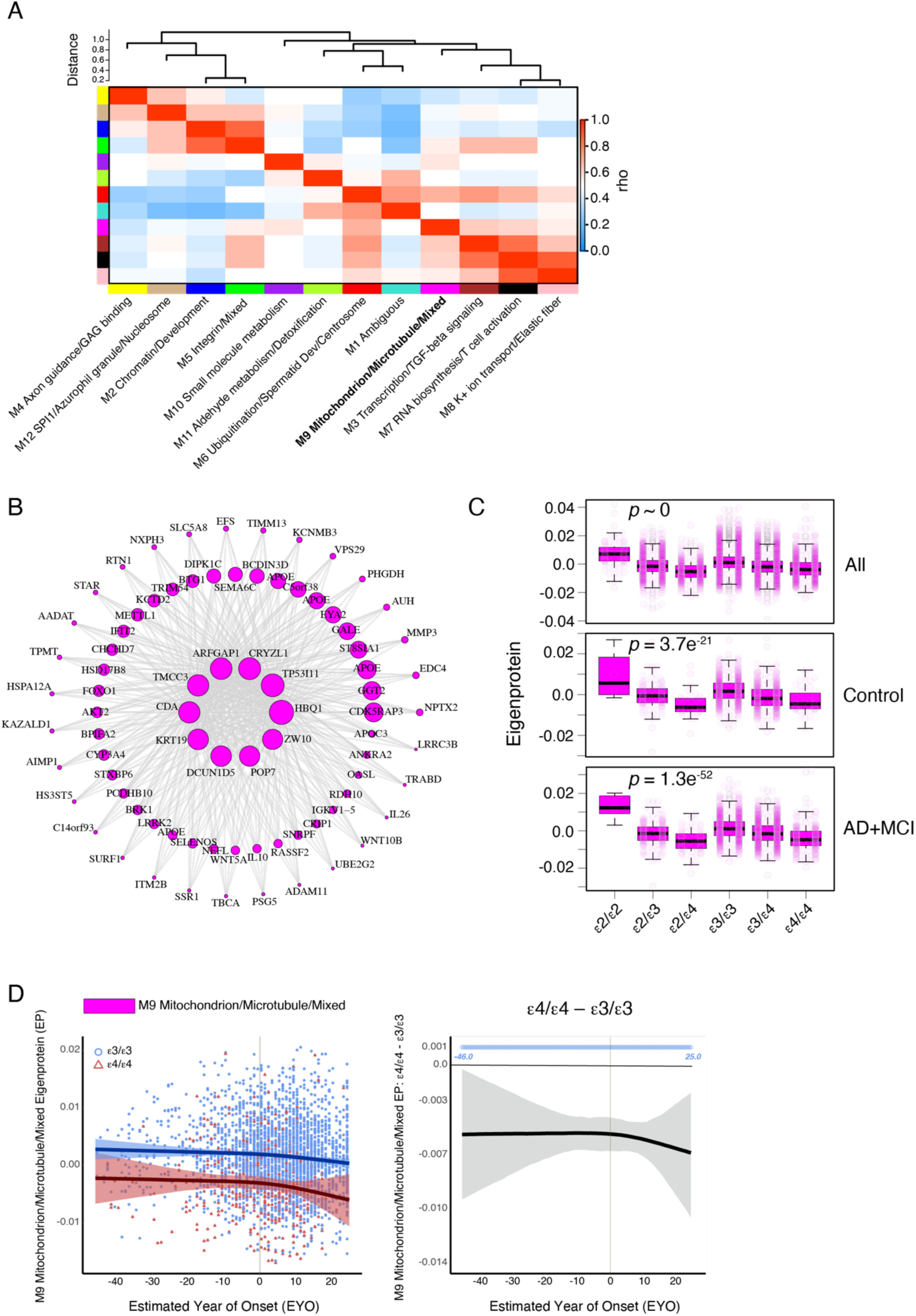
Protein Co-expression Network of GNPC Dataset. A total of *n*=22,392 samples were used to construct a plasma protein co-expression network (A). Heat represents correlation (bicor) of module eigenproteins. Modules were annotated with their primary ontologies. Ontology information is provided in **Supplementary Table 6** and **Supplementary Information**. (B) Proteins in the M9 Mitochondrion/Microtubule/Mixed module. Size of the circles correspond to the network kME value, or degree of correlation to the module eigenprotein. (C) Levels of M9 by *APOE* genotype in all individuals (top, *n*=17,484 total), control only (middle, *n*=4,455 total), and AD plus MCI individuals (bottom, *n*=2,095 total). Differences were assessed by one-way ANOVA. (D) (Left) Levels of M9 across EYO in ε4/ε4 (red, *n*=413) and ε3/ε3 (blue, *n*=2764) individuals. (Right) Difference between ε4/ε4 and ε3/ε3 individuals across EYO. The blue circles indicate periods of significantly lower levels of M9 in ε4/ε4 individuals. Shaded areas indicate the 99 percent credible interval.

**Supplementary Figure 8.**
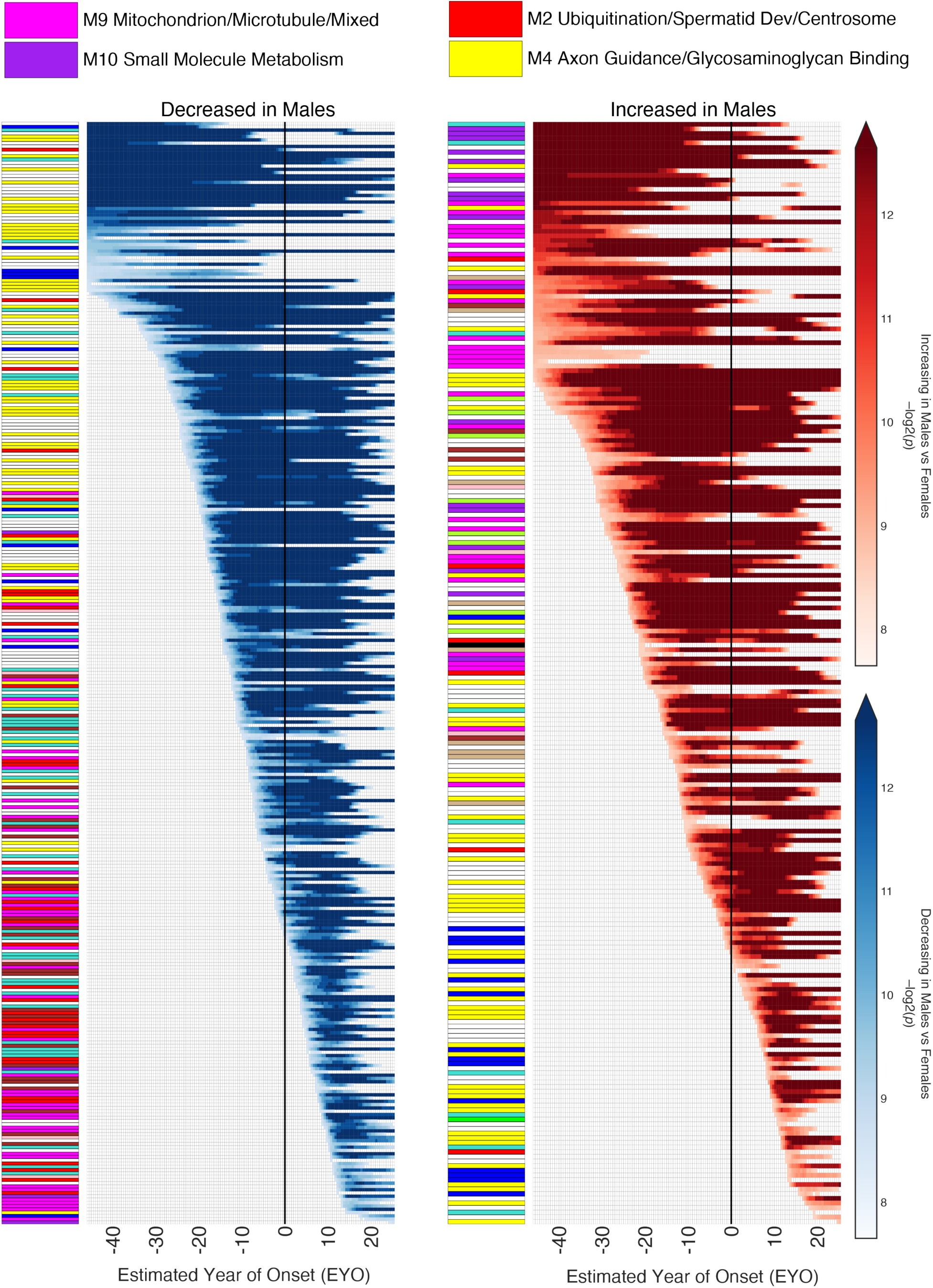
Proteins Altered in ε4 Homozygotes Influenced by Sex. *n*=533 out of 878 proteins were influenced by sex. (Left) Proteins decreased in males. (Right) Proteins increased in males. Heat is proportional to degree of statistical significance. The vertical black line indicates EYO 0, or age 65.6. Each protein was annotated with the plasma protein co-expression network module in which it resides. Network module colors and ontology annotations are provided in **Supplementary** Figure 7A. Individual protein information is provided in **Supplementary Table 7**.

**Supplementary Figure 9.**
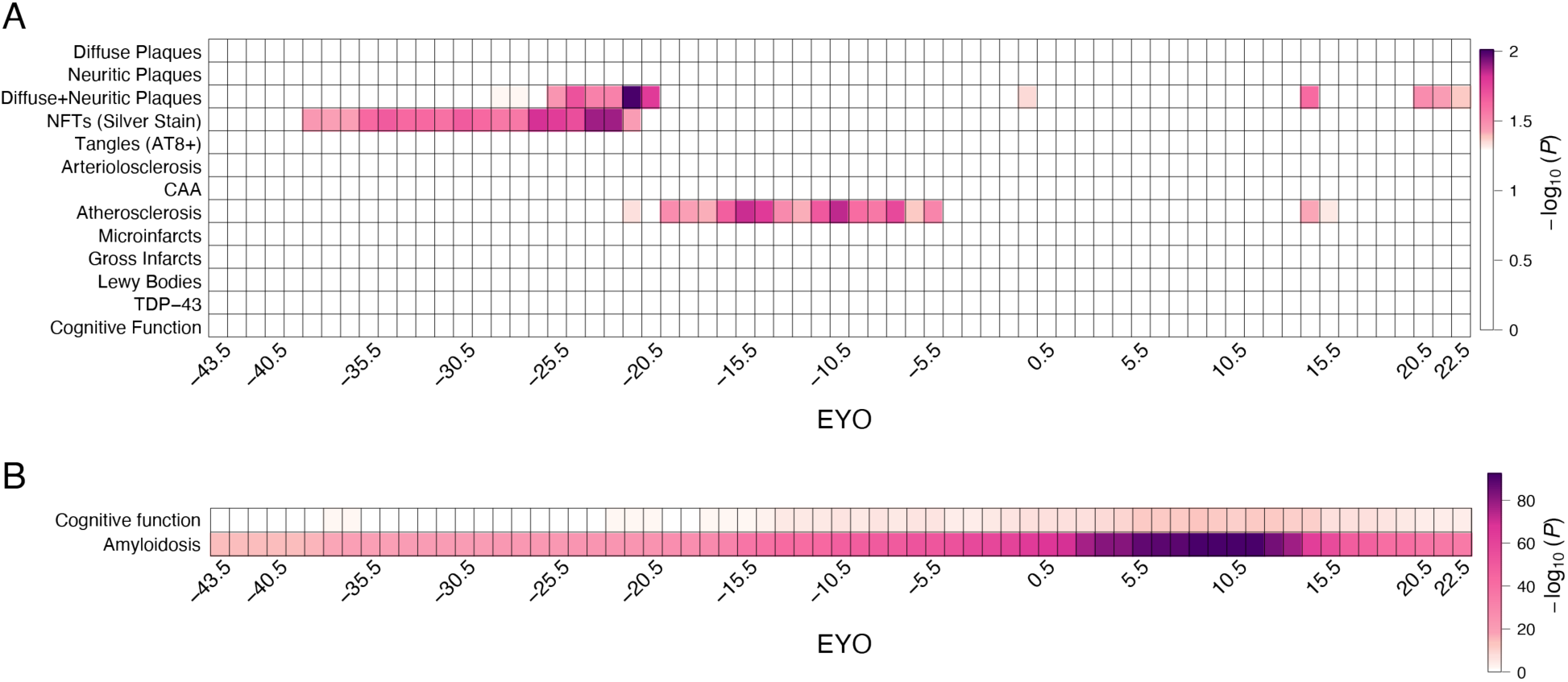
Overlap of ε4/ε4-associated Proteins with AD Endophenotypes. Proteins altered in ε4 homozygotes at different EYO intervals were assessed for enrichment in plasma proteins associated with AD endophenotypes as described in Afshar *et al*.(*18*) using a 5-year sliding window. Periods of significance for each endophenotype are indicated with heat. (A) Enrichment for plasma proteins associated with different ROSMAP endophenotypes. (B) Enrichment for plasma proteins associated with cognitive function and cerebral β-amyloidosis after meta-analysis across four cohorts. Enrichments are reported at a nominal level and do not survive FDR correction in (A).

**Supplementary Figure 10.**
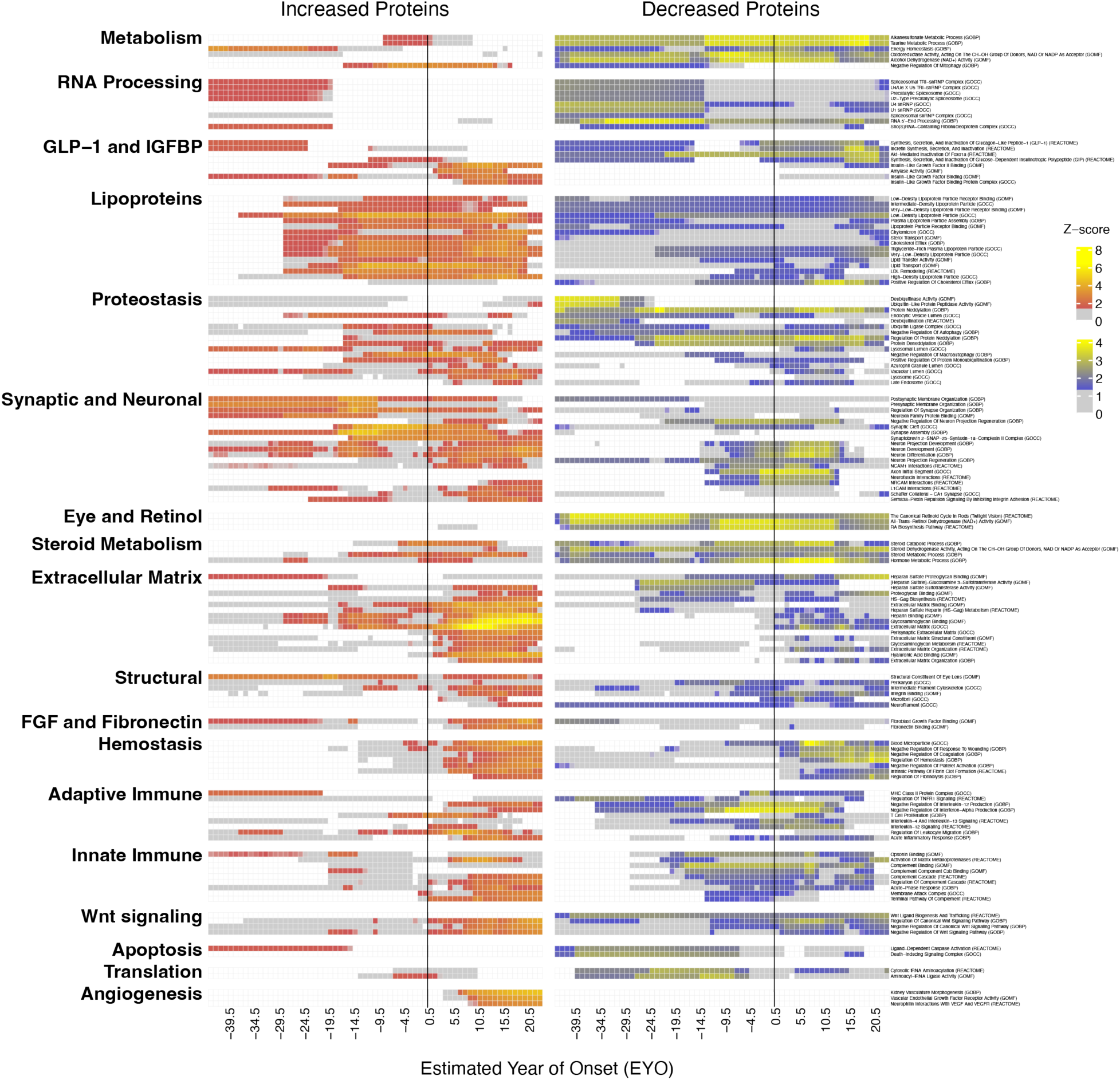
Biological Pathways Enriched in Proteins Increased or Decreased in ε4 Homozygotes. The top pathways enriched in proteins increased (left, red) or decreased (right, blue) in ε4 homozygotes from the GO biological process (GOBP), molecular function (GOMF), cellular component (GOCC), and Reactome databases were grouped by category, with 0.5 EYO periods of significant enrichment using a 5-year sliding window indicated by heat color and near-significant enrichment indicated by gray. The vertical black line indicates EYO 0, or age 65.6. Information on pathways and periods of enrichment is also provided in **Supplementary Tables 10 and 11**.

**Supplementary Figure 11.**
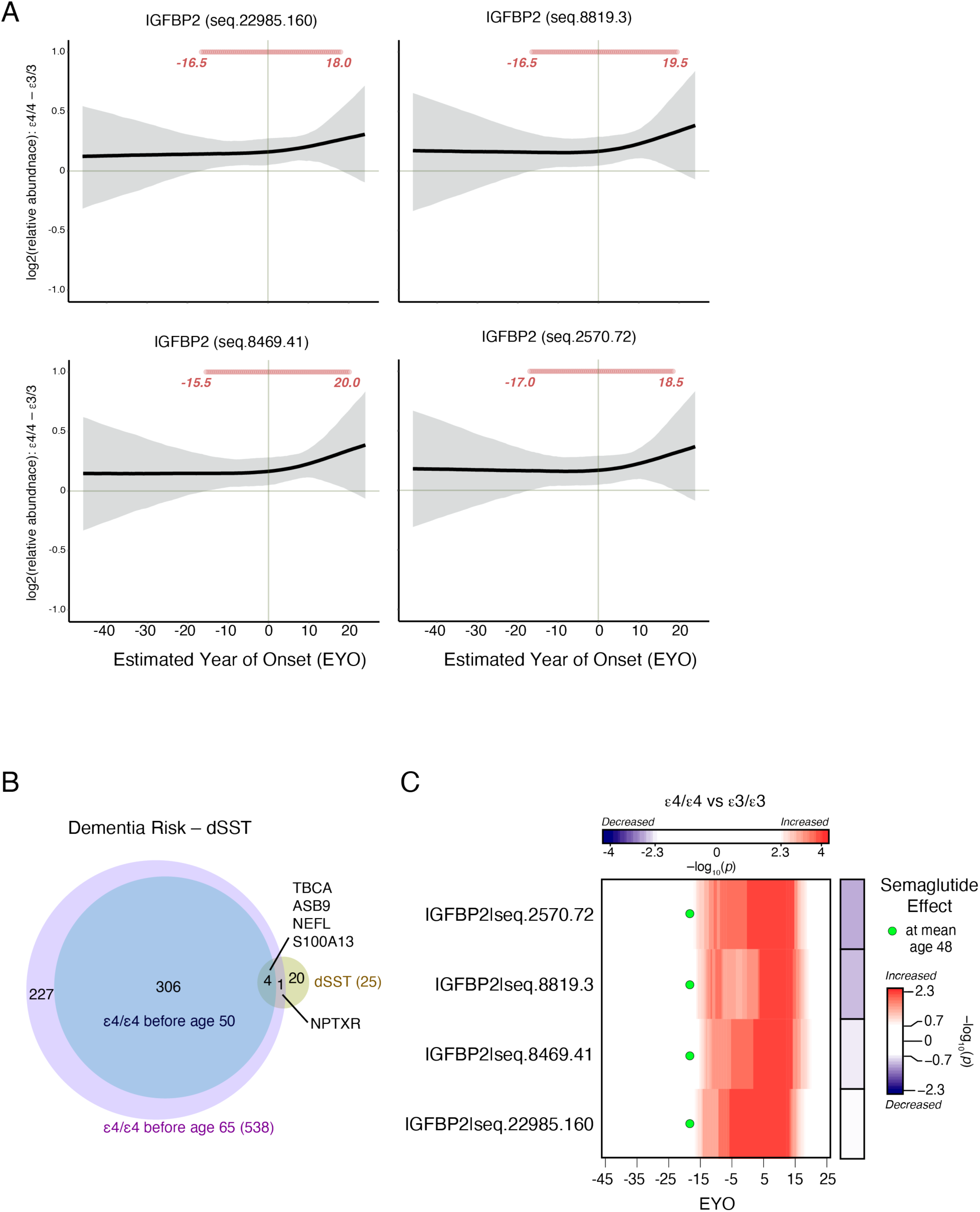
Increased IGFBP2 Levels in ε4 Homozygotes, Overlap with the dSST, and Semaglutide IGFBP2 Reversal Effect. (A) Difference in IGFBP2 protein levels between ε3/ε3 and ε4/ε4 individuals as measured by four different Somamers. The red circles indicate periods of significantly higher levels of IGFBP2 in ε4/ε4 individuals. Shaded areas indicate the 99 percent credible interval. (B) Proteins altered in ε4 homozygotes prior to age 50 and prior to age 65 were assessed for overlap with the 25 proteins used in the dementia SomaSignal® test (dSST) that provides 5-year and 20-yr dementia risk estimates(*27*), with common proteins highlighted. Overlap was not statistically significant. (C) Same data in (A) plotted by heatmap (red, increased levels in ε4 homozygotes), with the effect of semaglutide for each IGFBP2 assay provided to the right (blue, decreased with semaglutide treatment). Green circles indicate average age (48) of participants in the STEP1 semaglutide trial.

**Supplementary Figure 12.**
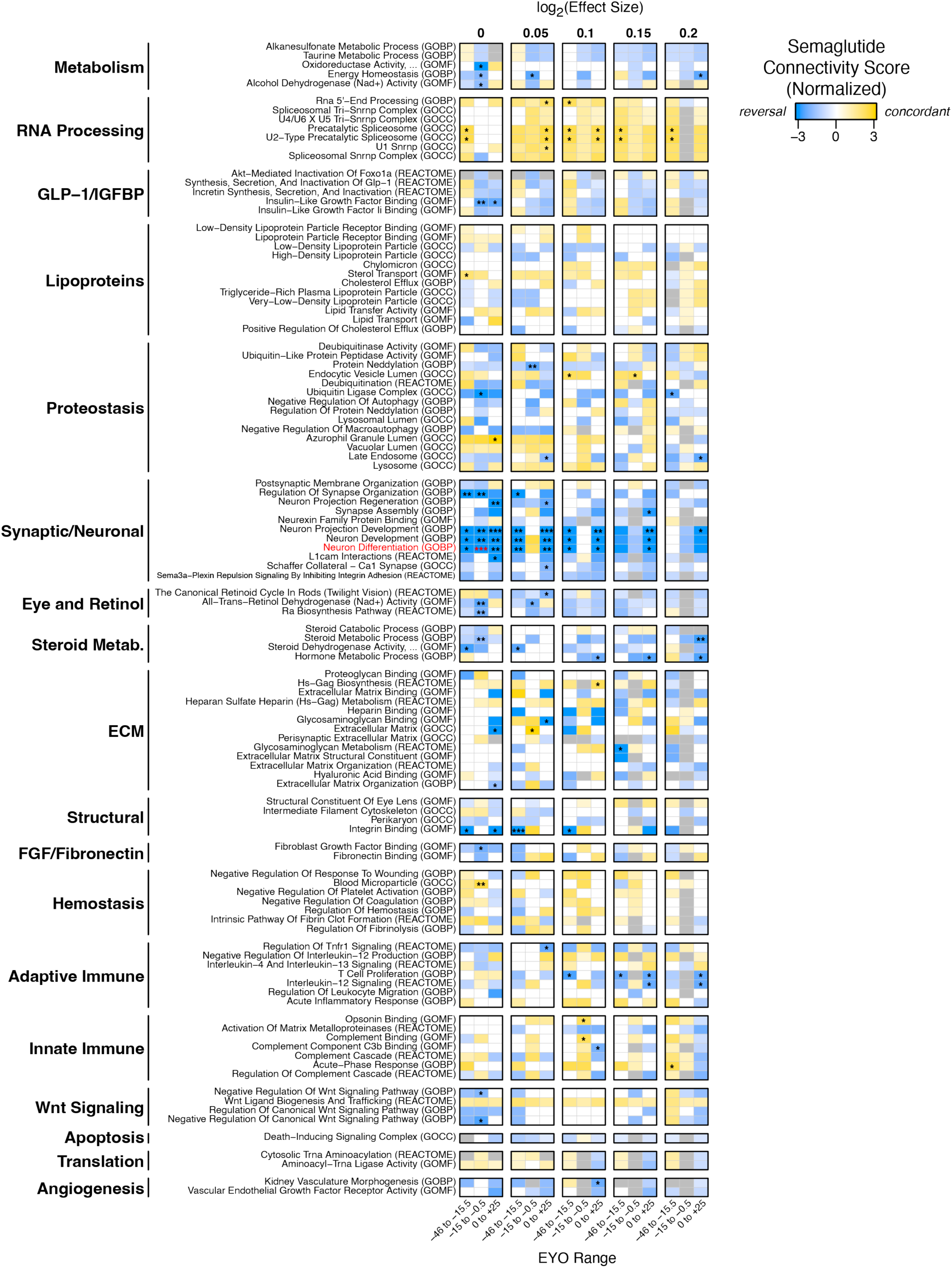
Predicted Effect of Semaglutide on Pathways Altered in ε4 Homozygotes by ε4 Effect Size. The ε4 effect size threshold for connectivity analysis was varied between log2(0) (no threshold) and log2(0.2), including log2(0.1) shown in Figure 6, and the potential effect of semaglutide on each biological pathway at three EYO epochs was evaluated. Gray indicates connectivity could not be calculated due to insufficient number of proteins. Connectivity score is reported after permutation. *p< 0.05; \*\**p*< 0.01; \*\*\**p*< 0.001. Red highlight indicates significance after FDR for the given effect size.

**Supplementary Figure 13.**
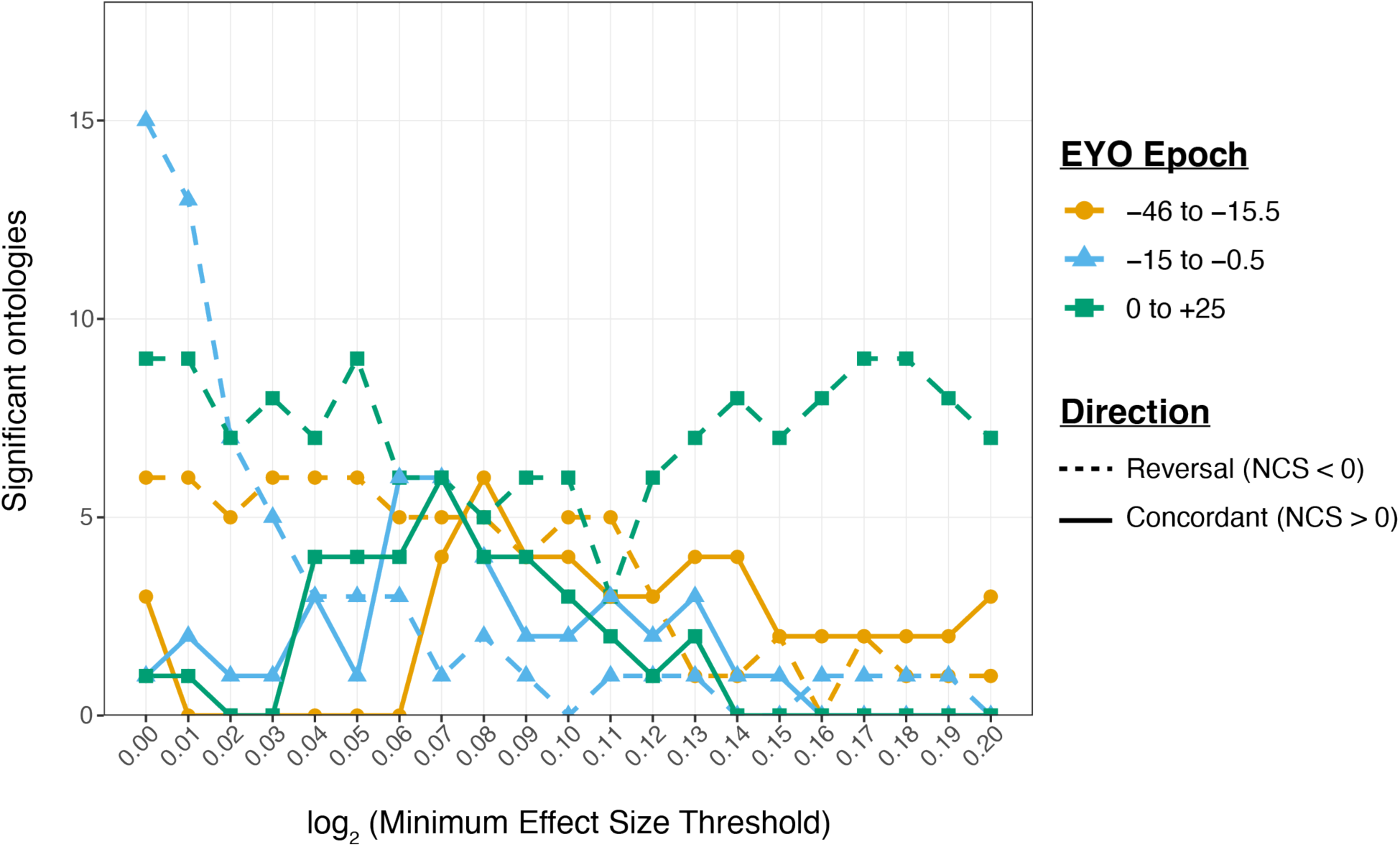
Effect of Effect Size Threshold on Ontology Modulation by Semaglutide. The minimum effect size difference for proteins within ontologies significantly enriched in ε4-affected proteins as shown in Figure 3 was varied between log2(0) and log2(0.2), and connectivity analysis was performed at each threshold to assess the number of ontologies significantly affected by semaglutide. Results are shown for both reversal and concordant semaglutide effects in the three EYO epochs analyzed in Figure 6 and **Supplementary** Figure 12. NCS, normalized connectivity score.

## References

1. C. Bellenguez et al., New insights into the genetic etiology of Alzheimer’s disease and related dementias. Nat Genet 54, 412–436 (2022).

2. A. J. Griswold et al., Increased APOE epsilon4 expression is associated with the difference in Alzheimer’s disease risk from diverse ancestral backgrounds. Alzheimers Dement 17, 1179–1188 (2021).

3. L. A. Farrer et al., Effects of age, sex, and ethnicity on the association between apolipoprotein E genotype and Alzheimer disease. A meta-analysis. APOE and Alzheimer Disease Meta Analysis Consortium. JAMA 278, 1349–1356 (1997).

4. A. Ward et al., Prevalence of apolipoprotein E4 genotype and homozygotes (APOE e4/4) among patients diagnosed with Alzheimer’s disease: a systematic review and meta-analysis. Neuroepidemiology 38, 1–17 (2012).

5. E. H. Corder et al., Gene dose of apolipoprotein E type 4 allele and the risk of Alzheimer’s disease in late onset families. Science 261, 921–923 (1993).

6. A. D. Roses, Apolipoprotein E alleles as risk factors in Alzheimer’s disease. Annu Rev Med 47, 387–400 (1996).

7. A. M. Saunders et al., Association of apolipoprotein E allele epsilon 4 with late-onset familial and sporadic Alzheimer’s disease. Neurology 43, 1467–1472 (1993).

8. A. Shvetcov et al., APOE epsilon4 carriers share immune-related proteomic changes across neurodegenerative diseases. Nature medicine 31, 2590–2601 (2025).

9. J. Fortea et al., Publisher Correction: APOE4 homozygosity represents a distinct genetic form of Alzheimer’s disease. Nature medicine 30, 2093 (2024).

10. D. C. Ryman et al., Symptom onset in autosomal dominant Alzheimer disease: a systematic review and meta-analysis. Neurology 83, 253–260 (2014).

11. J. Fortea et al., Clinical and biomarker changes of Alzheimer’s disease in adults with Down syndrome: a cross-sectional study. Lancet 395, 1988–1997 (2020).

12. E. A. Frick et al., Serum proteomics reveal APOE-epsilon4-dependent and APOE-epsilon4-independent protein signatures in Alzheimer’s disease. Nat Aging 4, 1446–1464 (2024).

13. S. M. Philippi, K. Bp, T. Raj, J. M. Castellano, APOE genotype and brain amyloid are associated with changes in the plasma proteome in elderly subjects without dementia. Ann Clin Transl Neurol 12, 366–382 (2025).

14. K. A. Walker et al., Proteomic analysis of APOEepsilon4 carriers implicates lipid metabolism, complement and lymphocyte signaling in cognitive resilience. Mol Neurodegener 19, 81 (2024).

15. E. C. B. Johnson et al., Cerebrospinal fluid proteomics define the natural history of autosomal dominant Alzheimer’s disease. Nature medicine 29, 1979–1988 (2023).

16. L. Montoliu-Gaya et al., Proteomic analysis of Down syndrome cerebrospinal fluid compared to late-onset and autosomal dominant Alzheimer s disease. Nat Commun 16, 6003 (2025).

17. F. Imam et al., The Global Neurodegeneration Proteomics Consortium: biomarker and drug target discovery for common neurodegenerative diseases and aging. Nature medicine 31, 2556–2566 (2025).

18. S. Afshar et al., Plasma proteomic associations with Alzheimer’s disease endophenotypes. Nat Aging, (2025).

19. K. Garai, P. B. Verghese, B. Baban, D. M. Holtzman, C. Frieden, The binding of apolipoprotein E to oligomers and fibrils of amyloid-beta alters the kinetics of amyloid aggregation. Biochemistry 53, 6323–6331 (2014).

20. K. A. Walker et al., Proteomics analysis of plasma from middle-aged adults identifies protein markers of dementia risk in later life. Science translational medicine 15, eadf5681 (2023).

21. K. A. Walker et al., Large-scale plasma proteomic analysis identifies proteins and pathways associated with dementia risk. Nature Aging 1, 473–489 (2021).

22. J. D. Wright et al., The ARIC (Atherosclerosis Risk In Communities) Study: JACC Focus Seminar 3/8. J Am Coll Cardiol 77, 2939–2959 (2021).

23. T. B. Harris et al., Age, Gene/Environment Susceptibility-Reykjavik Study: multidisciplinary applied phenomics. Am J Epidemiol 165, 1076–1087 (2007).

24. J. V. Lindbohm et al., Plasma proteins, cognitive decline, and 20-year risk of dementia in the Whitehall II and Atherosclerosis Risk in Communities studies. Alzheimers Dement 18, 612–624 (2022).

25. Y. Guo et al., Plasma proteomic profiles predict future dementia in healthy adults. Nat Aging 4, 247–260 (2024).

26. D. A. Bennett et al., Religious Orders Study and Rush Memory and Aging Project. Journal of Alzheimer’s disease : JAD 64, S161–S189 (2018).

27. M. R. Duggan et al., The Dementia SomaSignal Test (dSST): A plasma proteomic predictor of 20-year dementia risk. Alzheimers Dement 21, e14549 (2025).

28. R. Haque et al., A protein panel in cerebrospinal fluid for diagnostic and predictive assessment of Alzheimer’s disease. Science translational medicine 15, eadg4122 (2023).

29. D. J. Drucker, GLP-1-based therapies for diabetes, obesity and beyond. Nat Rev Drug Discov 24, 631–650 (2025).

30. L. Maretty et al., Proteomic changes upon treatment with semaglutide in individuals with obesity. Nature medicine 31, 267–277 (2025).

31. Q. Zhang et al., Comparative analysis and expression of CLUL1, a cone photoreceptor-specific gene. Invest Ophthalmol Vis Sci 44, 4542–4549 (2003).

32. K. Wakasugi et al., A human aminoacyl-tRNA synthetase as a regulator of angiogenesis. Proc Natl Acad Sci U S A 99, 173–177 (2002).

33. R. J. Lumpkin, R. W. Baker, A. E. Leschziner, E. A. Komives, Structure and dynamics of the ASB9 CUL-RING E3 Ligase. Nat Commun 11, 2866 (2020).

34. Y. Huang, Abeta-independent roles of apolipoprotein E4 in the pathogenesis of Alzheimer’s disease. Trends Mol Med 16, 287–294 (2010).

35. E. Schmukler et al., Altered mitochondrial dynamics and function in APOE4-expressing astrocytes. Cell death & disease 11, 578 (2020).

36. S. Lee et al., APOE modulates microglial immunometabolism in response to age, amyloid pathology, and inflammatory challenge. Cell Rep 42, 112196 (2023).

37. H. Y. Sohn et al., ApoE4 attenuates autophagy via FoxO3a repression in the brain. Sci Rep 11, 17604 (2021).

38. S. Simonovitch, E. Schmukler, E. Masliah, R. Pinkas-Kramarski, D. M. Michaelson, The Effects of APOE4 on Mitochondrial Dynamics and Proteins in vivo. Journal of Alzheimer’s disease : JAD 70, 861–875 (2019).

39. A. L. Orr et al., Neuronal Apolipoprotein E4 Expression Results in Proteome-Wide Alterations and Compromises Bioenergetic Capacity by Disrupting Mitochondrial Function. Journal of Alzheimer’s disease : JAD 68, 991–1011 (2019).

40. T. Nakamura, A. Watanabe, T. Fujino, T. Hosono, M. Michikawa, Apolipoprotein E4 (1-272) fragment is associated with mitochondrial proteins and affects mitochondrial function in neuronal cells. Mol Neurodegener 4, 35 (2009).

41. A. Suzuki et al., Astrocyte-neuron lactate transport is required for long-term memory formation. Cell 144, 810–823 (2011).

42. E. M. Reiman et al., Declining brain activity in cognitively normal apolipoprotein E epsilon 4 heterozygotes: A foundation for using positron emission tomography to efficiently test treatments to prevent Alzheimer’s disease. Proc Natl Acad Sci U S A 98, 3334–3339 (2001).

43. E. M. Reiman et al., Functional brain abnormalities in young adults at genetic risk for late-onset Alzheimer’s dementia. Proc Natl Acad Sci U S A 101, 284–289 (2004).

44. E. B. Dammer et al., Proteomic analysis of Alzheimer’s disease cerebrospinal fluid reveals alterations associated with APOE epsilon4 and atomoxetine treatment. Science translational medicine 16, eadn3504 (2024).

45. S. Guo et al., Phosphorylation of serine 256 by protein kinase B disrupts transactivation by FKHR and mediates effects of insulin on insulin-like growth factor-binding protein-1 promoter activity through a conserved insulin response sequence. J Biol Chem 274, 17184–17192 (1999).

46. I. C. Lee, K. L. Chiang, Clinical Diagnosis and Treatment of Leigh Syndrome Based on SURF1: Genotype and Phenotype. Antioxidants (Basel) 10, (2021).

47. P. Agarwal et al., Association of Mediterranean-DASH Intervention for Neurodegenerative Delay and Mediterranean Diets With Alzheimer Disease Pathology. Neurology 100, e2259–e2268 (2023).

48. M. Fekete et al., The role of the Mediterranean diet in reducing the risk of cognitive impairement, dementia, and Alzheimer’s disease: a meta-analysis. Geroscience 47, 3111–3130 (2025).

49. Y. Liu et al., Interplay of genetic predisposition, plasma metabolome and Mediterranean diet in dementia risk and cognitive function. Nature medicine 31, 3790–3800 (2025).

50. C. Luna-Marco et al., Effects of GLP-1 receptor agonists on mitochondrial function, inflammatory markers and leukocyte-endothelium interactions in type 2 diabetes. Redox Biol 66, 102849 (2023).

51. A. Seminer et al., Cardioprotective Glucose-Lowering Agents and Dementia Risk: A Systematic Review and Meta-Analysis. JAMA Neurol 82, 450–460 (2025).

52. E. B. Dammer et al., Polyubiquitin linkage profiles in three models of proteolytic stress suggest the etiology of Alzheimer disease. J Biol Chem 286, 10457–10465 (2011).

53. X. He, A. Zhu, J. Feng, X. Wang, Role of neddylation in neurological development and diseases. Biotechnol Appl Biochem 69, 330–341 (2022).

54. S. Zhang, Q. Yu, Z. Li, Y. Zhao, Y. Sun, Protein neddylation and its role in health and diseases. Signal Transduct Target Ther 9, 85 (2024).

55. J. L. Robinson et al., Neurodegenerative disease concomitant proteinopathies are prevalent, age-related and APOE4-associated. Brain : a journal of neurology 141, 2181–2193 (2018).

56. Y. Jin et al., APOE4 exacerbates alpha-synuclein seeding activity and contributes to neurotoxicity in Alzheimer’s disease with Lewy body pathology. Acta Neuropathol 143, 641–662 (2022).

57. B. Bai et al., U1 small nuclear ribonucleoprotein complex and RNA splicing alterations in Alzheimer’s disease. Proc Natl Acad Sci U S A 110, 16562–16567 (2013).

58. C. M. Hales et al., Aggregates of small nuclear ribonucleic acids (snRNAs) in Alzheimer’s disease. Brain Pathol 24, 344–351 (2014).

59. E. C. B. Johnson et al., Deep proteomic network analysis of Alzheimer’s disease brain reveals alterations in RNA binding proteins and RNA splicing associated with disease. Mol Neurodegener 13, 52 (2018).

60. H. Vakifahmetoglu-Norberg, H. G. Xia, J. Yuan, Pharmacologic agents targeting autophagy. J Clin Invest 125, 5–13 (2015).

61. Y. T. Lin et al., APOE4 Causes Widespread Molecular and Cellular Alterations Associated with Alzheimer’s Disease Phenotypes in Human iPSC-Derived Brain Cell Types. Neuron 98, 1141–1154 e1147 (2018).

62. W. Khan et al., A Multi-Cohort Study of ApoE varepsilon4 and Amyloid-beta Effects on the Hippocampus in Alzheimer’s Disease. Journal of Alzheimer’s disease : JAD 56, 1159–1174 (2017).

63. R. J. Piers, Structural brain volume differences between cognitively intact ApoE4 carriers and non-carriers across the lifespan. Neural Regen Res 13, 1309–1312 (2018).

64. A. Snellman et al., SV2A PET shows hippocampal synaptic loss in cognitively unimpaired APOE epsilon4/epsilon4 homozygotes. Alzheimers Dement 20, 8802–8813 (2024).

65. R. Najm, E. A. Jones, Y. Huang, Apolipoprotein E4, inhibitory network dysfunction, and Alzheimer’s disease. Mol Neurodegener 14, 24 (2019).

66. E. B. Dammer et al., Multi-platform proteomic analysis of Alzheimer’s disease cerebrospinal fluid and plasma reveals network biomarkers associated with proteostasis and the matrisome. Alzheimers Res Ther 14, 174 (2022).

67. A. Lleo et al., Changes in Synaptic Proteins Precede Neurodegeneration Markers in Preclinical Alzheimer’s Disease Cerebrospinal Fluid. Mol Cell Proteomics 18, 546–560 (2019).

68. M. P. Vitek, C. M. Brown, C. A. Colton, APOE genotype-specific differences in the innate immune response. Neurobiol Aging 30, 1350–1360 (2009).

69. S. Parhizkar, D. M. Holtzman, APOE mediated neuroinflammation and neurodegeneration in Alzheimer’s disease. Semin Immunol 59, 101594 (2022).

70. Y. Shi et al., ApoE4 markedly exacerbates tau-mediated neurodegeneration in a mouse model of tauopathy. Nature 549, 523–527 (2017).

71. A. F. Batista, K. A. Khan, M. T. Papavergi, C. A. Lemere, The Importance of Complement-Mediated Immune Signaling in Alzheimer’s Disease Pathogenesis. Int J Mol Sci 25, (2024).

72. S. Hong et al., Complement and microglia mediate early synapse loss in Alzheimer mouse models. Science 352, 712–716 (2016).

73. J. A. Zimmer et al., Amyloid-Related Imaging Abnormalities With Donanemab in Early Symptomatic Alzheimer Disease: Secondary Analysis of the TRAILBLAZER-ALZ and ALZ 2 Randomized Clinical Trials. JAMA Neurol 82, 461–469 (2025).

74. P. Bathini, I. Navani, C. A. Lemere, Complement-mediated vascular inflammation contributes to anti-amyloid mAb-induced ARIA in mice. Alzheimers Dement 21, (2025).

75. E. van Exel et al., Effect of APOE epsilon4 allele on survival and fertility in an adverse environment. PLoS One 12, e0179497 (2017).

76. H. Yaribeygi, M. Maleki, T. Jamialahmadi, A. Sahebkar, Anti-inflammatory benefits of semaglutide: State of the art. J Clin Transl Endocrinol 36, 100340 (2024).

77. N. Basisty et al., A proteomic atlas of senescence-associated secretomes for aging biomarker development. PLoS biology 18, e3000599 (2020).

78. J. Reyes, G. S. Yap, Emerging Roles of Growth Differentiation Factor 15 in Immunoregulation and Pathogenesis. J Immunol 210, 5–11 (2023).

79. Y. Fujita, Y. Taniguchi, S. Shinkai, M. Tanaka, M. Ito, Secreted growth differentiation factor 15 as a potential biomarker for mitochondrial dysfunctions in aging and age-related disorders. Geriatr Gerontol Int 16 **Suppl 1**, 17–29 (2016).

80. T. Kuboki, S. Kidoaki, Surface-elastic hydrogels delay senescence via the modulation of redox homeostasis and cytoskeletal tension. Sci Rep 15, 20460 (2025).

81. L. Jakel et al., Altered brain expression and cerebrospinal fluid levels of TIMP4 in cerebral amyloid angiopathy. Acta Neuropathol Commun 12, 103 (2024).

82. A. M. Wojtas et al., Proteomic changes in the human cerebrovasculature in Alzheimer’s disease and related tauopathies linked to peripheral biomarkers in plasma and cerebrospinal fluid. Alzheimers Dement 20, 4043–4065 (2024).

83. E. C. B. Johnson et al., Large-scale deep multi-layer analysis of Alzheimer’s disease brain reveals strong proteomic disease-related changes not observed at the RNA level. Nature neuroscience 25, 213–225 (2022).

84. G. J. McKay et al., Evidence of association of APOE with age-related macular degeneration: a pooled analysis of 15 studies. Hum Mutat 32, 1407–1416 (2011).

85. K. L. Rasmussen, A. Tybjaerg-Hansen, B. G. Nordestgaard, R. Frikke-Schmidt, Associations of Alzheimer Disease-Protective APOE Variants With Age-Related Macular Degeneration. JAMA Ophthalmol 141, 13–21 (2023).

86. W. Yi, D. Lv, Y. Sun, J. Mu, X. Lu, Role of APOE in glaucoma. Biochem Biophys Res Commun 694, 149414 (2024).

87. M. A. Margeta et al., Apolipoprotein E4 impairs the response of neurodegenerative retinal microglia and prevents neuronal loss in glaucoma. Immunity 55, 1627–1644 e1627 (2022).

88. H. N. Yassine et al., APOE-Targeted Therapeutics for Alzheimer’s Disease. J Neurosci 45, (2025).

89. J. M. Vance et al., Report of the APOE4 National Institute on Aging/Alzheimer Disease Sequencing Project Consortium Working Group: Reducing APOE4 in Carriers is a Therapeutic Goal for Alzheimer’s Disease. Ann Neurol 95, 625–634 (2024).

90. J. L. Cummings et al., Efficacy and safety of oral semaglutide 14 mg (flexible dose) in early-stage symptomatic Alzheimer’s disease (evoke and evoke+): two phase 3, randomised, placebo-controlled trials. Lancet 407, 2167–2179 (2026).

91. P. Edison et al., Liraglutide in mild to moderate Alzheimer’s disease: a phase 2b clinical trial. Nature medicine 32, 353–361 (2026).

92. J. R. Sims et al., Donanemab in Early Symptomatic Alzheimer Disease: The TRAILBLAZER-ALZ 2 Randomized Clinical Trial. JAMA 330, 512–527 (2023).

93. C. H. van Dyck et al., Lecanemab in Early Alzheimer’s Disease. N Engl J Med, (2022).

94. S. Afshar et al., Plasma proteomic associations with Alzheimer’s disease endophenotypes. Nature Aging, (2025).

95. P. Langfelder, S. Horvath, WGCNA: an R package for weighted correlation network analysis. BMC Bioinformatics 9, 559 (2008).

96. E. C. B. Johnson et al., Large-scale deep multi-layer analysis of Alzheimer’s disease brain reveals strong proteomic disease-related changes not observed at the RNA level. Nature Neuroscience 25, 213–225 (2022).

97. J. Reimand et al., Pathway enrichment analysis and visualization of omics data using g:Profiler, GSEA, Cytoscape and EnrichmentMap. Nat Protoc 14, 482–517 (2019).

98. C. Muth, Z. Oravecz, J. Gabry, User-friendly Bayesian regression modeling: A tutorial with rstanarm and shinystan. The Quantitative Methods for Psychology 14, 99–119 (2018).

99. O. Preische et al., Serum neurofilament dynamics predicts neurodegeneration and clinical progression in presymptomatic Alzheimer’s disease. Nature medicine 25, 277–283 (2019).

100. J. Gabry, Goodrich, B., Prior Distributions for rstanarm Models. (2020).

101. L. Coenen, B. Lehallier, H. E. de Vries, J. Middeldorp, Markers of aging: Unsupervised integrated analyses of the human plasma proteome. Front Aging 4, 1112109 (2023).

102. J. Lamb et al., The Connectivity Map: using gene-expression signatures to connect small molecules, genes, and disease. Science 313, 1929–1935 (2006).

103. A. Subramanian et al., A Next Generation Connectivity Map: L1000 Platform and the First 1,000,000 Profiles. Cell 171, 1437–1452 e1417 (2017).

104. A. Subramanian et al., Gene set enrichment analysis: a knowledge-based approach for interpreting genome-wide expression profiles. Proc Natl Acad Sci U S A 102, 15545–15550 (2005).

