## Supplementary material for "Plasma Proteomic Analysis of *APOE* ε4 Homozygotes Identifies Preclinical Alzheimer’s Disease Alterations Potentially Modulated by Semaglutide": Dammer et al. Supplementary Information

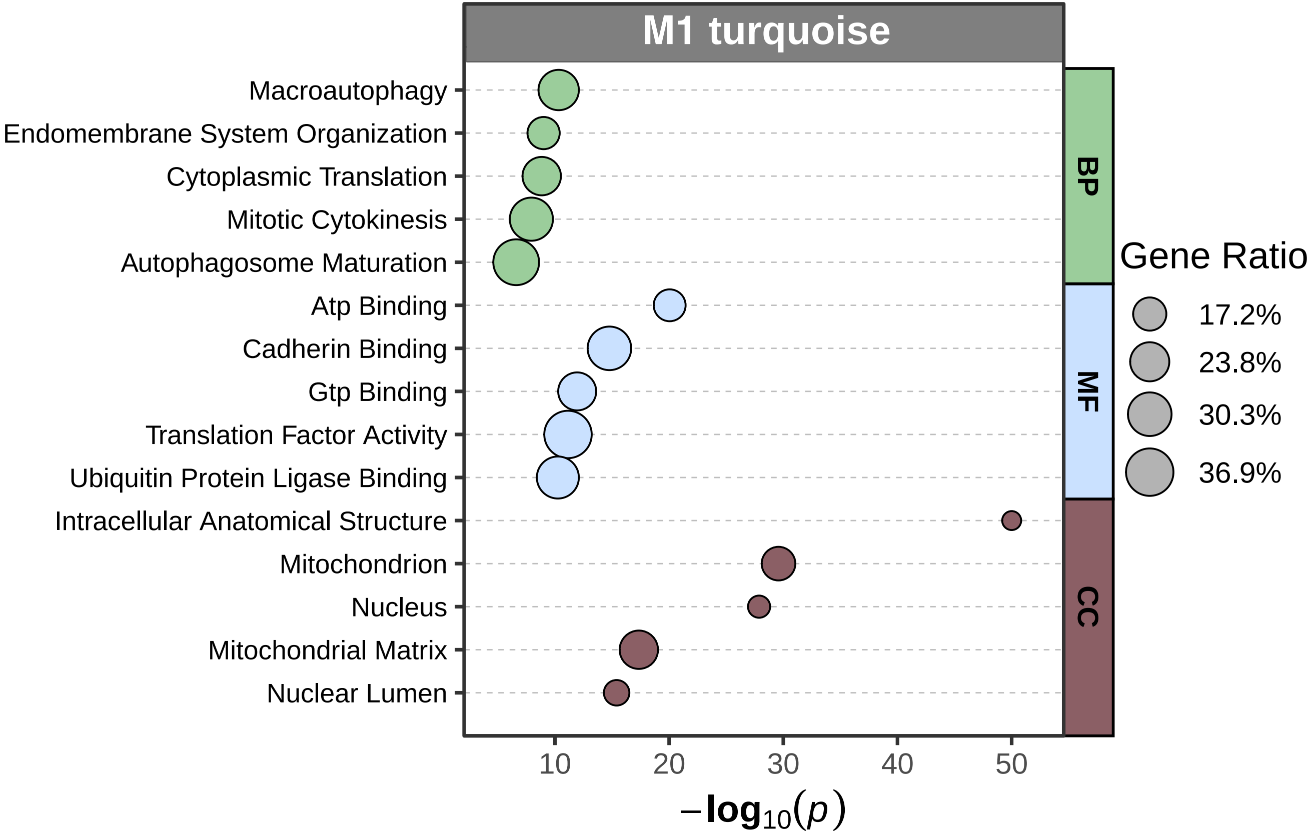
M1 turquoise
Macroautophagy
Endomembrane System Organization
Cytoplasmic Translation
Mitotic Cytokinesis
Autophagosome Maturation
Atp Binding
Cadherin Binding
Gtp Binding
Translation Factor Activity
Ubiquitin Protein Ligase Binding
Intracellular Anatomical Structure
Mitochondrion
Nucleus
Mitochondrial Matrix
Nuclear Lumen
BP
MF
CC
Gene Ratio
17.2%
23.8%
30.3%
36.9%
10
20
30
40
50
$-\log_{10}(p)$

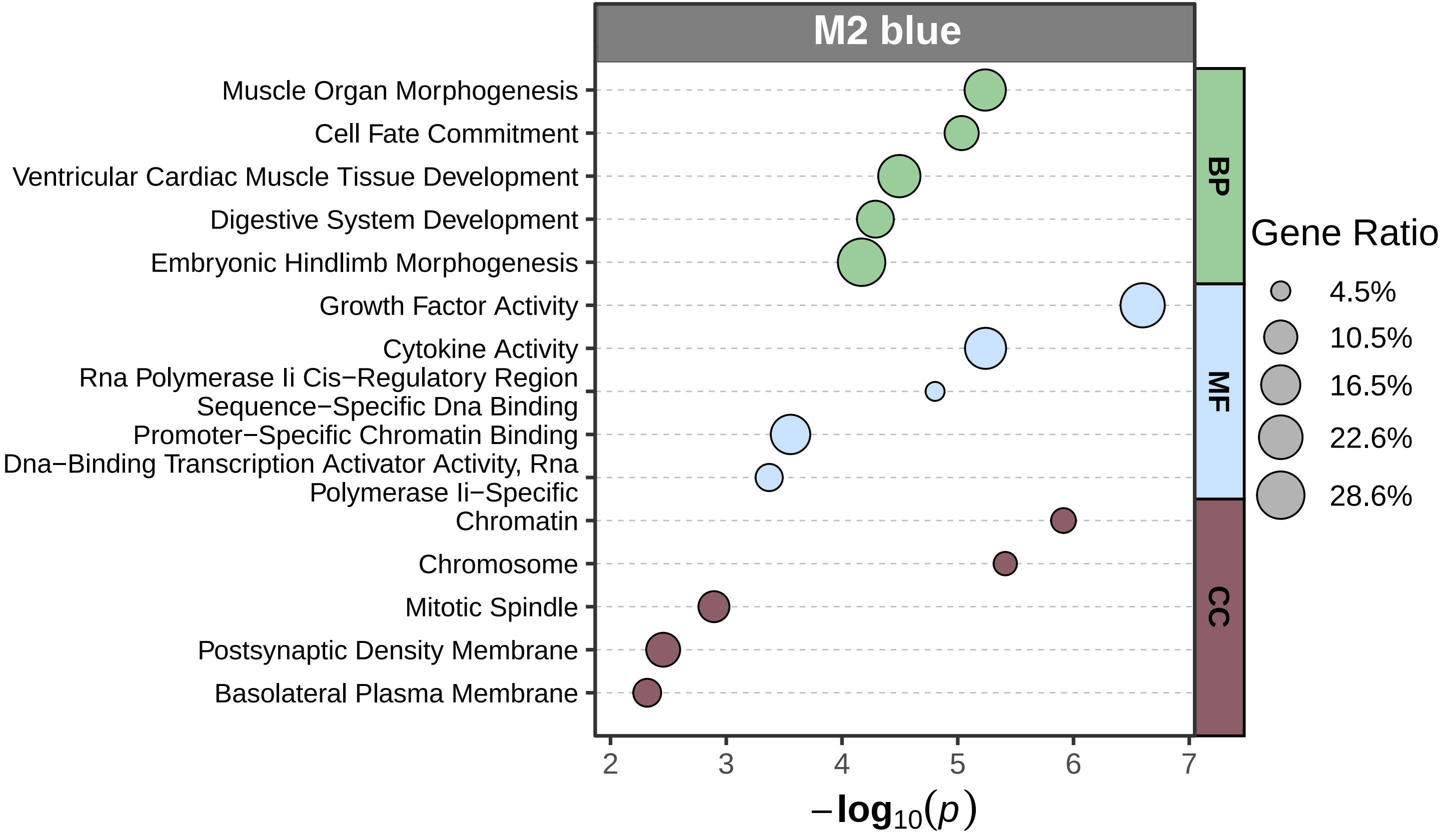
M2 blue
Muscle Organ Morphogenesis
Cell Fate Commitment
Ventricular Cardiac Muscle Tissue Development
Digestive System Development
Embryonic Hindlimb Morphogenesis
Growth Factor Activity
Cytokine Activity
Rna Polymerase Ii Cis−Regulatory Region Sequence−Specific Dna Binding
Promoter−Specific Chromatin Binding
Dna−Binding Transcription Activator Activity, Rna Polymerase Ii−Specific
Chromatin
Chromosome
Mitotic Spindle
Postsynaptic Density Membrane
Basolateral Plasma Membrane
BP
MF
CC
Gene Ratio
4.5%
10.5%
16.5%
22.6%
28.6%
2
3
4
5
6
7
$-\log_{10}(p)$

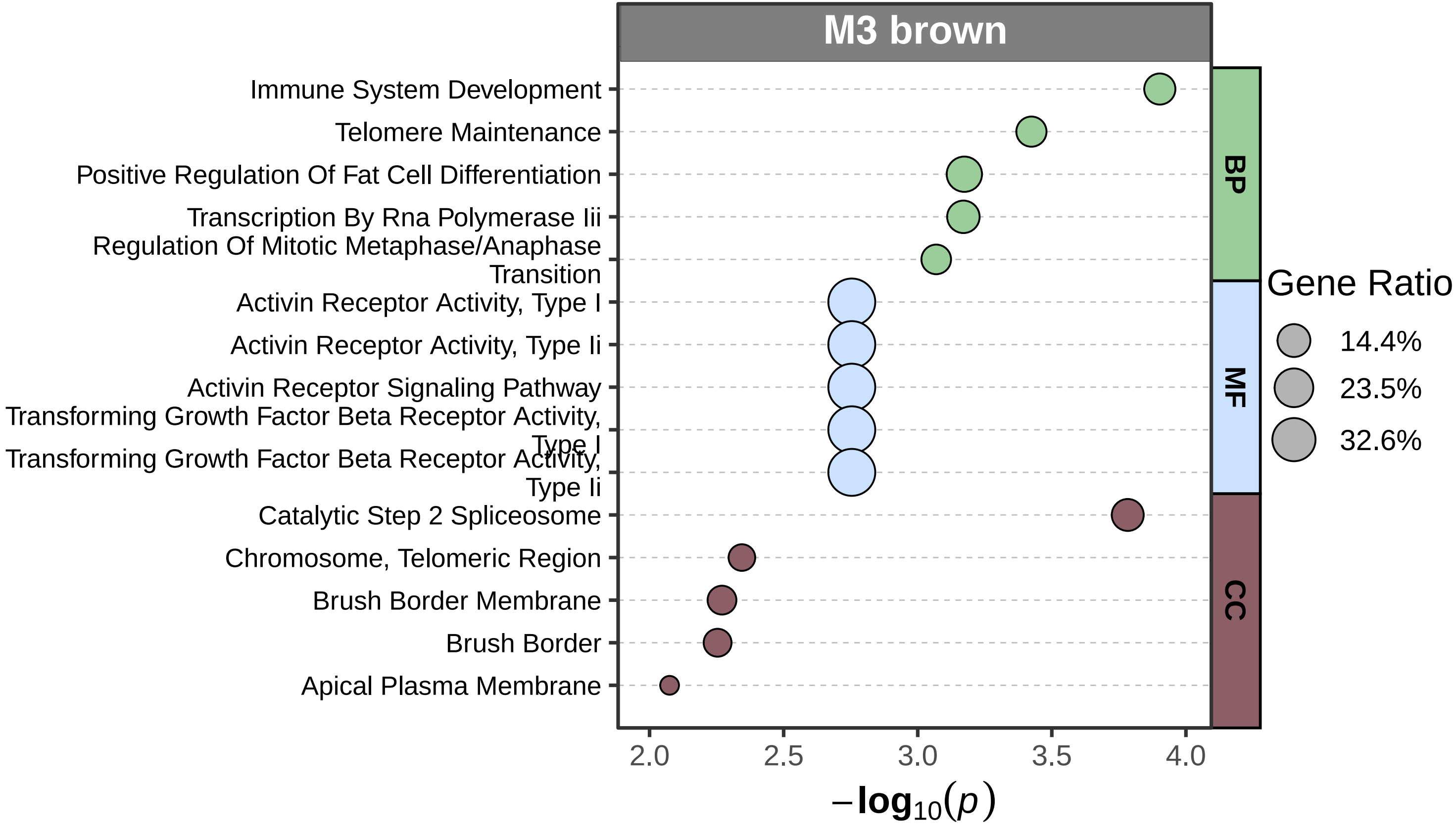
M3 brown
Immune System Development
Telomere Maintenance
Positive Regulation Of Fat Cell Differentiation
Transcription By Rna Polymerase Iii
Regulation Of Mitotic Metaphase/Anaphase Transition
Activin Receptor Activity, Type I
Activin Receptor Activity, Type Ii
Activin Receptor Signaling Pathway
Transforming Growth Factor Beta Receptor Activity, Type I
Transforming Growth Factor Beta Receptor Activity, Type Ii
Catalytic Step 2 Spliceosome
Chromosome, Telomeric Region
Brush Border Membrane
Brush Border
Apical Plasma Membrane
BP
MF
CC
Gene Ratio
14.4%
23.5%
32.6%
2.0
2.5
3.0
3.5
4.0
$-\log_{10}(p)$

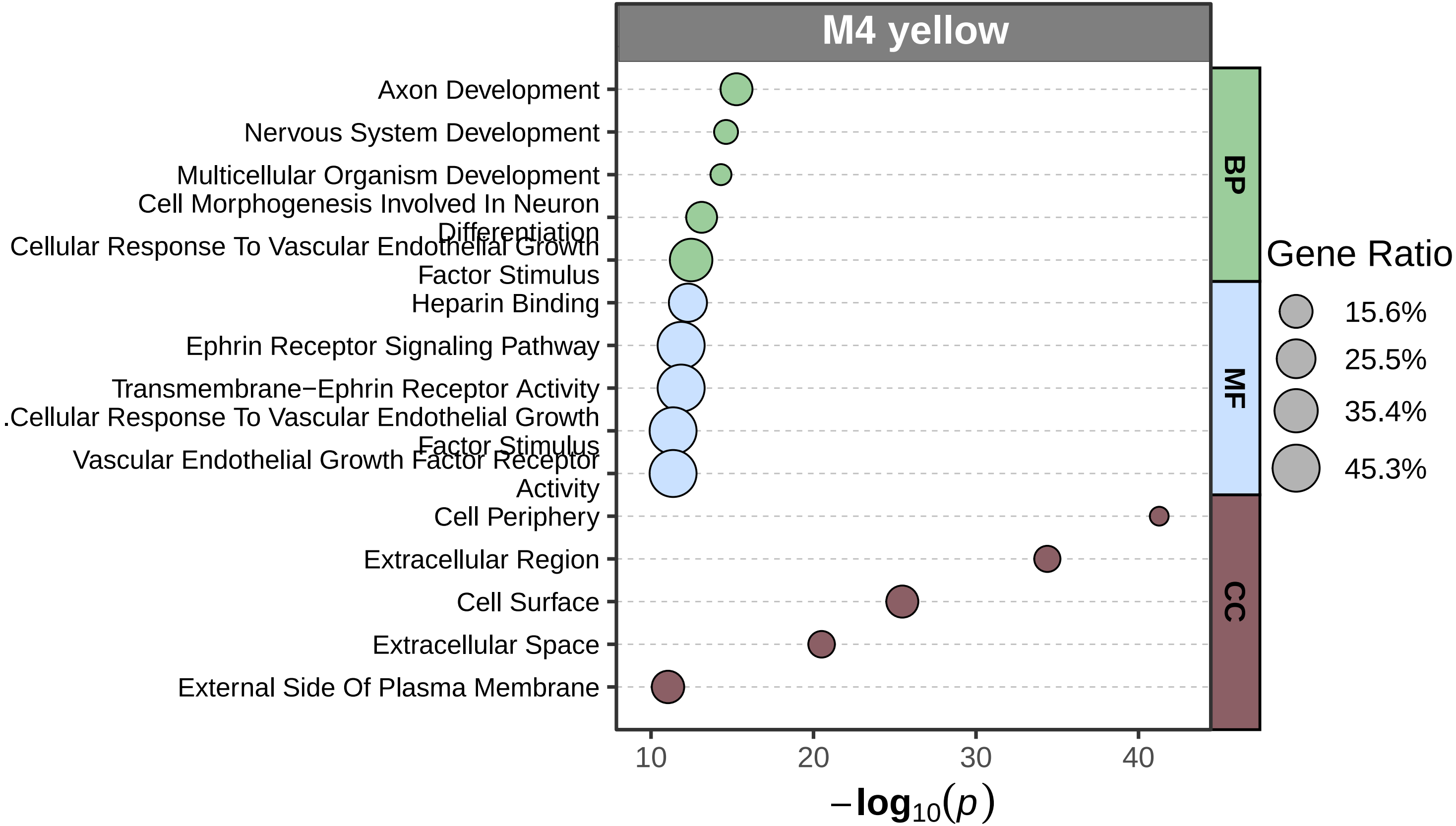

M4 yellow
Axon Development
Nervous System Development
Multicellular Organism Development
Cell Morphogenesis Involved In Neuron Differentiation
Cellular Response To Vascular Endothelial Growth Factor Stimulus
Heparin Binding
Ephrin Receptor Signaling Pathway
Transmembrane−Ephrin Receptor Activity
.Cellular Response To Vascular Endothelial Growth Factor Stimulus
Vascular Endothelial Growth Factor Receptor Activity
Cell Periphery
Extracellular Region
Cell Surface
Extracellular Space
External Side Of Plasma Membrane
BP
MF
CC
Gene Ratio
15.6%
25.5%
35.4%
45.3%
10
20
30
40
$-\log_{10}(p)$

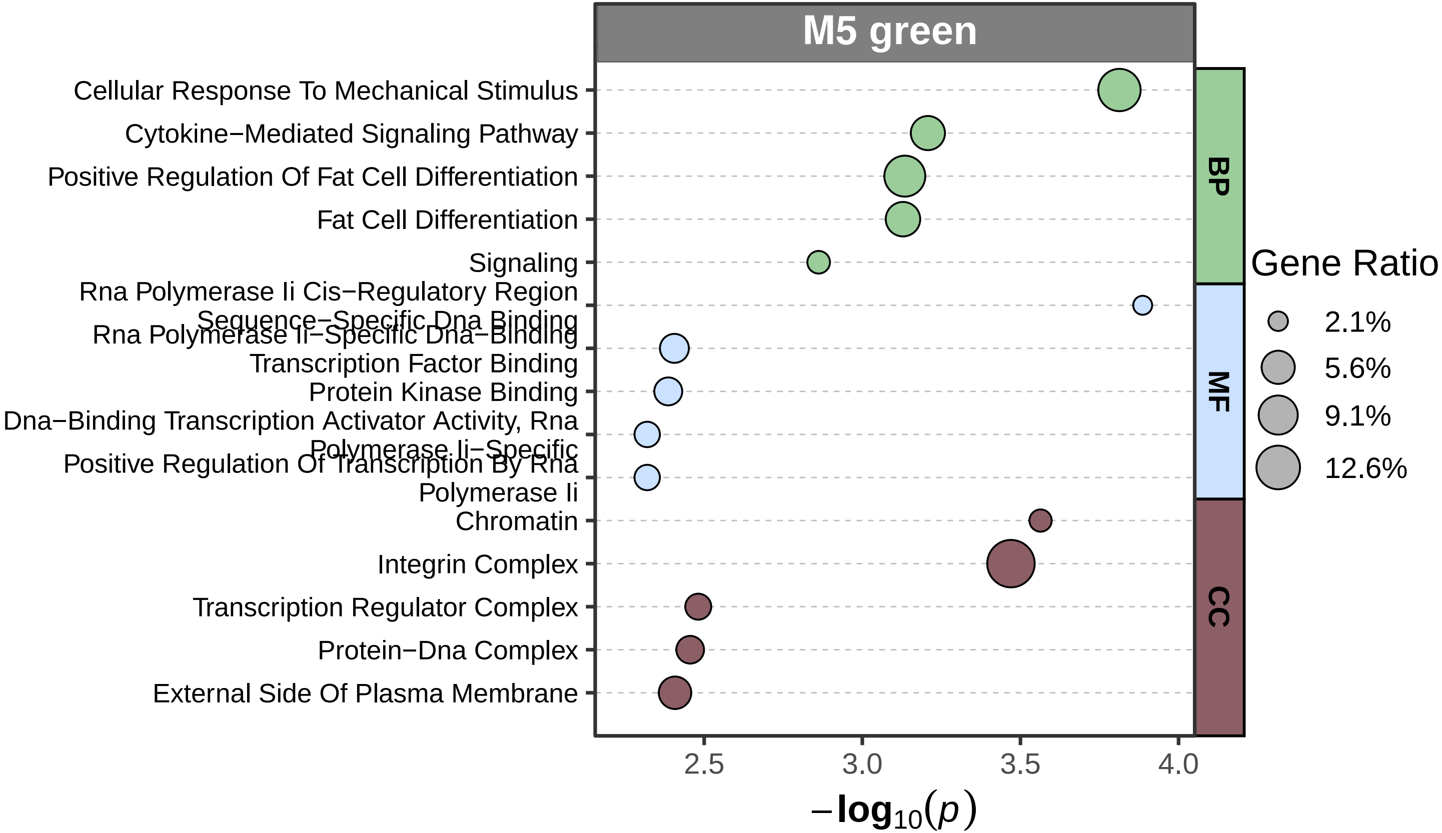
M5 green
Cellular Response To Mechanical Stimulus
Cytokine−Mediated Signaling Pathway
Positive Regulation Of Fat Cell Differentiation
Fat Cell Differentiation
Signaling
Rna Polymerase Ii Cis−Regulatory Region Sequence−Specific Dna Binding
Rna Polymerase Ii−Specific Dna−Binding Transcription Factor Binding
Protein Kinase Binding
Dna−Binding Transcription Activator Activity, Rna Polymerase Ii−Specific
Positive Regulation Of Transcription By Rna Polymerase Ii
Chromatin
Integrin Complex
Transcription Regulator Complex
Protein−Dna Complex
External Side Of Plasma Membrane
BP
MF
CC
Gene Ratio
2.1%
5.6%
9.1%
12.6%
2.5
3.0
3.5
4.0
$-\log_{10}(p)$

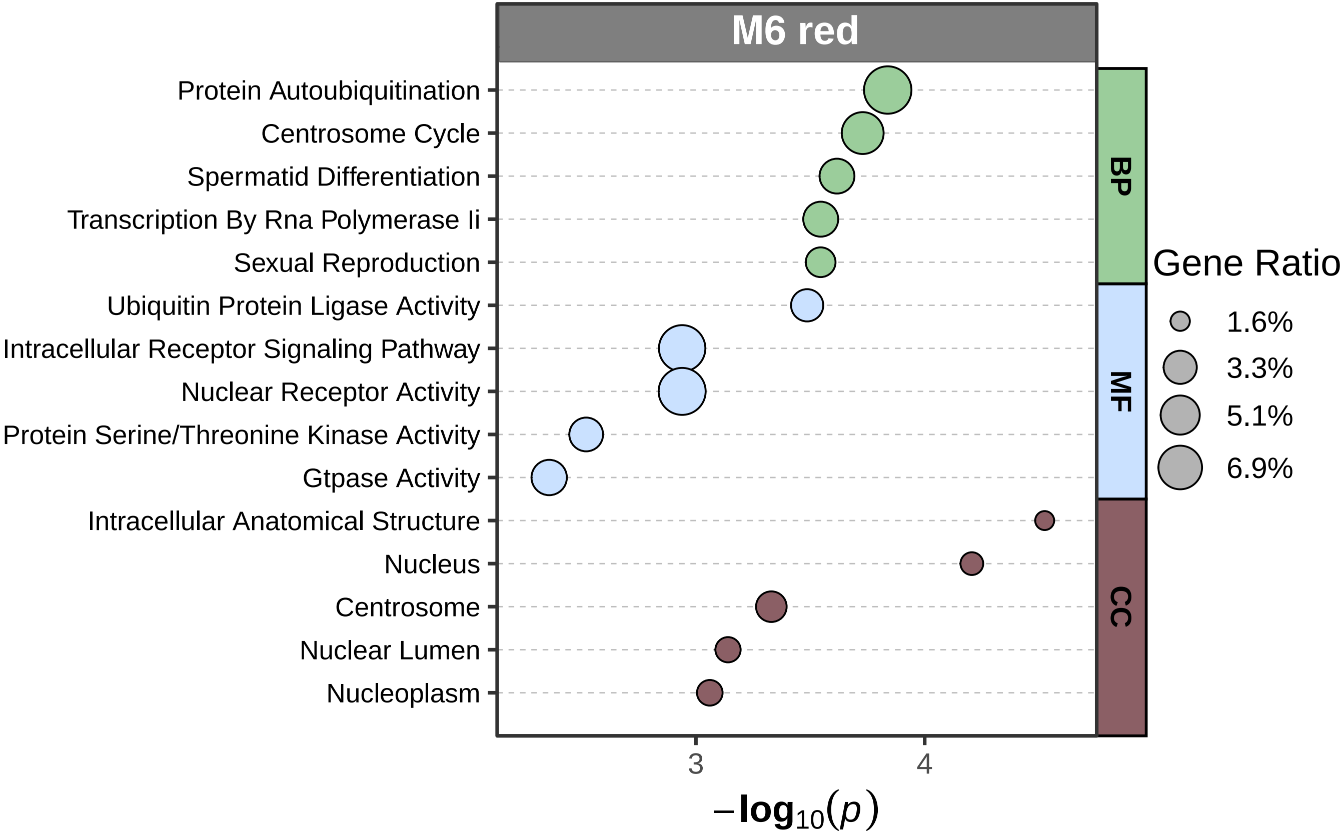
M6 red
Protein Autoubiquitination
Centrosome Cycle
Spermatid Differentiation
Transcription By Rna Polymerase Ii
Sexual Reproduction
Ubiquitin Protein Ligase Activity
Intracellular Receptor Signaling Pathway
Nuclear Receptor Activity
Protein Serine/Threonine Kinase Activity
Gtpase Activity
Intracellular Anatomical Structure
Nucleus
Centrosome
Nuclear Lumen
Nucleoplasm
BP
MF
CC
3
4
$-\log_{10}(p)$
Gene Ratio
1.6%
3.3%
5.1%
6.9%

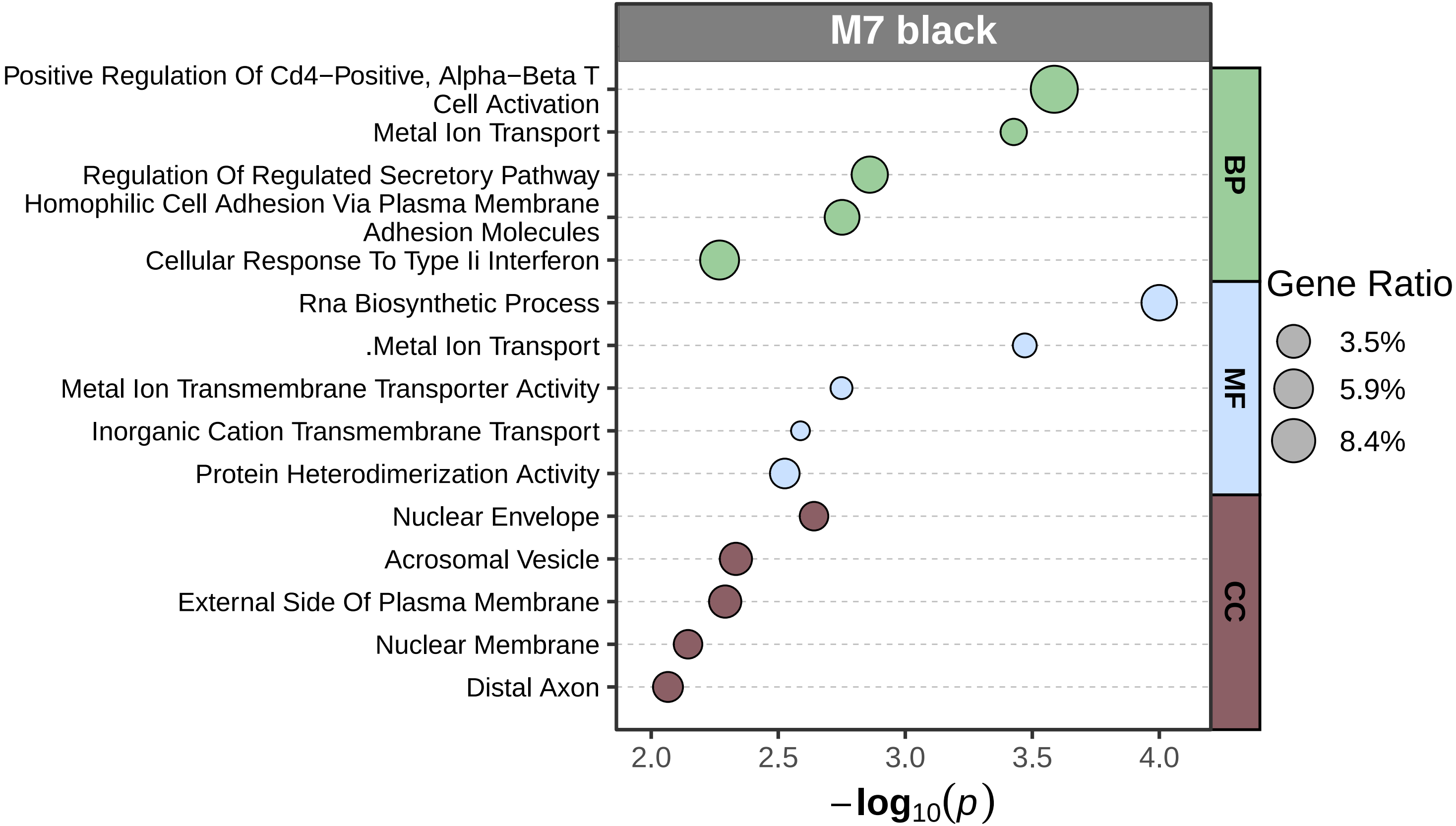

M7 black
Positive Regulation Of Cd4−Positive, Alpha−Beta T Cell Activation
Metal Ion Transport
Regulation Of Regulated Secretory Pathway
Homophilic Cell Adhesion Via Plasma Membrane Adhesion Molecules
Cellular Response To Type Ii Interferon
Rna Biosynthetic Process
.Metal Ion Transport
Metal Ion Transmembrane Transporter Activity
Inorganic Cation Transmembrane Transport
Protein Heterodimerization Activity
Nuclear Envelope
Acrosomal Vesicle
External Side Of Plasma Membrane
Nuclear Membrane
Distal Axon
BP
MF
CC
Gene Ratio
3.5%
5.9%
8.4%
2.0
2.5
3.0
3.5
4.0
$-\log_{10}(p)$

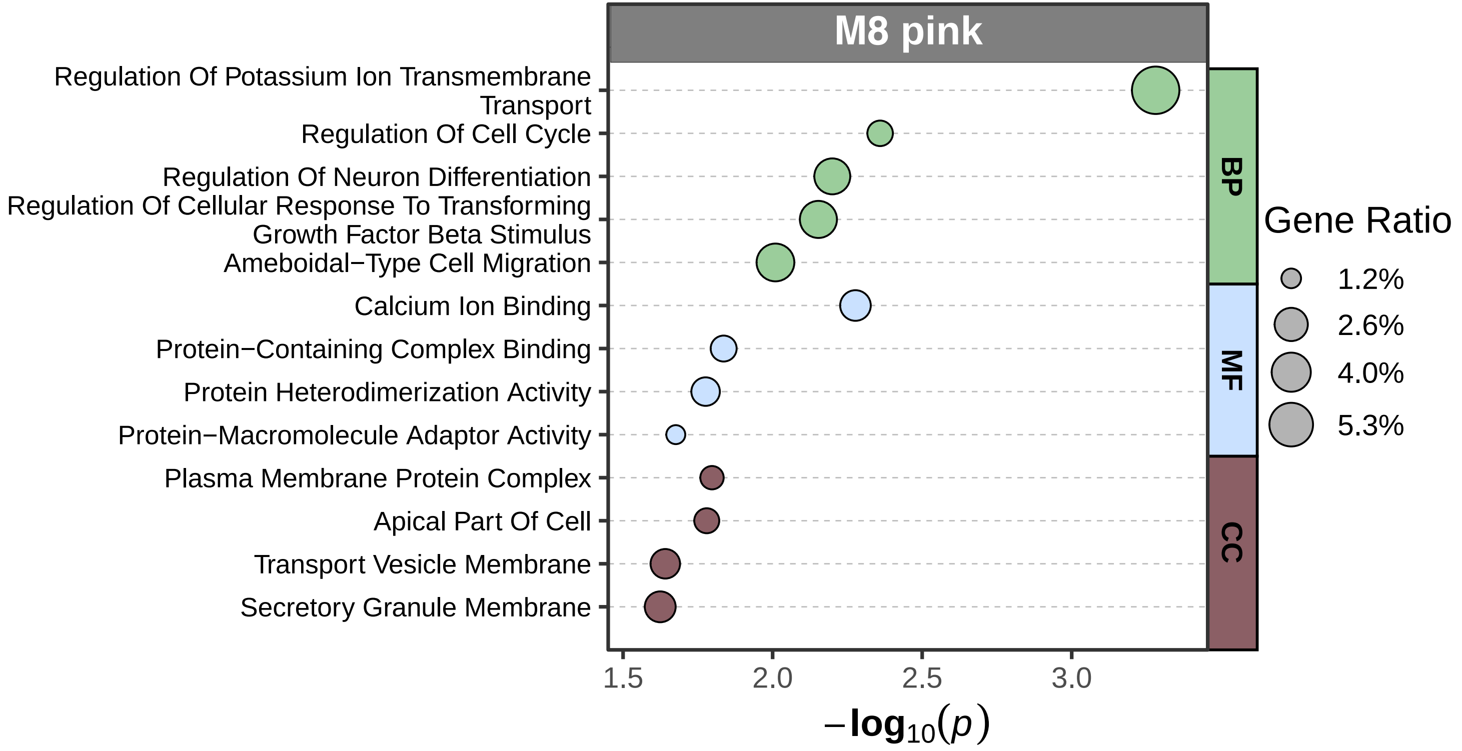

M8 pink
Regulation Of Potassium Ion Transmembrane Transport
Regulation Of Cell Cycle
Regulation Of Neuron Differentiation
Regulation Of Cellular Response To Transforming Growth Factor Beta Stimulus
Ameboidal−Type Cell Migration
Calcium Ion Binding
Protein−Containing Complex Binding
Protein Heterodimerization Activity
Protein−Macromolecule Adaptor Activity
Plasma Membrane Protein Complex
Apical Part Of Cell
Transport Vesicle Membrane
Secretory Granule Membrane
BP
MF
CC
Gene Ratio
1.2%
2.6%
4.0%
5.3%
1.5
2.0
2.5
3.0
$-\log_{10}(p)$

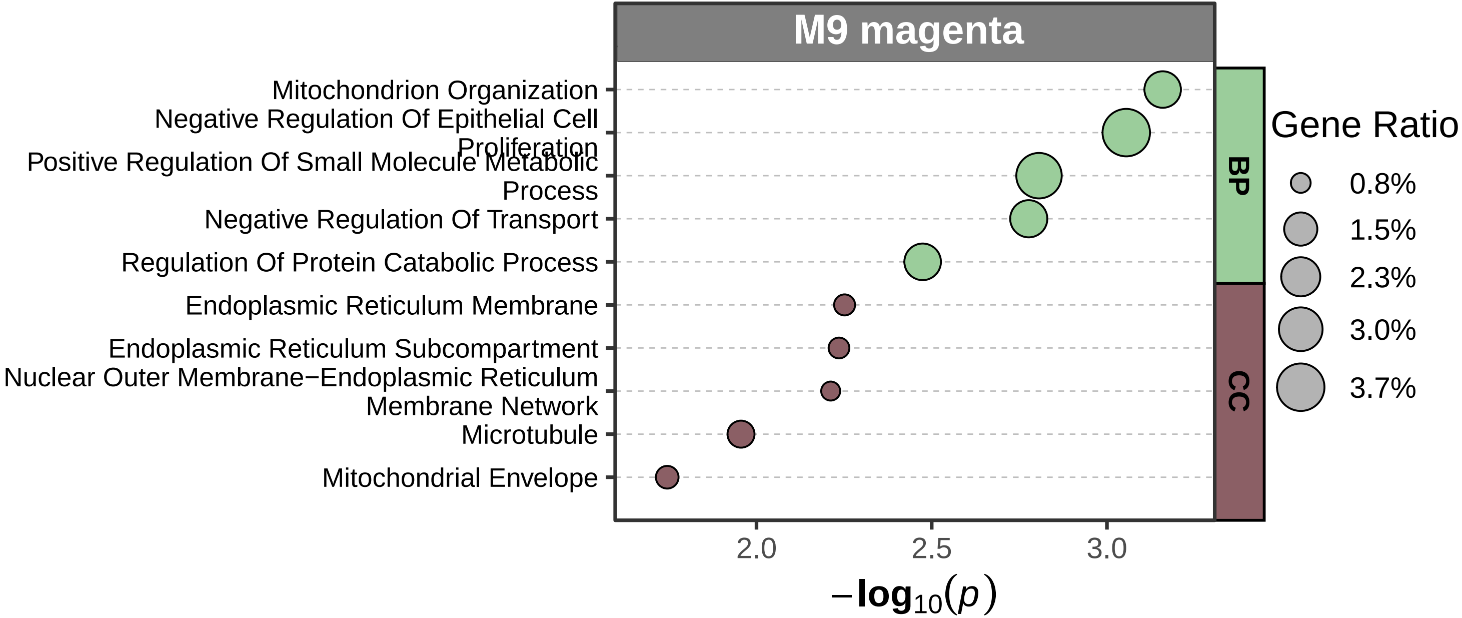
M9 magenta
Mitochondrion Organization
Negative Regulation Of Epithelial Cell Proliferation
Positive Regulation Of Small Molecule Metabolic Process
Negative Regulation Of Transport
Regulation Of Protein Catabolic Process
Endoplasmic Reticulum Membrane
Endoplasmic Reticulum Subcompartment
Nuclear Outer Membrane−Endoplasmic Reticulum Membrane Network
Microtubule
Mitochondrial Envelope
BP
CC
Gene Ratio
0.8%
1.5%
2.3%
3.0%
3.7%
2.0
2.5
3.0
$-\log_{10}(p)$

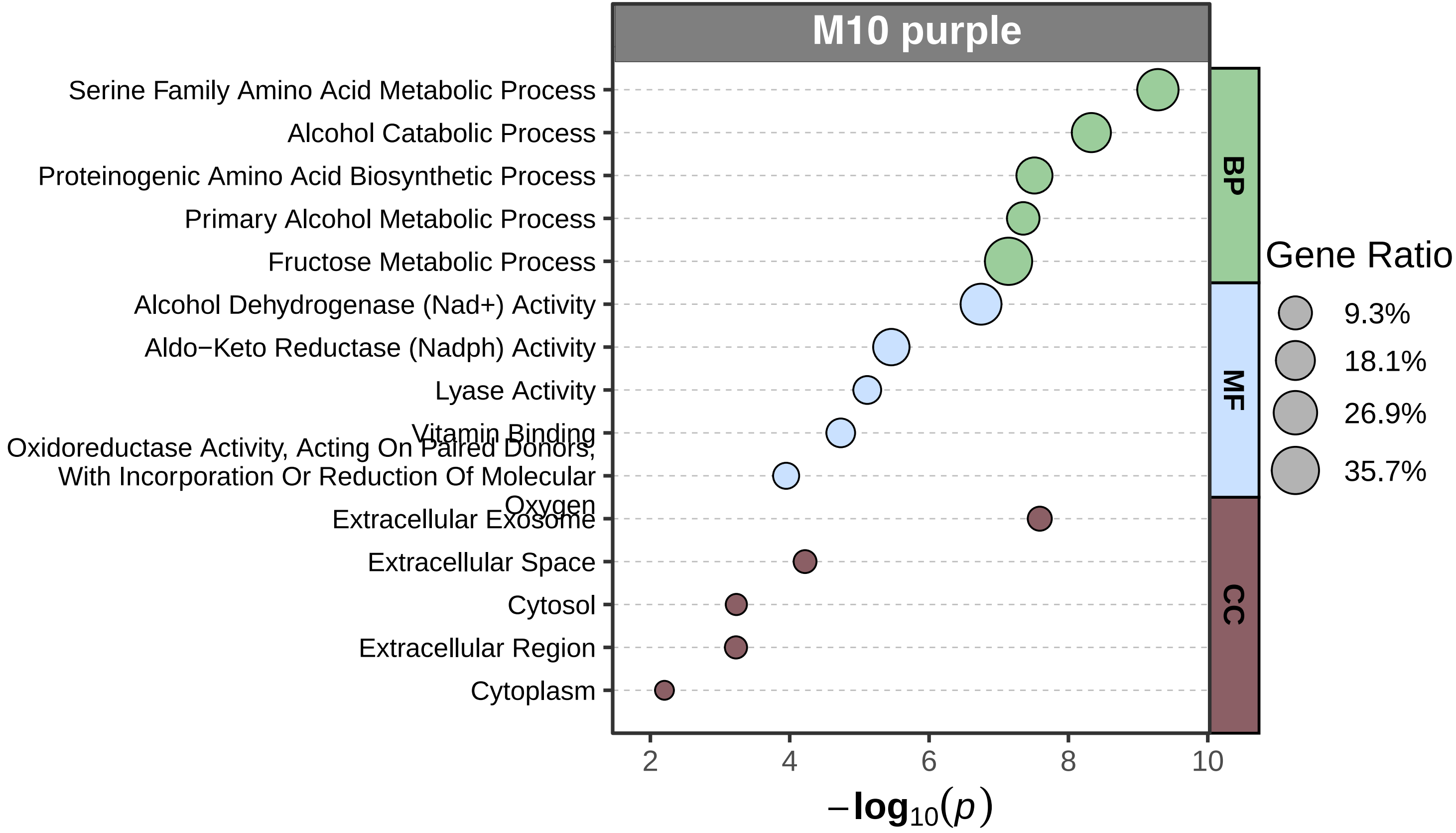

M10 purple
Serine Family Amino Acid Metabolic Process
Alcohol Catabolic Process
Proteinogenic Amino Acid Biosynthetic Process
Primary Alcohol Metabolic Process
Fructose Metabolic Process
Alcohol Dehydrogenase (Nad+) Activity
Aldo−Keto Reductase (Nadph) Activity
Lyase Activity
Vitamin Binding
Oxidoreductase Activity, Acting On Paired Donors, With Incorporation Or Reduction Of Molecular Oxygen
Extracellular Exosome
Extracellular Space
Cytosol
Extracellular Region
Cytoplasm
BP
MF
CC
Gene Ratio
9.3%
18.1%
26.9%
35.7%
2
4
6
8
10
$-\log_{10}(p)$

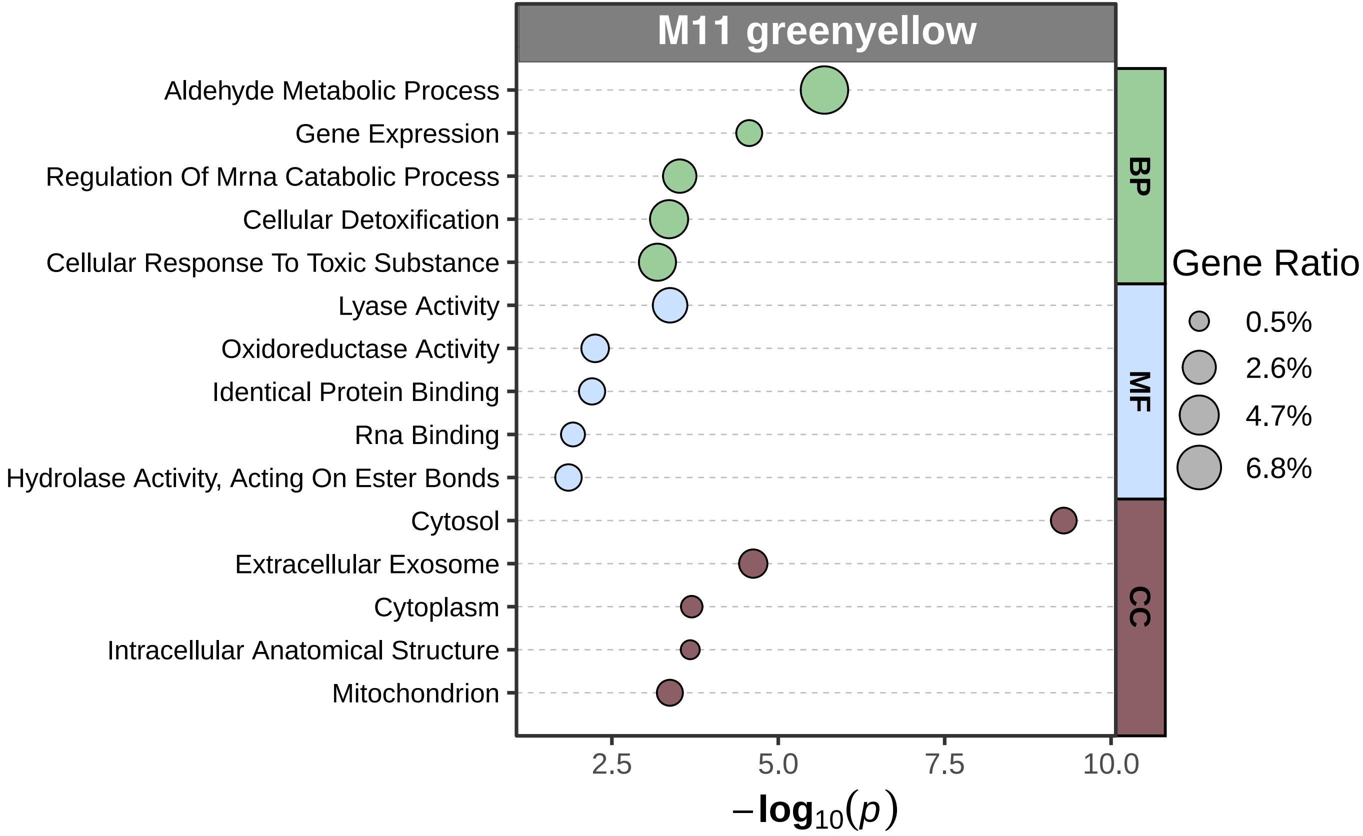
M11 greenyellow
Aldehyde Metabolic Process
Gene Expression
Regulation Of Mrna Catabolic Process
Cellular Detoxification
Cellular Response To Toxic Substance
Lyase Activity
Oxidoreductase Activity
Identical Protein Binding
Rna Binding
Hydrolase Activity, Acting On Ester Bonds
Cytosol
Extracellular Exosome
Cytoplasm
Intracellular Anatomical Structure
Mitochondrion
BP
MF
CC
Gene Ratio
0.5%
2.6%
4.7%
6.8%
2.5
5.0
7.5
10.0
$-\log_{10}(p)$

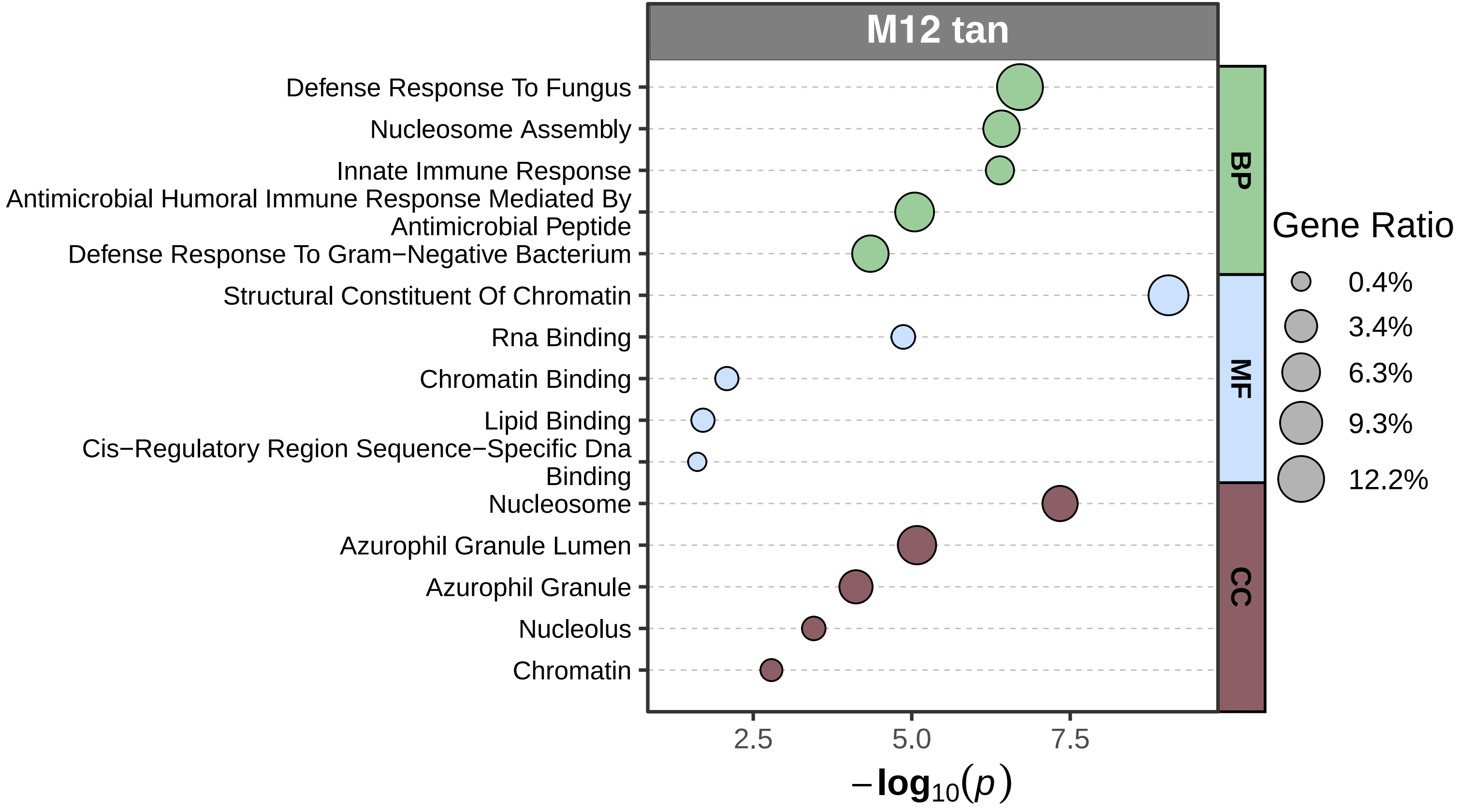
M12 tan
Defense Response To Fungus
Nucleosome Assembly
Innate Immune Response
Antimicrobial Humoral Immune Response Mediated By Antimicrobial Peptide
Defense Response To Gram−Negative Bacterium
Structural Constituent Of Chromatin
Rna Binding
Chromatin Binding
Lipid Binding
Cis−Regulatory Region Sequence−Specific Dna Binding
Nucleosome
Azurophil Granule Lumen
Azurophil Granule
Nucleolus
Chromatin
BP
MF
CC
Gene Ratio
0.4%
3.4%
6.3%
9.3%
12.2%
2.5
5.0
7.5
$-\log_{10}(p)$
